# New Phylogenetic Models Incorporating Interval-Specific Dispersal Dynamics Improve Inference of Disease Spread

**DOI:** 10.1101/2021.12.02.21267221

**Authors:** Jiansi Gao, Michael R. May, Bruce Rannala, Brian R. Moore

**Affiliations:** Department of Evolution and Ecology, University of California, Davis, Storer Hall, Davis, CA 95616, U.S.A.; Department of Integrative Biology, University of California, Berkeley, 3060 VLSB, Berkeley, CA 94720-3140, U.S.A.

**Keywords:** Phylodynamic models, Biogeographic history, Epidemiology, Phylogeography

## Abstract

Phylodynamic methods reveal the spatial and temporal dynamics of viral geographic spread, and have featured prominently in studies of the COVID-19 pandemic. Virtually all such studies are based on phylodynamic models that assume—despite direct and compelling evidence to the contrary—that rates of viral geographic dispersal are constant through time. Here, we: (1) extend phylodynamic models to allow both the average and relative rates of viral dispersal to vary independently between pre-specified time intervals; (2) implement methods to infer the number and timing of viral dispersal events between areas; and (3) develop statistics to assess the absolute fit of discrete-geographic phylodynamic models to empirical datasets. We first validate our new methods using simulations, and then apply them to a SARS-CoV-2 dataset from the early phase of the COVID-19 pandemic. We show that: (1) under simulation, failure to accommodate interval-specific variation in the study data will severely bias parameter estimates; (2) in practice, our interval-specific discrete-geographic phylodynamic models can significantly improve the relative and absolute fit to empirical data; and (3) the increased realism of our interval-specific models provides qualitatively different inferences regarding key aspects of the COVID-19 pandemic—revealing significant temporal variation in global viral dispersal rates, viral dispersal routes, and the number of viral dispersal events between areas—and alters interpretations regarding the efficacy of intervention measures to mitigate the pandemic.

## Introduction

Phylodynamic methods encompass a suite of models for inferring various aspects of pathogen biology, including: (1) patterns of variation in demography through time (Drummond et al., 2005; Gill et al., 2016, 2013; Minin et al., 2008); (2) the history of geographic spread either over continuous space (Gill et al., 2017; Lemey et al., 2010; Pybus et al., 2012) or among a set of discrete-geographic areas (Edwards et al., 2011; Lemey et al., 2009), and; (3) the interaction between demography and geographic history (De Maio et al., 2015; Kühnert et al., 2016; Müller et al., 2019, 2017). Our focus here is on discrete-geographic phylodynamic models (Edwards et al., 2011; Lemey et al., 2009). These phylodynamic methods have been used extensively to understand the spatial and temporal spread of disease outbreaks and have played a central role for inferring key aspects of the COVID-19 pandemic, such as the geographic location and time of origin of the disease, the rates and geographic routes by which it spread, and the efficacy of various mitigation measures to limit its geographic expansion (Alpert et al., 2021; Bedford et al., 2020; Candido et al., 2020; Davies et al., 2021; Dellicour et al., 2021; Douglas et al., 2021; du Plessis et al., 2021; Fauver et al., 2020; Kraemer et al., 2021; Lemey et al., 2021; Müller et al., 2021; Nadeau et al., 2021; Tegally et al., 2021; Washington et al., 2021; Wilkinson et al., 2021; Worobey et al., 2020).

These phylodynamic methods adopt an explicitly probabilistic approach that model the process of viral dispersal among a set of discrete-geographic areas (Baele et al., 2017). The observations include the times and locations of viral sampling, and the genomic sequences of the sampled viruses. These data are used to estimate the parameters of phylodynamic models, which include a dated phylogeny of the viral samples, the global dispersal rate (the average rate of dispersal among all geographic areas), and the relative dispersal rates (the dispersal rate between each pair of geographic areas).

The vast majority (651 of 666, 97.7%; Fig. S1) of discrete-geographic phylodynamic studies are based on the earliest models (Edwards et al., 2011; Lemey et al., 2009), which assume that viral dispersal dynamics—including the average and relative rates of viral dispersal—remain constant over time (see the caption of Fig. S1 for the search query we used to identify these studies). However, real-world observations indicate that the average and/or relative rates of viral dispersal inevitably vary during disease outbreaks. For example, relative rates of viral dispersal typically change as a disease is introduced to (and becomes prevalent in) new areas, and begins dispersing from those areas to other areas. Dispersal dynamics are also generally impacted by the initiation (or alteration or cessation) of area-specific mitigation measures (*e*.*g*., domestic shelter-in-place policies) that change the rate of viral transmission within an area and the relative rate of dispersal to other areas. Similarly, average rates of viral dispersal may change in response to the initiation (or alteration or cessation) of more widespread intervention efforts—*e*.*g*., multiple area-specific mitigation measures, international-travel bans—that collectively impact the overall viral dispersal rate.

In this paper, we: (1) extend discrete-geographic phylodynamic models to allow both the average and relative dispersal rates to vary independently across pre-specified time intervals; (2) enable stochastic mapping under these interval-specific discrete-geographic phylodynamic models to estimate the number and timing of viral dispersal events between areas, and; (3) develop statistics to assess the absolute fit of discrete-geographic phylodynamic models to empirical datasets. We first validate the theory and implementation of our new phylodynamic methods using analyses of simulated data, and then provide an empirical demonstration of these methods with analyses of a SARS-CoV-2 dataset from the early phase of the COVID-19 pandemic.

## Extending Phylodynamic Models

### Anatomy of interval-specific discrete-geographic phylodynamic models

Phylodynamic models of dispersal include two main components (Fig. 1): a *phylogenetic model* that allows us to estimate a dated phylogeny for the sampled viruses, Ψ, and a *biogeographic model* that describes the history of viral dispersal over the tree as a continuous-time Markov chain. For a geographic history with *k* discrete areas, this stochastic process is fully specified by a *k ×k* instantaneous-rate matrix, **Q**, where an element of the matrix, *q*_*ij*_, is the instantaneous rate of change between state *i* and state *j* (*i*.*e*., the instantaneous rate of dispersal from area *i* to area *j*). We rescale the **Q** matrix such that the average rate of dispersal between all areas is *µ*; this represents the average rate of viral dispersal among all areas (Yang, 2014).

**Fig. 1.**
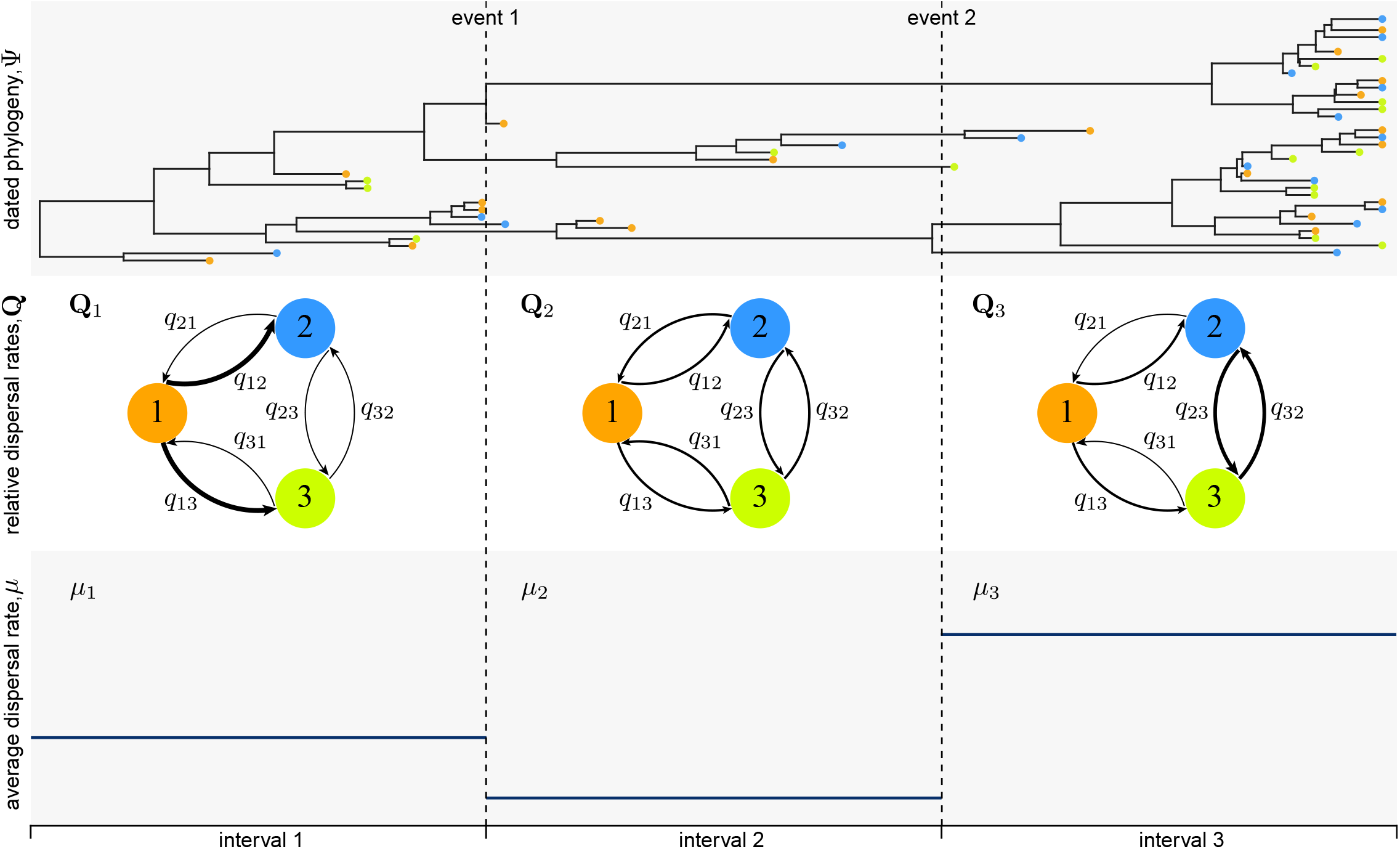
Interval-specific discrete-geographic phylodynamic models accommodate variation in the process of viral dispersal. Phylodynamic models include two main components: a *phylogenetic model* that specifies the relationships and divergence times of the sampled viruses, Ψ (top panel), and a *biogeographic model* that describes the history of viral dispersal among a set of discrete-geographic areas—here, areas 1 (orange), 2 (blue), and 3 (green)—from the root to the tips of the dated viral tree. Parameters of the biogeographic model include an instantaneous-rate matrix, **Q**, that specifies relative rates of viral dispersal between each pair of areas (here, each element of the matrix, *q*_*ij*_, is represented as an arrow that indicates the direction and relative dispersal rate from area *i* to area *j*; middle panel), and a parameter that specifies the average rate of viral dispersal between all areas, *µ* (lower panel). Although most phylodynamic studies assume that the process of viral dispersal is constant through time, disease outbreaks are typically punctuated by events that impact the average and/or relative rates of viral dispersal among areas. Here, for example, the history involves two events (*e*.*g*., mitigation measures) that define three intervals, where both **Q** and *µ* are impacted by each of these events, such that the interval-specific parameters are (**Q**_1_,**Q**_2_,**Q**_3_) and (*µ*_1_,*µ*_2_,*µ*_3_). Our framework allows investigators to specify discrete-geographic phylodynamic models with two or more intervals, where each interval has independent relative and/or average dispersal rates, which are then estimated from the data.

We could specify alternative biogeographic models based on the assumed constancy of the dispersal process. For example, the simplest possible model assumes that the average dispersal rate, *µ*, and the relative dispersal rates, **Q**, remain constant over the entire history of the viral outbreak. Typically, viral outbreaks are punctuated by events that are likely to impact the average rate of viral dispersal (*e*.*g*., the onset of an international-travel ban) and/or the relative rates of viral dispersal between pairs of areas (*e*.*g*., the initiation of localized mitigation measures). We can incorporate information on such events into our phylodynamic inference by specifying interval-specific models. That is, the investigator specifies the number of intervals, the boundaries between each interval, and the parameters that are specific to each interval according to the presumed changes in the history of viral dispersal. For example, we might specify an interval-specific model (Membrebe et al., 2019) that assumes that the average rate of viral dispersal varies among two or more intervals (while assuming that the relative rates of viral dispersal remain constant across intervals). Conversely, an interval-specific model (Bielejec et al., 2014) might allow the relative rates of viral dispersal to vary among two or more time intervals (while assuming that the average rate of viral dispersal remains constant across intervals).

Alternatively, a more complex interval-specific model might allow both the average rate of viral dispersal and the relative rates of viral dispersal to vary among two or more intervals. We extend interval-specific discrete-geographic phylodynamic models to allow *both* the relative *and* average dispersal rates to vary independently across two or more pre-defined intervals. Here, we describe how to compute transition probabilities, perform inference, simulate histories, and assess the absolute fit of the interval-specific models.

### Computing Transition Probabilities

The transition-probability matrix, **P**, describes the probability of transitioning from state *i* to state *j* (*i*.*e*., dispersing from area *i* to area *j*) along a branch with a finite duration; importantly, a branch may span two or more intervals with different relative and/or absolute dispersal rates.

#### Allowing average dispersal rates to vary across intervals

Under a constant-rate discrete-geographic phylodynamic model, the transition-probability matrix for a branch is **P** =exp(**Q***v*), where *v* = *µt* represents the expected number of dispersal events on a branch of duration *t* with an average dispersal rate *µ*. However, under a phylodynamic model with interval-specific average dispersal rates (Membrebe et al., 2019)—which allows the average dispersal rate to vary among intervals, but assumes that relative dispersal rates are constant across all intervals— a given branch in a phylogeny may span two or more intervals with different average dispersal rates (“average-rate intervals”). The transition-probability matrix for the branch is then computed as the matrix exponential:

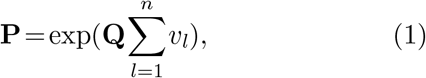

where **Q** is the instantaneous-rate matrix, *n* is the number of average-rate intervals spanned by the branch, and *v*_*l*_ is the expected number of dispersal events on the branch in average-rate interval *l*. Recall that *v*_*l*_ = *µ*_*l*_*t*_*l*_, where *µ*_*l*_ is the average dispersal rate during interval *l* and *t*_*l*_ is the time spent in interval *l*.

#### Allowing relative dispersal rates to vary across intervals

Under a phylodynamic model with interval-specific relative dispersal rates (Bielejec et al., 2014)— which allows the instantaneous-rate of dispersal between each pair of areas to vary among intervals, but assumes that the average dispersal rate is constant across all intervals—a given branch may span two or more intervals with different **Q** matrices (“relative-rate intervals”). In this case, the transition-probability matrix for each relative-rate interval *l*, **P**_*l*_, is computed as:

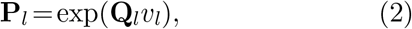

where **Q**_*l*_ is the instantaneous-rate matrix in relative-rate interval *l*, and *v*_*l*_ = *µt*_*l*_ is the average dispersal rate multiplied by the time spent in interval *l*. The transition-probability matrix for the entire branch is then computed as the matrix product of interval-specific transition-probability matrices:

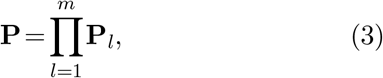

where *m* is the number of relative-rate intervals spanned by the branch.

#### Allowing average and relative dispersal rates to vary across intervals

We combine the two approaches described above to compute transition-probability matrices under an interval-specific model that allows both the average dispersal rate and the relative dispersal rates to vary independently among intervals. Let a given branch span *m* relative-rate intervals. The expected number of dispersal events in each such interval *l, v*_*l*_, is computed as:

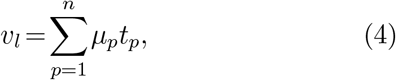

where *n* is the number of average-rate intervals spanned by interval *l, µ*_*p*_ is the dispersal rate in average-rate interval *p*, and *t*_*p*_ is the time spent in average-rate interval *p*. We then substitute equation (4) into equation (2), and apply equation (3) as normal to compute the transition-probability matrix for the entire branch. An example computation is illustrated in Figure 2 for a scenario in which a branch spans two different relative-rate intervals and three different average-rate intervals.

**Fig. 2.**
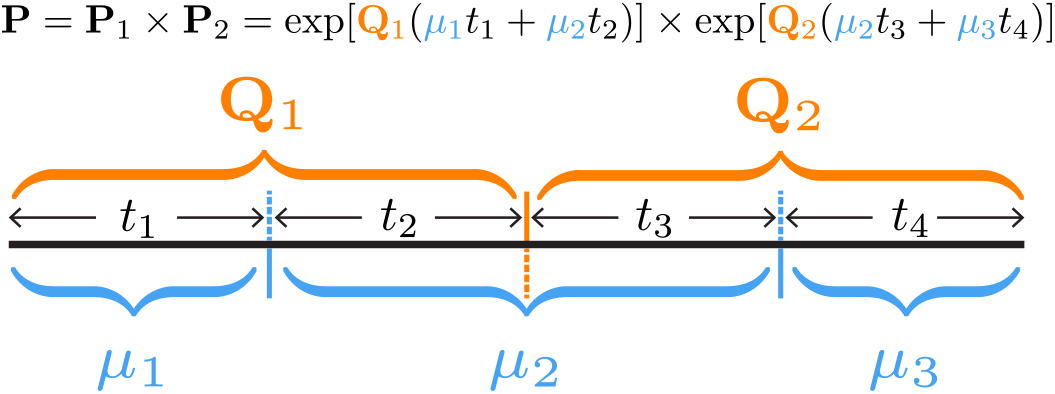
Computing the transition-probability matrix for a branch spanning intervals where both the average and relative dispersal rates vary. An example illustrating the transition-probability matrix computation for a branch spanning two relative-rate intervals (**Q**_1_ and **Q**_2_) and three average-rate intervals (*µ*_1_,*µ*_2_,*µ*_3_).

We modified BEAST source code to implement the above equation for computing **P** under our interval-specific phylodynamic models that allow both *µ* and **Q** to vary independently among two or more pre-specified intervals.

### Inference under interval-specific discrete-geographic phylodynamic models

We estimate parameters of the interval-specific models within a Bayesian statistical framework. Specifically, we use numerical algorithms—Markov chain Monte Carlo (MCMC) simulation—to approximate the joint posterior probability distribution of the phylodynamic model parameters—the dated phylogeny, Ψ, the set of relative dispersal rates, **Q**, and the average dispersal rates, *µ*—from the study data (*i*.*e*., the location and times of viral sampling, and the genomic sequences of the sampled viruses).

### Simulating dispersal histories under interval-specific discrete-geographic phylodynamic models

We have also implemented numerical algorithms— stochastic mapping—to simulate histories of viral dispersal under the interval-specific models; these methods allow us to estimate the number of dispersal events between a specific pair of areas, the number of dispersal events from one area to a set of two or more areas, and the total number dispersal events among all areas. Stochastic mapping—initially proposed by Nielsen (2002; see also Bollback, 2006; Hobolth and Stone, 2009; Huelsenbeck et al., 2003; Minin and Suchard, 2008)—is commonly used to sample dispersal histories over branches of a phylogeny. Here, we extend this approach to sample dispersal histories under our interval-specific models.

Let a given branch start at time *T*_0_ with state *i* and end at time *T*_*m*_ with state *k*. Further, let the dispersal process change (either by changing the average or relative dispersal rates) *m−*1 times on the branch at times *{T*_1_,…,*T*_*m−*1_*}*, resulting in *m* intervals. For interval *l*, denote the average dispersal rate as *µ*_*l*_, the instantaneous-rate matrix as **Q**_*l*_, and the duration as *t*_*l*_. We simulate a dispersal history along this branch using a two-step procedure: (1) we first sample the state at each of the *m−*1 time points, and; (2) we then simulate the history between each time point, conditional on the states sampled in the first step.

To simulate the states at each time point, we first compute a transition-probability matrix for each interval:

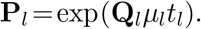

We then calculate the probability of state *j* at the first time point, *T*_1_, given that the branch begins in state *i* and ends in state *k*, as:

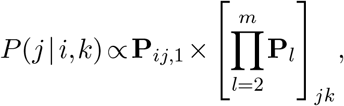

where the first term is the probability of transitioning from state *i* (the state at the beginning of the branch) to state *j* at the first time point, and the second term is the probability of transitioning from state *j* to state *k* (the state at the end of the branch) over the remaining time intervals. We compute this for each state *j*, and sample the state in proportion to these probabilities. We then repeat this process for each remaining time point, recursively conditioning on the state sampled at the previous time point and the state at the end of the branch.

Second, we simulate histories within each interval. For a given time interval, we simulate histories conditional on the start and end states generated in the first step using the uniformization algorithm described by Hobolth and Stone (2009).

### Assessing the absolute fit of interval-specific discrete-geographic phylodynamic models

For a given phylodynamic study, we might wish to consider several candidate interval-specific models (where each candidate model specifies a unique number of intervals, set of interval boundaries, and/or interval-specific parameters). Comparing the fit of these competing discrete-geographic phylodynamic models to the data offers two benefits: (1) confirming that our inference model adequately describes the process that gave rise to data will improve the accuracy of the corresponding inferences (*i*.*e*., estimates of relative and/or average dispersal-rate parameters and viral dispersal histories), and; (2) comparing alternative models provides a means to objectively test hypotheses regarding the impact of events on the history of viral dispersal (*i*.*e*., by assessing the relative fit of data to competing models that include/exclude the impact of a putative event on the average and/or relative viral dispersal rates). We can assess the *relative* fit of two or more candidate phylodynamic models to a given dataset using Bayes factors; this requires that we first estimate the marginal likelihood for each model (which represents the average fit of a model to a dataset), and then compute the Bayes factor as twice the difference in the log marginal likelihoods of the competing models (Kass and Raftery, 1995).

However, even the best candidate model may fail to provide an adequate description of the process that gave rise to our study data. We can leverage our ability to simulate histories under the interval-specific models to develop new methods to assess the *absolute* fit of a candidate discrete-geographic phylodynamic model using posterior-predictive assessment (Gelman et al., 1996). This Bayesian approach for assessing model adequacy is based on the following premise: if our inference model provides an adequate description of the process that gave rise to our observed data, then we should be able to use that model to simulate datasets that resemble our original data. The resemblance between the observed and simulated datasets is quantified using a summary statistic. Accordingly, posterior-predictive simulation requires: (1) the ability to simulate geographic datasets under interval-specific discrete-geographic phylodynamic models for a given set of parameter values, and; (2) summary statistics that allow us to compare the resulting simulated datasets to the observed dataset. We describe each of these components below.

### Simulating under interval-specific discrete-geographic phylodynamic models

We draw *m* random samples from the joint posterior distribution of the model; each sample *i* consists of a fully specified phylodynamic model, *θ*_*i*_ = *{*Ψ_*i*_,**Q**_*i*_,*µ*_*i*_*}*. For each sample, we simulate a new geographic dataset on the sampled tree, Ψ_*i*_, given the sampled parameters of the geographic model, *{***Q**_*i*_,*µ*_*i*_*}*; we label the newly simulated dataset 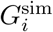. Under a constant-rate discrete-geographic phylodynamic model, we simulate full dispersal histories forward in time over a tree using the `sim.history()` function in the `R` package `phytools` (Revell, 2012). We implemented an extension of the `sim.history()` function to simulate dispersal histories under interval-specific discrete-geographic phylodynamic models. These functions allow us to perform posterior-predictive simulation to assess the adequacy of both the constant-rate and the interval-specific models.

#### Summary statistics

We define a summary statistic, which we generically denote *T* (*G* | *θ*_*i*_), where *G* is either the simulated or observed dataset. For each simulated dataset, we compute a discrepancy statistic,

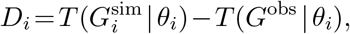

where *G*^obs^ is the observed geographic dataset and *G*^sim^ is a simulated dataset. We developed two summary statistics to assess the adequacy of interval-specific discrete-geographic phylodynamic models: (1) the *parsimony statistic*, and; (2) the *tipwise-multinomial statistic*. The parsimony statistic is calculated as the difference in the parsimony score for the observed areas and the simulated areas across the tips of the tree (where the parsimony score is the minimum number of dispersal events required to explain the distribution of areas across the tips of a tree). We compute parsimony scores using the `parsimony()` function in the R package, `phangorn` (Schliep, 2010). The tipwise-multinomial statistic is inspired by the multinomial statistic that was proposed by Goldman (1993) and later used by Bollback (2002) to assess the adequacy (absolute fit) of substitution models to sequence alignments. Our tipwise statistic treats the set of states (areas) across the tips of the tree as an outcome of a multinomial trial. Specifically, we calculate the tipwise-multinomial statistic as the difference in the multinomial probabilities for the observed set of areas versus the simulated set of areas across the tips of the tree. We calculate each multinomial probability as:

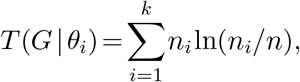

where *n* is the number of tips in the tree, and *n*_*i*_ is the number of tips that occur area *i*.

#### Time-slice summary statistics

To assess the ability of discrete-geographic phylodynamic models to describe the temporal distribution of dispersal events, we extend the parsimony and tipwise-multinomial summary statistics to assess time slices of the geographic history^1^. We calculate these summary statistics for *k* pre-specified time slices, resulting in *k* parsimony statistics and *k* tipwise-multinomial statistics for each simulated dataset. We compute the time-slice variant of the parsimony summary statistic as follows: (1) we first infer the most-parsimonious dispersal history (*i*.*e*., the minimum number of dispersal events) for a given simulated dataset and the observed dataset using the `ancestral.pars()` function in the `R` package, `phangorn` (Schliep, 2010); (2) we then assign each inferred dispersal event to one of the *k* time slices based on the time span of the branch along which the dispersal event was inferred (when a dispersal event is inferred to occur along a branch that spans two or more time slices, we locate the event uniformly along the branch, and then assign it to the corresponding slice), and finally; (3) we compute the difference in the number of dispersal events between the simulated and observed dataset for each time slice. We compute the time-slice variant of the tipwise-multinomial summary statistic in a similar manner; *i*.*e*., we first find the set of tips in each time slice, and then compute the tipwise-multinomial statistic for that time slice (as described above) for the corresponding set of tips. Further details regarding the computation of these summary statistics are available in an R script provided in our GitHub and Dryad repositories.

### Simulation Study

We performed a simulation study to explore the statistical behavior of the interval-specific phylodynamic models. Specifically, the goals of this simulation study are to assess: (1) our ability to perform reliable inference under interval-specific models; (2) the impact of model misspecification, and; (3) our ability to identify the correct model. To this end, we simulated 200 geographic datasets under each of two models: the first assumes a constant *µ* and **Q** (1*µ*1**Q**), and the second allows *µ* and **Q** to vary over two intervals (2*µ*2**Q**). The parameter values used in the simulation are from empirical analyses of a SARS-CoV-2 dataset (with 1271 viral sequences sampled from three coarsely aggregated geographic areas) under each corresponding model. For each simulated dataset, we separately inferred the history of viral dispersal under each model, resulting in four true:inference model combinations: 1*µ*1**Q**:1*µ*1**Q**, 2*µ*2**Q**:2*µ*2**Q**, 1*µ*1**Q**:2*µ*2**Q**, and 2*µ*2**Q**:1*µ*1**Q**. We provide detailed descriptions of the simulation analyses and results in Section S2 of the supplementary material.

#### Ability to reliably estimate parameters of interval-specific discrete-geographic phylodynamic models

Interval-specific discrete-geographic phylodynamic models are inherently more complex than their constant-rate counterparts, and therefore contain many more parameters that must be inferred from geographic datasets that contain minimal information; these datasets only include a single observation (*i*.*e*., the area in which each virus was sampled). These considerations raise concerns about our ability to reliably estimate parameters of interval-specific phylodynamic models. Encouragingly, when the inference model is correctly specified (*i*.*e*., where both the true and inference models include [or exclude] interval-specific parameters, 2*µ*2**Q**:2*µ*2**Q** and 1*µ*1**Q**:1*µ*1**Q**), our simulation study demonstrates that estimates under interval-specific models are as reliable as those under constant-rate models (Fig. 3, green, blue). Moreover, when the inference model is overspecified (*i*.*e*., it includes interval-specific parameters not included in the true model) inferences are comparable to those under correctly specified models (Fig. 3, purple). However, when the inference model is underspecified (*i*.*e*., it excludes interval-specific parameters of the true model) inferences are severely biased estimates (Fig. 3, orange).

**Fig. 3.**
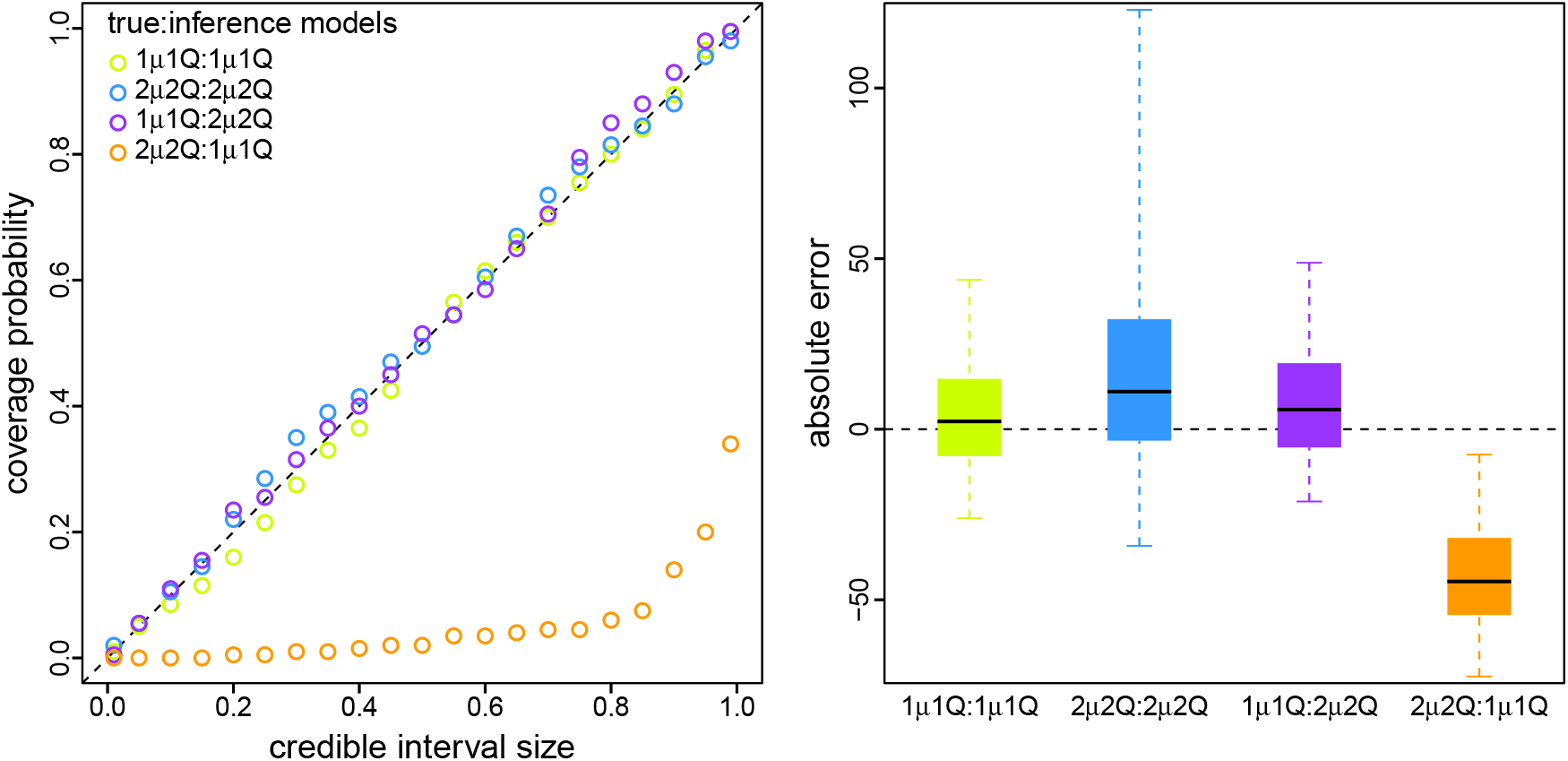
Simulation demonstrates that reliable inference of viral dispersal history requires a correctly specified discrete-geographic phylodynamic model. We simulated 200 geographic datasets under each of two models: one that assumed a constant *µ* and **Q** (1*µ*1**Q**), and one that allowed *µ* and **Q** to vary over two intervals (2*µ*2**Q**). For each simulated dataset, we separately inferred the total number of dispersal events under each model, resulting in four true:inference model combinations (1*µ*1**Q**:1*µ*1**Q**, 2*µ*2**Q**:2*µ*2**Q**, 1*µ*1**Q**:2*µ*2**Q**, and 2*µ*2**Q**:1*µ*1**Q**). Left) For each combination of true and inference model, we computed the coverage probability (the frequency with which the true number of dispersal events was contained in the corresponding *X*% credible interval; y-axis) as a function of the size of the credible interval (x-axis). When the model is true, we expect the coverage probability to be equal to the size of the credible interval (Cook et al., 2006). As expected, coverage probabilities fall along the one-to-one line when the model is correctly specified (green and blue). Moreover, coverage probabilities are also appropriate when the inference model is overspecified (*i*.*e*., the inference model includes interval-specific parameters not included in the true model; purple). However, coverage probabilities are extremely unreliable when the inference model is underspecified (*i*.*e*., the inference model excludes interval-specific parameters of the true model; orange). Right) For each true:inference model combination, we summarized the absolute error (estimated minus true number of dispersal events) as boxplots (median [horizontal bar], 50% probability interval [boxes], and 95% probability interval [whiskers]). Again, when the model is underspecified (orange) inferences are strongly biased compared to those under the correctly specified (green and blue) and overspecified (purple) models.

#### Ability to accurately identify an appropriately specified discrete-geographic phylodynamic model

Our simulation study demonstrates the importance of identifying scenarios where an inference model is underspecified; failure to accommodate interval-specific variation in the study data can severely bias parameter estimates. Fortunately, our simulation study demonstrates that we can reliably identify when a given model is correctly specified, overspecified, or underspecified using a combination of Bayes factors (to assess the relative fit of competing models to the data; Fig. 4, left) and posterior-predictive simulation (to assess the absolute fit of each candidate model to the data; Fig. 4, right). Using a combination of Bayes factors and posterior-predictive simulation allows us to not only identify the best of the candidate models, but also to ensure that the best model provides an adequate description of the true process that gave rise to our study data.

**Fig. 4.**
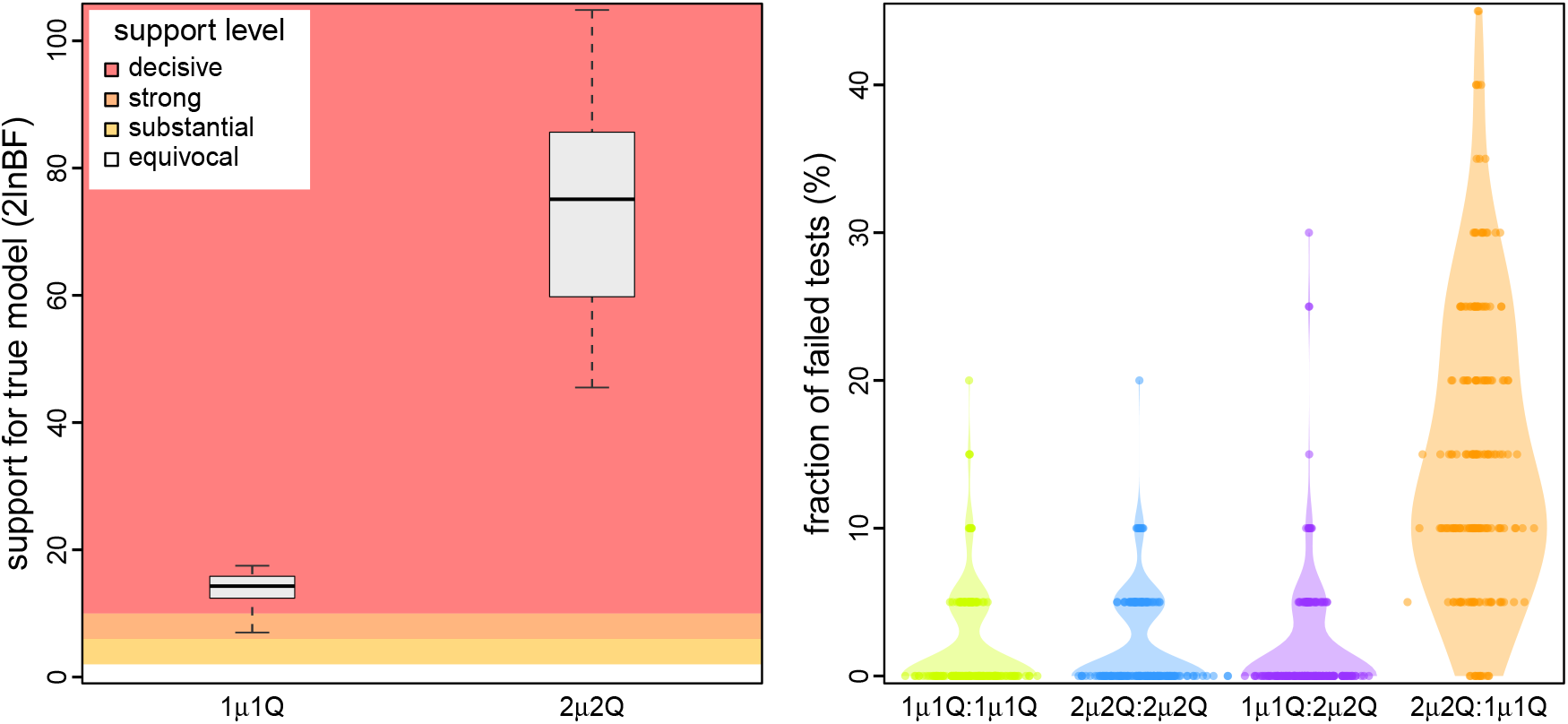
Simulation demonstrates our ability to accurately identify a correctly specified discrete-geographic phylodynamic model. We assessed the relative and absolute fit of alternative models to the simulated datasets described in Fig. 3. Left) For each simulated dataset, we compared the relative fit of the true and alternative models using Bayes factors. The boxplots summarize Bayes factors for datasets simulated under the constant-rate (1*µ*1**Q**, left) and interval-specific (2*µ*2**Q**, right) models, which demonstrate that we are able to decisively identify the true phylodynamic model. Right) For each combination of true:inference model, we assessed absolute model fit using posterior-predictive simulation with a set of 20 summary statistics. Each dot represents the fraction of those 20 summary statistics for which the corresponding inference model provides an inadequate fit to a single simulated dataset. The violin plots summarize the distribution of these values for all datasets under each true:inference model combination. As expected, the true model is overwhelmingly inferred to be adequate (green and blue). Encouragingly, model overspecification appears to have a negligible impact on model adequacy (purple). By contrast, an underspecified model severely impacts model adequacy (orange).

### Empirical Application

We demonstrate our new phylodynamic methods with analyses of all publicly available SARS-CoV-2 genomes sampled during the early phase of the COVID-19 pandemic (with 2598 viral genomes collected from 23 geographic areas between Dec. 24, 2019–Mar. 8, 2020 [downloaded from GISAID, Shu and McCauley, 2017]). We used our study dataset to estimate the parameters of—and assess the relative and absolute fit to—nine candidate discrete-geographic phylodynamic models. These models assign interval-specific parameters—for the average rate of viral dispersal, *µ*, and/or relative rates of viral dispersal, **Q**—to one, two, four, or five pre-specified time intervals; *i*.*e*., 1*µ*1**Q**, 2*µ*1**Q**, 1*µ*2**Q**, 2*µ*2**Q**, 4*µ*1**Q**, 1*µ*4**Q**, 4*µ*4**Q**, 5*µ*5**Q**, and 5*µ*5**Q**^*\**^. We specified interval boundaries based on external information regarding events within the study period that might plausibly impact viral dispersal dynamics, including: (A) start of the Spring Festival travel season in China (the highest annual period of domestic travel, Jan. 12); (B) onset of mitigation measures in Hubei province, China (Jan. 26); (C) onset of international air-travel restrictions against China (Feb. 2), and; (D) relaxation of domestic travel restrictions in China (Feb. 16). Phylodynamic models with two intervals include event C, models with four intervals include events A, C, and D, and the 5*µ*5**Q** model includes all four events. The final candidate model, 5*µ*5**Q**^*\**^, includes five arbitrary and uniform (14-day) intervals. We provide detailed descriptions of our empirical data collection, analyses, and results in Section S3 of the supplementary material.

#### An interval-specific model best describes viral dispersal in the early phase of the pandemic

Our phylodynamic analyses of the SARS-CoV-2 dataset reveal that the early phase of the COVID-19 pandemic exhibits significant variation in both the average and relative rates of viral dispersal over four time intervals. Bayes factor comparisons (Fig. 5, left) demonstrate that the 4*µ*4**Q** interval-specific model is decisively preferred both over all less complex candidate models—including models that allow *either* the average dispersal rate *or* relative dispersal rates to vary over the same four intervals (4*µ*1**Q** and 1*µ*4**Q**, respectively)—and also over more complex candidate models (5*µ*5**Q**, and 5*µ*5**Q**^*\**^). Posterior-predictive analyses (Fig. 5, right) demonstrate that the preferred model, 4*µ*4**Q**, also provides an adequate description of the process that gave rise to our SARS-CoV-2 dataset. Below, we will use the preferred (4*µ*4**Q**) interval-specific phylodynamic model to explore various aspects of viral dispersal during the early phase of the COVID-19 pandemic and—for the purposes of comparison—we also present corresponding results inferred using the (underspecified) constant-rate (1*µ*1**Q**) phylodynamic model.

**Fig. 5.**
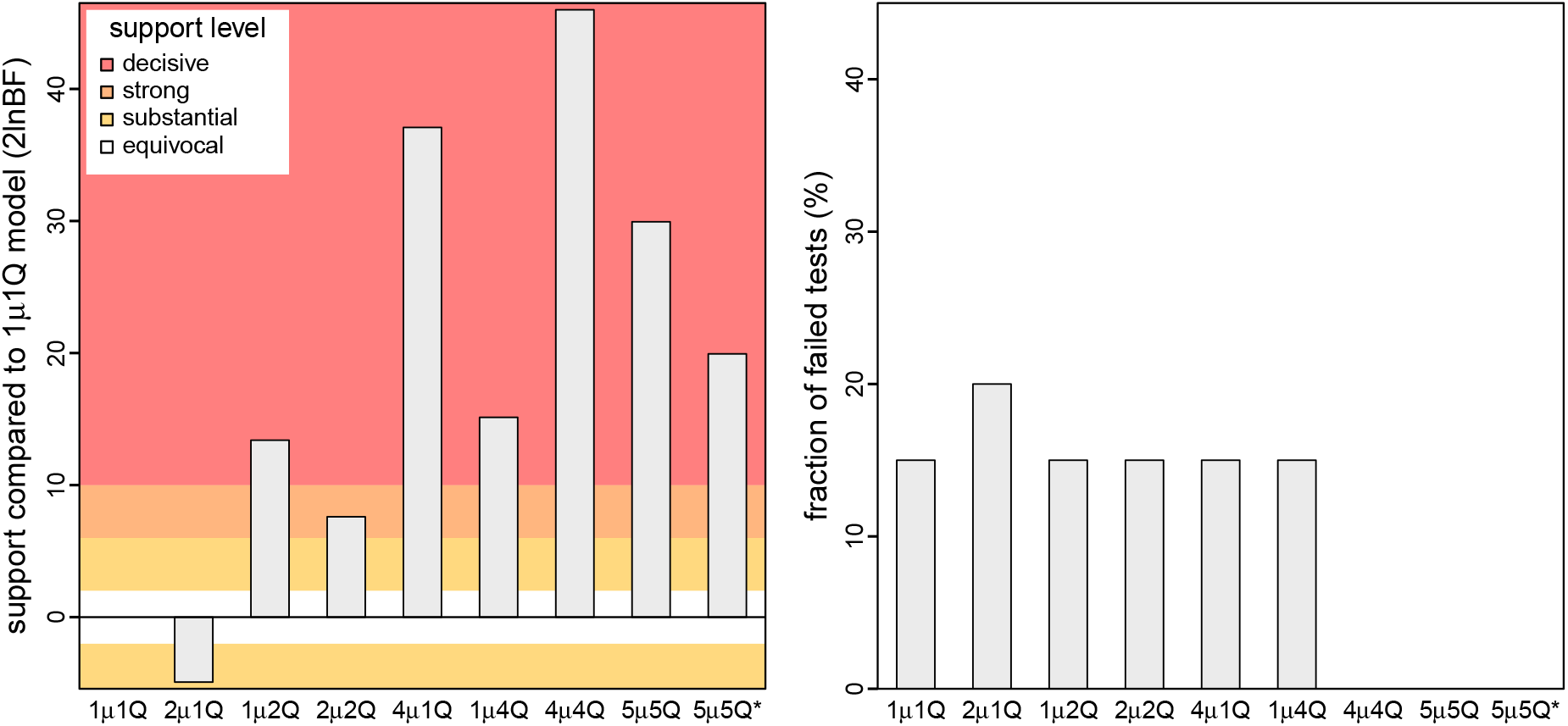
An interval-specific model provides the best relative and absolute fit to our SARS-CoV-2 dataset. We assessed the relative and absolute fit of nine candidate discrete-geographic phylodynamic models to our study dataset (comprised of all publicly available SARS-CoV-2 genomes from the early phase of the COVID-19 pandemic). Left) We compared the relative fit of each candidate model to the constant-rate (1*µ*1**Q**) phylodynamic model using Bayes factors, which indicate that the 4*µ*4**Q** interval-specific model outcompetes both less complex and more complex models. Right) We performed posterior-predictive simulation for each candidate model using 20 summary statistics, plotting the fraction of those summary statistics indicating that a given candidate model was inadequate. Our results indicate that three candidate models (4*µ*4**Q**, 5*µ*5**Q**, and 5*µ*5**Q**^*\**^) provide an adequate fit to our SARS-CoV-2 dataset. The simplest of these adequate models (4*µ*4**Q**) also provides the best relative fit. Collectively, these results identify the 4*µ*4**Q** model as the clear choice for phylodynamic analyses of our study dataset.

#### Variation in global viral dispersal rates

Between late 2019 and early March, 2020, COVID-19 emerged (in Wuhan, China) and established a global distribution—with reported cases in 83% of the study areas by this date (WHO, 2020)—despite the implementation of numerous intervention efforts to slow the spread of the causative SARS-CoV-2 virus (Hsiang et al., 2020). This crucial early phase of the pandemic provides a unique opportunity to explore the dispersal dynamics that led to the worldwide establishment of the virus and to assess the efficacy of key public-health measures to mitigate the spread of COVID-19. The constant-rate (1*µ*1**Q**) model infers a static rate of global viral dispersal throughout the study period (Fig. 6, orange). By contrast, inferences under the preferred (4*µ*4**Q**) model reveal significant variation in global viral dispersal rates over four intervals, exhibiting both increases and decreases over the early phase of the pandemic (Fig. 6, dark blue). The significant decrease in the global viral dispersal rate between the second and third interval (with a boundary at Feb. 2) coincides with the initiation of international air-travel bans with China (imposed by 34 countries and nation states by this date). To further explore the possible impact of the air-travel ban on the global spread of COVID-19, we inferred daily rates of global viral dispersal under a more granular interval-specific model (71*µ*4**Q**; Fig. 6, light blue). Our estimates of daily rates of global viral dispersal are significantly correlated with independent information on daily global air-travel volume (Fig. 6, dashed) over the interval from Jan. 31 (when the virus first achieved a cosmopolitan distribution; WHO, 2020) to the end of our study period (see Section S3.3 of the supplementary material for detailed descriptions of the correlation test and results).

**Fig. 6.**
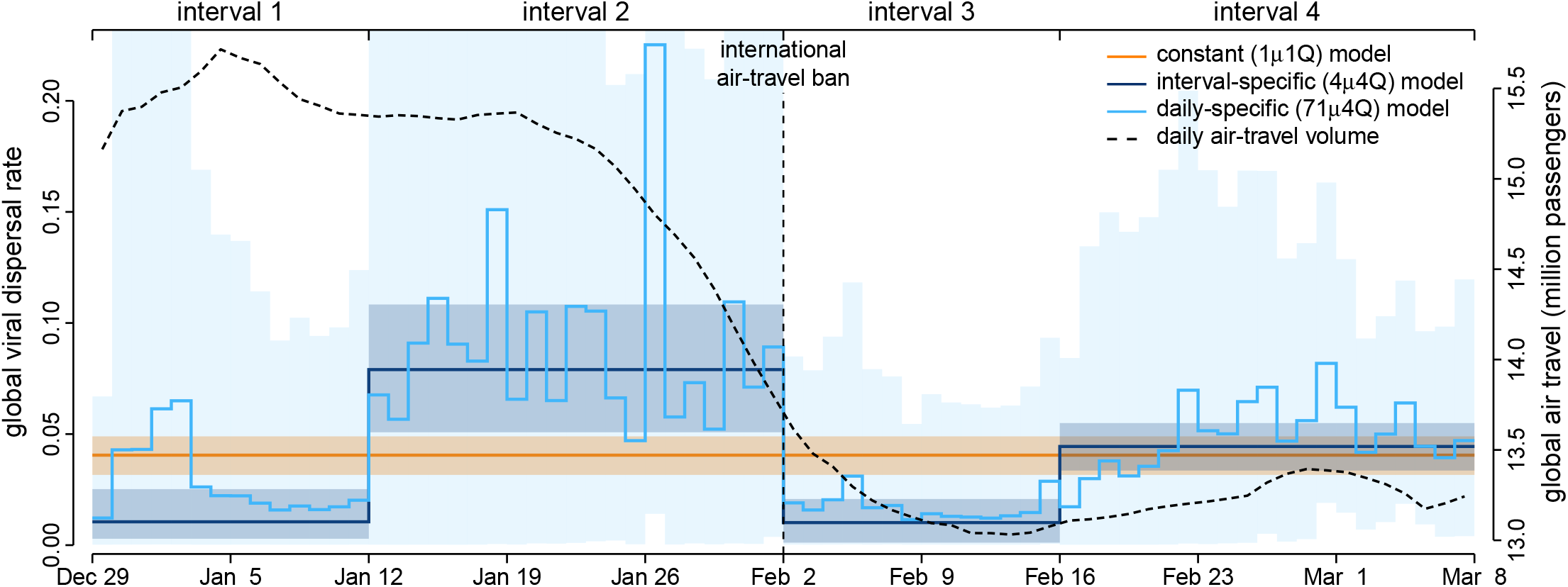
Patterns and correlates of variation in global viral dispersal rate early in the COVID-19 pandemic. The COVID-19 pandemic emerged in Wuhan, China, in late 2019, and established a global distribution by Mar. 8, 2020. Our phylodynamic analyses of this critical early phase of the pandemic provide estimates of the average rate of viral dispersal across all 23 study areas, *µ* (posterior mean [solid lines], 95% credible interval [shaded areas]). By assumption, the constant-rate (1*µ*1**Q**) model infers a static rate of global viral dispersal (orange). By contrast, the preferred interval-specific (4*µ*4**Q**) model reveals significant variation in the global viral dispersal rate (dark blue). Notably, the global viral dispersal rate decreases sharply on Feb. 2, which coincides with the onset of international air-travel bans with China. The efficacy of these air-travel restrictions is further corroborated by estimates of daily global viral dispersal rates (light blue)—inferred under a more granular, interval-specific (71*µ*4**Q**) model—that are significantly correlated with independent information on daily global air-travel volume (dashed line, obtained from FlightAware).

#### Variation in viral dispersal routes

In addition to revealing differences in the global viral dispersal rate, our interval-specific phylodynamic models allow us to explore how relative dispersal rates vary through time. Specifically, our analyses allow us to identify the dispersal routes by which the SARS-CoV-2 virus achieved a global distribution during the early phase of the COVID-19 pandemic. We focus on dispersal routes involving China both because it was the point of origin, and because it was the area against which travel bans were imposed. Inferences under the constant-rate (1*µ*1**Q**) and preferred (4*µ*4**Q**) phylodynamic models imply strongly contrasting viral dispersal dynamics (Fig. 7). In contrast to the invariant set of dispersal routes identified by the constant-rate model, the preferred interval-specific model reveals that the number and intensity of dispersal routes varied significantly over the four intervals, with a sharp decrease in the number of dispersal routes following the onset of air-travel bans on Feb. 2. Moreover, the constant-rate model infers one spurious dispersal route, while failing to identify six significant dispersal routes; the preferred model implies a more significant role for Hubei as a source of viral spread in the first and second intervals and reveals additional viral dispersal routes originating from China in the third and fourth intervals. The patterns of variation in dispersal routes among all 23 study areas are similar to—but more pronounced than—those involving China; *e*.*g*., where the constant-rate model infers a total of nine spurious dispersal routes, and the interval-specific model reveals a total of ten significant dispersal routes that were not detected by the constant-rate model (Figs. S16–S17).

**Fig. 7.**
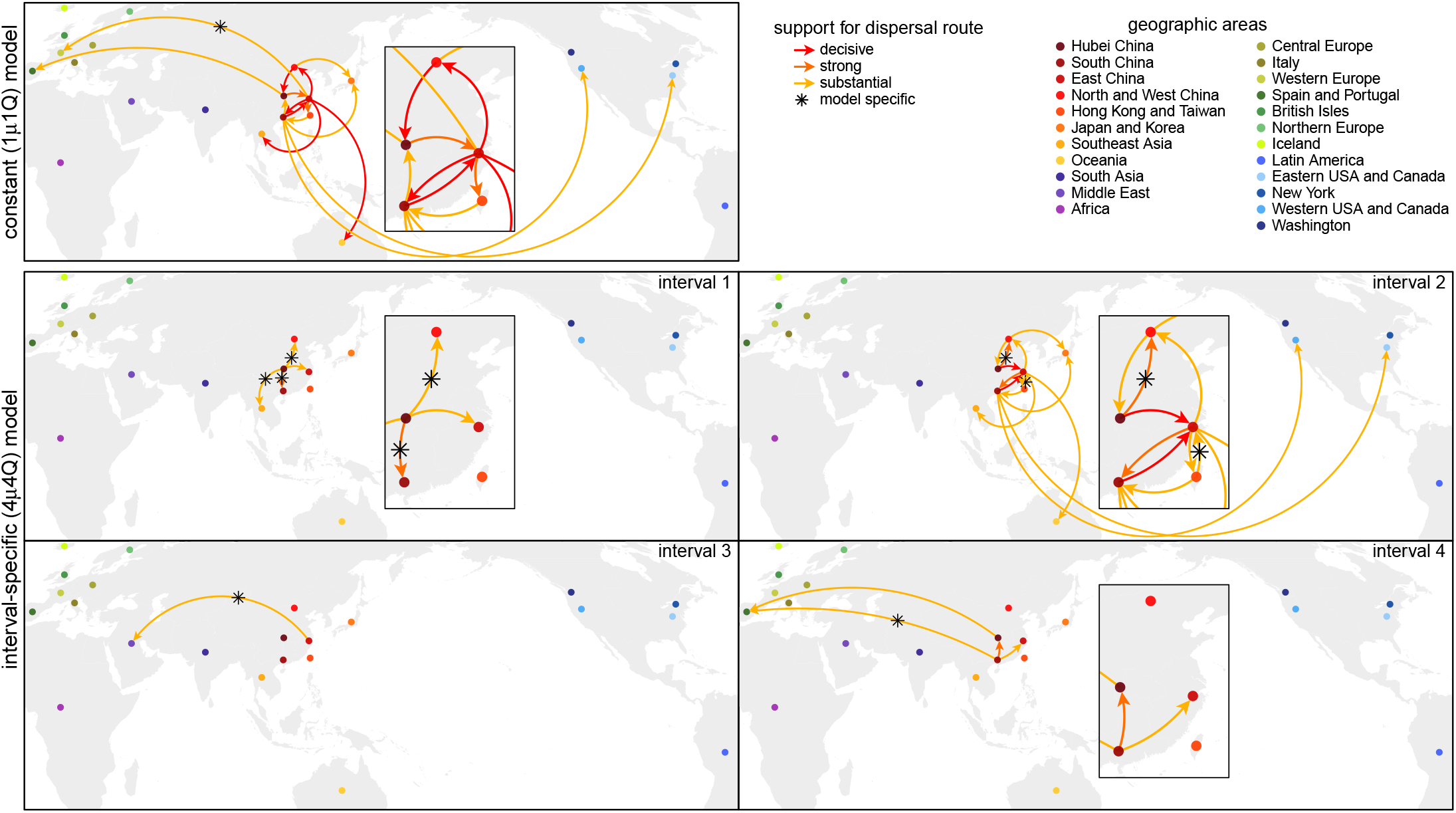
Variation in viral dispersal routes involving China during the early phase of the pandemic. Arrows indicate routes inferred to play a significant role in viral dispersal to/from China during the early phase of the COVID-19 pandemic; colors indicate the level of evidential support for each dispersal route (as 2 ln Bayes factors). We focus on dispersal routes involving China both because it was the point of origin, and because it was the area against which travel bans were imposed. The number, duration, and significance of dispersal routes inferred under the constant-rate (1*µ*1**Q**) model differ strongly from those inferred under the preferred (4*µ*4**Q**) interval-specific model. By assumption, the constant-rate (1*µ*1**Q**) model implies an invariant set of dispersal routes. By contrast, the preferred (4*µ*4**Q**) interval-specific model reveals that the number and intensity of dispersal routes varied over the four intervals. The first interval (Nov. 17–Jan. 12) is dominated by dispersal from Hubei to other areas in China, and the second interval (Jan. 12–Feb. 2) exhibits more widespread international dispersal originating from China. The third interval (Feb. 2–Feb. 16)—immediately following the onset of international air-travel bans with China—exhibits a sustained reduction in the number of dispersal routes. Note that the constant-rate model infers a spurious dispersal route from East China to West Europe. Conversely, the preferred interval-specific model reveals six significant dispersal routes (not detected under the constant-rate model) that imply a more significant role for Hubei as a source of viral spread in the first and second intervals, and also reveals additional dispersal routes emanating from China (to the Middle East in the third interval and to Spain/Portugal in the fourth interval).

#### Variation in the number of viral dispersal events

Our phylodynamic analyses also allow us to infer the number of SARS-CoV-2 dispersal events between areas during the early phase of the COVID-19 pandemic. Here, we focus on the number of viral dispersal events originating from China because it was the point of origin and primary source of SARS-CoV-2 spread early in the pandemic. The constant-rate (1*µ*1**Q**) and preferred (4*µ*4**Q**) phylodynamic models infer distinct trends in—and support different conclusions regarding the impact of mitigation measures on—the number of viral dispersal events out of China. The constant-rate model infers a gradual decrease in the number of dispersal events from late Jan. through mid-Feb. (Fig. 8, orange). By contrast, the preferred interval-specific model reveals a sharp decrease in the number of dispersal events on Feb. 2, which coincides with the onset of air-travel bans imposed against China (Fig. 8, blue). Moreover, the preferred phylodynamic model infers an uptick in the number of viral dispersal events on Feb. 17 (not detected by the constant-rate model), which coincides with the lifting of domestic travel restrictions within China (except for Hubei, where the travel restrictions were enforced through late Mar.).

**Fig. 8.**
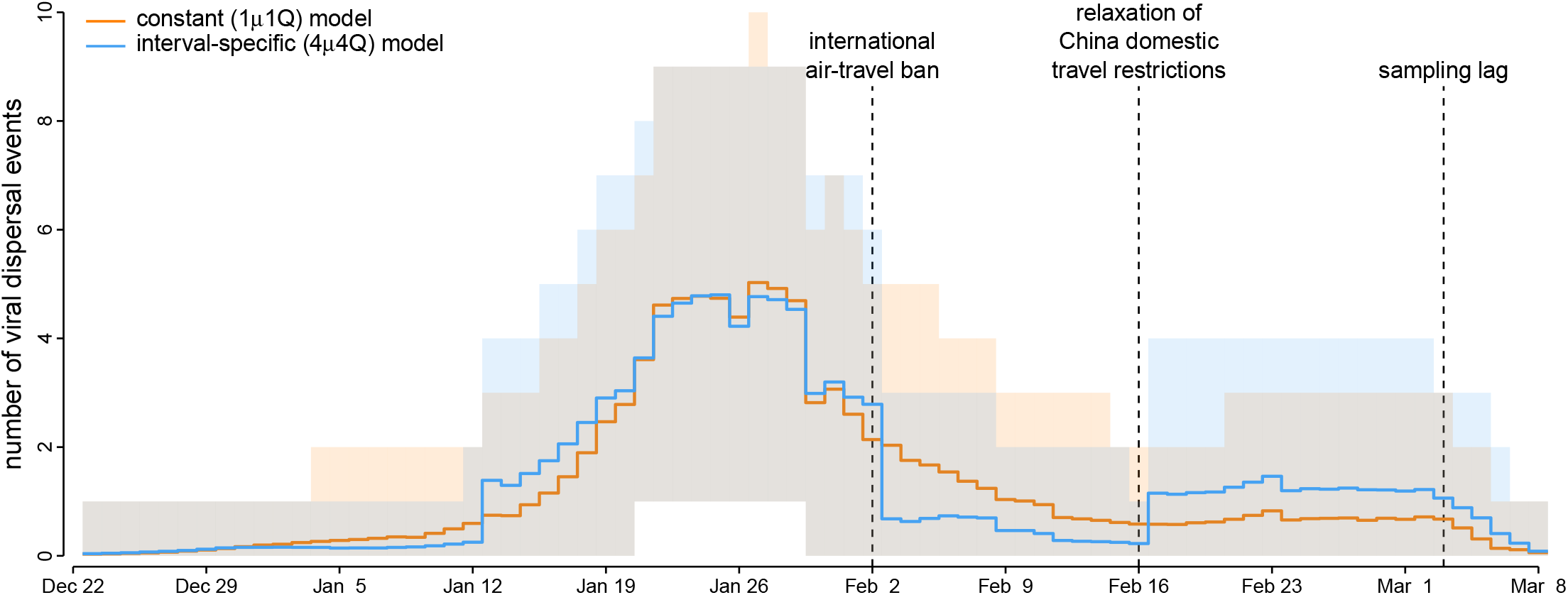
Variation in the number of viral dispersal events out of China early in the COVID-19 pandemic. Our phylogenetic analyses of SARS-CoV-2 genomes sampled during the early phase of the COVID-19 pandemic allow us to estimate the number of viral dispersal events from China to all other study areas (posterior mean [solid lines], 95% credible interval [shaded areas]). The constant-rate (1*µ*1**Q**) model implies that the number of viral dispersal events emanating from China remained relatively high following the onset of international air-travel bans on Feb. 2 (orange). By contrast, the preferred interval-specific (4*µ*4**Q**) model reveals that the number of viral dispersal events emanating from China decreased sharply on Feb. 2 (blue), which supports the efficacy of these international air-travel restrictions. The preferred model also infers an uptick in the number of viral dispersal events on Feb. 17 (not detected by the constant-rate model), which coincides with the relaxation of domestic travel restrictions in China. Note that sampling lag causes the number of dispersal events near the end of the sampling period to be underestimated.

## Discussion

Phylodynamic methods increasingly inform our understanding of the spatial and temporal dynamics of viral spread. The vast majority of discrete-geographic phylodynamic studies assume—despite direct (and compelling) evidence to the contrary—that disease outbreaks are intrinsically constant: *≈*98% of all such studies are based on the constant-rate models. These considerations have motivated previous extensions of phylodynamic models that allow *either* the average (Membrebe et al., 2019) *or* relative (Bielejec et al., 2014) dispersal rates to vary, and our development of more complex phylodynamic models that allow *both* the average and relative dispersal rates to vary independently over two or more pre-specified intervals. By accommodating ubiquitous temporal variation in the dynamics of disease outbreaks—and by allowing us to incorporate independent information regarding events that may impact viral dispersal— our new interval-specific phylodynamic models are more realistic (providing a better description of the processes that gives rise to empirical datasets), thereby enhancing the accuracy of our epidemiological inferences based on these models.

Our simulation study demonstrates that (in principle): (1) we are able to accurately identify when discrete-geographic phylodynamic models are correctly specified, overspecified, or underspecified (Fig. 4); (2) when the discrete-geographic phylodynamic model is correctly specified, we are able to reliably estimate parameters of these more complex interval-specific models (Fig. 3), and; (3) when the discrete-geographic phylodynamic model is underspecified, failure to accommodate interval-specific variation in the study data can bias parameter estimates and mislead inferences about viral dispersal history based on those biased estimates (Fig. 3).

Our empirical study of SARS-CoV-2 data from the early phase of the COVID-19 pandemic demonstrates that (in practice): (1) our interval-specific phylodynamic model (where *both* the global rate of viral dispersal *and* the relative rates of viral dispersal vary over four distinct intervals) significantly improves the relative and absolute fit to our study dataset compared to constant-rate phylodynamic models (Edwards et al., 2011; Lemey et al., 2009) and to phylodynamic models that allow *either* the average dispersal rate (Membrebe et al., 2019) *or* the relative dispersal rates (Bielejec et al., 2014) to vary over the same four intervals; (2) the preferred interval-specific phylodynamic model provides qualitatively different insights on key aspects of viral dynamics during the early phase of the pandemic—on global rates of viral dispersal (Fig. 6), viral dispersal routes (Fig. 7), and the number of viral dispersal events (Fig. 8)—compared to conventional estimates based on constant-rate (and underspecified) phylodynamic models, and; (3) inferences under the preferred interval-specific phylodynamic model support qualitatively different conclusions regarding the impact of mitigation measures to limit the spread of the COVID-19 pandemic; *e*.*g*., the variation in global viral dispersal rate, viral dispersal routes, and number of viral dispersal events revealed by the interval-specific model (but masked by the constant-rate model) collectively support the efficacy of the international air-travel bans in slowing the progression of the COVID-19 pandemic.

Our interval-specific models promise to enhance the accuracy of discrete-geographic phylodynamic inferences not only by virtue of their increased realism, but also by allowing us to incorporate additional information (related to events in the history of disease outbreaks) in our phylodynamic inferences. The ability to incorporate independent/external information is particularly valuable for phylodynamic inference—where many parameters must be estimated from datasets with limited information—which has also motivated the development of other innovative phylodynamic approaches for incorporating external information (Bielejec et al., 2016; Lemey et al., 2014). The potential benefit of harnessing external information is evident in our empirical study: our inference model—4*µ*4**Q**, with four intervals that we specified based on external evidence regarding events that might plausibly impact viral dispersal dynamics—is decisively preferred (2lnBF= 27.3) over a substantially more complex model, 5*µ*5**Q**^*\**^, with five *arbitrarily* specified (14-day) intervals.

Importantly, comparison of alternative interval-specific discrete-geographic phylodynamic models provides a powerful framework for testing hypotheses about the impact of various events (*i*.*e*., assessing the efficacy of mitigation measures) on viral dispersal dynamics. Our empirical study allows us, for example, to assess the impact of domestic mitigation measures imposed in the Hubei province of China. This simply involves comparing the relative fit of our data to two candidate phylodynamic models: 4*µ*4**Q** and 5*µ*5**Q**. The 5*µ*5**Q** model adds an interval (corresponding to the onset of the Hubei lockdown on Jan. 26) to the otherwise identical 4*µ*4**Q** model. In contrast to the international air-travel ban, this domestic mitigation measure does not appear to have significantly impacted global SARS-CoV-2 dispersal dynamics: the 5*µ*5**Q** model is decisively rejected when compared to the 4*µ*4**Q** model (2lnBF= *−*15.9).

We have focused on interval-specific models where each interval involves a change in both the average and relative dispersal rates. For example, the scenario depicted in Figure 1 involves two events that define three intervals, where both **Q** and *µ* are impacted by each event, such that the interval-specific parameters are (**Q**_1_,**Q**_2_,**Q**_3_) and (*µ*_1_,*µ*_2_,*µ*_3_). However, our interval-specific models also allow the average and relative dispersal rates to vary *independently* across intervals. For example, under an alternative scenario for Figure 1, the first event may have impacted both the relative and average dispersal rates, **Q** and *µ*, whereas the second event may have only changed the relative dispersal rates, **Q**; in this case, the interval-specific parameters would be (**Q**_1_,**Q**_2_,**Q**_3_) and (*µ*_1_,*µ*_2_,*µ*_2_). Allowing dispersal rates to vary independently enables these models to accommodate more complex patterns of variation in empirical datasets (and thereby improve estimates from these more realistic models), and also provides tremendous flexibility for testing hypotheses about the impact of various mitigation measures on *either* the relative and/or average rates of viral dispersal.

Nevertheless, this flexibility comes at a cost: interval-specific models are inherently more complex than their constant-rate counterparts, with many parameters that must be estimated from minimal data (*i*.*e*., the geographic location of each virus). Accordingly, careful model selection and validation is necessary to avoid specification of an over-parameterized model (although the Kullback–Leibler divergence between the posterior and the prior reveals similar amounts of information gain under the constant-rate and interval-specific models; see Section S3.3 of the supplementary material for detailed descriptions of the KL-divergence computation and results). Moreover, the space of discrete-geographic phylodynamic models expands rapidly as we increase the number of intervals. For a model with three intervals, for example, we can specify five allocations for the average dispersal rate parameter, *µ*—(*µ*_1_,*µ*_1_,*µ*_1_), (*µ*_1_,*µ*_1_,*µ*_2_), (*µ*_1_,*µ*_2_,*µ*_1_), (*µ*_1_,*µ*_2_,*µ*_2_), and (*µ*_1_,*µ*_2_,*µ*_3_)—and, similarly, five allocations for the relative dispersal rate parameter, **Q**: (**Q**_1_,**Q**_1_,**Q**_1_), (**Q**_1_,**Q**_1_,**Q**_2_), (**Q**_1_,**Q**_2_,**Q**_1_), (**Q**_1_,**Q**_2_,**Q**_2_), and (**Q**_1_,**Q**_2_,**Q**_3_). We can therefore specify 25 unique three-interval phylodynamic models (representing all combinations of the two parameter-allocation vectors), 225 unique four-interval models, 2704 unique five-interval models, 41209 unique six-interval models, etc. Accordingly, the effort required to identify the best interval-specific phylodynamic model quickly becomes prohibitive, particularly because this search requires that we estimate the marginal likelihood for each candidate model using computationally intensive methods (Baele et al., 2012; Xie et al., 2011). Nevertheless, our interval-specific models establish a foundation for developing more computationally efficient methods; *e*.*g*., we could pursue a finite-mixture approach (Kazmi and Rodrigue, 2019) that averages inferences of dispersal dynamics over the space of all possible interval-specific phylodynamic models with a given number of intervals. We are optimistic that—by increasing (and providing a means to assess) model realism, incorporating additional information, and providing a powerful and flexible means to test alternative models/hypotheses—our phylodynamic methods will greatly enhance our ability to understand the dynamics of viral spread, and thereby inform policies to mitigate the impact of disease outbreaks.

## Data Availability

All data produced are available online at this GitHub repository (https://github.com/jsigao/interval_specific_phylodynamic_models_supparchive) and this Dryad repository (https://datadryad.org/stash/share/vTbeDwLq2uSL9rL4NCe_Cocp2bY7BgWTI2tUgoNrLDA).

https://datadryad.org/stash/share/vTbeDwLq2uSL9rL4NCe_Cocp2bY7BgWTI2tUgoNrLDA

## Data and Code Availability

GISAID accession IDs of the SARS-CoV-2 sequences used in this study, as well as the flight-volume data (obtained from FlightAware, LLC) and intervention-measure data, are maintained in the GitHub repository (https://github.com/jsigao/interval_specific_phylodynamic_models_supparchive) and archived in the Dryad repository (https://datadryad.org/stash/share/vTbeDwLq2uSL9rL4NCe_Cocp2bY7BgWTI2tUgoNrLDA). Our repositories also contain BEAST XML scripts used to perform the phylodynamic analyses, R scripts used to perform simulations and post processing, and a modified version of the BEAST program used for some of the analyses in this study.

## Acknowledgments

This research was supported by the National Science Foundation grants DEB-0842181, DEB-0919529, DBI-1356737, and DEB-1457835 awarded to BRM, and the National Institutes of Health grant RO1GM123306-S awarded to BR.

## Footnotes

1 Note that the time slices that we define for summary statistics are distinct from the intervals specified in an interval-specific discrete-geographic phylodynamic model. The time slices are motivated to better assess the adequacy of a discrete-geographic phylodynamic model, whereas the intervals are motivated to accommodate variation in dispersal dynamics in the empirical data. Accordingly, we might use time-slice summary statistics to assess the adequacy of both constant-rate and interval-specific discrete-geographic phylodynamic models.

