## Supplementary Material for "New Phylogenetic Models Incorporating Interval-Specific Dispersal Dynamics Improve Inference of Disease Spread"

#### Contents

**S1 Extending Phylodynamic Methods** **S3**
- S1.1 Computing the transition-probability matrix when both average and relative dispersal rates vary across intervals . . . . . S3
- S1.2 Stochastic mapping when both average and relative dispersal rates vary across intervals . S5
- S1.3 Assessing adequacy of interval-specific discrete-geographic phylodynamic models . . . . S6
- S1.4 Running `BEAST` analyses under the interval-specific phylodynamic models . . . . . . . . . S8

**S2 Simulation Study** **S9**
- S2.1 Simulation design . . . . . S9
- S2.2 Generating simulated datasets . . . . . S9
- S2.3 Analyzing simulated datasets . . . . . S10
- S2.4 Summarizing results of the simulation analyses . . . . . S11
- S2.5 Results . . . . . S11
- S2.6 Assessing model fit to simulated datasets . . . . . S17

**S3 Empirical Application** **S20**
- S3.1 Overview . . . . . S20
- S3.2 Detailed Description of Data Acquisition and Curation . . . . . S25
- S3.3 Detailed Description of Phylodynamic Analyses . . . . . S30

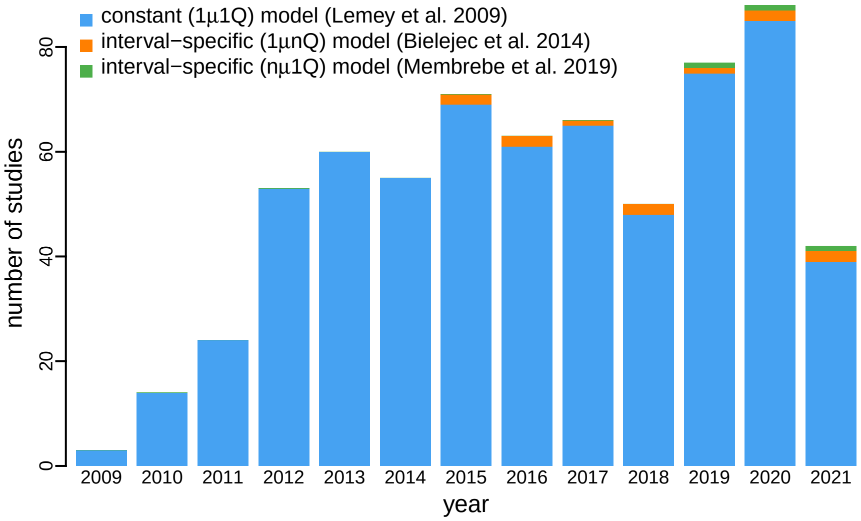


Figure S1: **Cited-reference search of empirical discrete-geographic phylodynamic studies.** The bar plot summarizes the number of published discrete-geographic studies (obtained from Google Scholar on June 30, 2021, searched among the publications that have cited Lemey et al. 2009, Bielejec et al. 2014, or Membrebe et al. 2019) that have inferred biogeographic history using the constant-rate phylodynamic model (blue; Lemey et al. 2009), an interval-specific model that allows only relative dispersal rates to vary (orange; Bielejec et al. 2014), or an interval-specific model that allows only the average dispersal rate to vary (green; Membrebe et al. 2019). The vast majority of these studies (651 of 666, 97.7%) are based on the constant-rate model, whereas only ∼2% of the studies used either of the interval-specific models.

### S1 Extending Phylodynamic Methods

We presented the theoretical base of the interval-specific discrete-geographic phylodynamic models in the main text; here we focus on the implementation extensions of `BEAST` that enable the inference and simulation under the model. Specifically, our implementation allows the transition-probability matrix, **P**, to be computed and the dispersal history to be simulated correctly along a branch when it spans multiple time intervals with different relative and/or absolute dispersal rates. We provide an executable `BEAST` program with our extensions in our GitHub and Dryad repositories.

#### S1.1 Computing the transition-probability matrix when both average and relative dispersal rates vary across intervals

Prior to our extension, `BEAST` computed the **P** matrix correctly (except for a couple of programming issues that we will describe below) along a branch spanning multiple relative-rate intervals *or* multiple average-rate intervals, but not *both*. Let a given branch of length $t$ time units span $m$ relative-rate intervals and $n$ average-rate intervals. The mean of the average dispersal rate over this branch is thus computed as:

$$\bar{\mu} = \frac{\sum_{p=1}^{n} \mu_p t_p}{t}, \tag{S1}$$

where $\mu_p$ and $t_p$ are the average dispersal rate and the time that the branch spent in average-rate interval $p$, respectively. `BEAST` previously computed the transition-probability matrix for each relative-rate interval $l$, $\mathbf{P}_l$, as:

$$\mathbf{P}_l = \exp(\mathbf{Q}_l \bar{\mu} t_l), \tag{S2}$$

where $\mathbf{Q}_l$ is the instantaneous-rate matrix in relative-rate interval $l$ and $t_l$ represents the time that the branch spent in relative-rate interval $l$. In other words, this way of computing $P_l$ effectively assumed that the average dispersal rate in each relative-rate interval was identical across all relative-rate intervals, which is not correct when the branch spans *both* multiple relative-rate intervals *and* multiple average-rate intervals.

To correctly compute the **P** matrix, we modified `BEAST` source code to set $t_p$ to the time that *relative-rate interval l* (instead of the *branch*) spent in average-rate interval $p$, rather than the time that the *branch* spent in average-rate interval $p$. Implementation details and source-code edits are available in this pull request to the source-code repository of `BEAST`. Putting all the steps together, the transition-probability matrix is now computed as:

$$\mathbf{P} = \prod_{l=1}^{m} \exp[\mathbf{Q}_l (\sum_{p=1}^{n_l} \mu_p t_p)], \tag{S3}$$

where $n_l$ is the number of average-rate intervals spanned by interval $l$, $\mu_p$ is the dispersal rate in average-rate interval $p$, and $t_p$ is the time that relative-rate interval $l$ spent in average-rate interval $p$.

##### Other relevant programming limitations that prohibited inferences under the interval-specific models

In addition, we identified and fixed two programming bugs that hinder correct inferences under interval-specific phylodynamic models: the first one matters when the relative dispersal rates vary across intervals, while the second one can be problematic even when the phylodynamic model is constant, but may be exacerbated when both average and relative dispersal rates vary across intervals.

**Programming bug 1: P-matrix ordering along a branch**

The transition-probability matrix for the entire branch is computed as the matrix product of interval-specific transition-probability matrices:

$$\mathbf{P} = \prod_{l=1}^{m} \mathbf{P}_l, \tag{S4}$$

Since matrix multiplication is not commutative, these **P** matrices should be ordered from the parent node to the child node of the branch, *i.e.*, forward in time, as shown in Eq. 7 of Bielejec et al. (2014). However, the vector of **Q**-matrix indices along a branch returned by the `getBranchModelMapping` function of the interval-specific relative dispersal rates (`EpochalBranchModel`) class was ordered from the child node to the parent node; there was no reversal of the order prior to or during the computation of **P** matrices, resulting in an incorrect transition-probability matrix computed for the entire branch. We fixed this issue by reversing the order of **Q** matrices that will be returned by the `getBranchModelMapping` function. Implementation details and source-code edits of this fix are available in this pull request to the source-code repository of `BEAST`.

**Programming bug 2: rescaling an asymmetric Q matrix**

By convention, we rescale the **Q** matrix such that the average rate of dispersal between all areas is $\mu$, which is computed as:

$$\mu = -\sum_{i=1}^{k} \pi_i q_{ii}, \tag{S5}$$

where $k$ is the number of discrete areas, $\pi_i$ is the stationary frequency of area $i$, and $q_{ii}$—the diagonal element of row $i$ of **Q**—is the negative of the total rate of leaving area $i$ (*i.e.*, $q_{ii} = -\sum_{j \neq i} q_{ij}$). (Note that the implicit assumption here is that the **Q** matrix is irreducible, which guarantees the existence and uniqueness of the stationary distribution, $\pi$.) After rescaling, **Q** becomes an instantaneous-rate matrix of relative dispersal rates whose average rate of dispersal is one; thus $\mu$ represents the average dispersal rate (among areas) in units of expected number of dispersal events per unit time. As the **Q** matrix is now constrained, $\mu$ is a free parameter of the model.

Therefore, to rescale **Q**, we need to know $\pi$. $\pi$ can be determined from **Q** by solving:

$$\pi \mathbf{Q} = 0. \tag{S6}$$

This way of determining $\pi$ is unnecessary when **Q** is symmetric (where the rate of dispersal from area $i$ to area $j$ is identical to the rate of dispersal from area $j$ to area $i$) or time reversible (*e.g.*, under the GTR substitution model; Tavaré 1986). Depending on the specified model, `BEAST` thus treats $\pi$ either as a constant uniform vector (when **Q** is symmetric) or a model parameter that will be used to construct **Q** and directly sampled in the MCMC (when **Q** is not symmetric but time reversible).

Conversely, when **Q** is asymmetric (Edwards et al. 2011)—where the rate of dispersal from area $i$ to area $j$ is different from the rate of dispersal from area $j$ to area $i$ (*i.e.*, $q_{ij}$ is different from $q_{ji}$)—$\pi$ is necessarily nonuniform nor an explicit model parameter, so it needs to be computed using (S6). However, previously `BEAST` did not allow this option; $\pi$ had to be explicitly specified as either a constant vector or model parameter in the XML file, and this specified $\pi$ would be used to rescale **Q**. As a result, the average rate of disperal for the **Q**-matrix would depart from 1, which risks conflating relative-rate matrix variation from overall dispersal-rate variation. We modified `BEAST` source code to allow $\pi$ to be provided optionally, and to rescale an asymmetric **Q** using a stationary distribution computed from (S6) through LU decomposition by adding a `computeStationaryDistribution` function to the asymmetric substitution model (`ComplexSubstitutionModel`) class. Implementation details and source-code edits of this fix are available in this pull request to the source-code repository of `BEAST`.

#### S1.2 Stochastic mapping when both average and relative dispersal rates vary across intervals

Stochastic mapping, initially proposed by Nielsen (2002; see also Huelsenbeck et al. 2003; Bollback 2006), is commonly used to sample dispersal histories over branches of a phylogeny conditioned on the observed tip geographic areas. `BEAST` implements the endpoint-conditioned uniformization stochastic-mapping algorithm (Hobolth and Stone 2009) to simulate full dispersal histories over the phylogeny, and a simulation-free algorithm (Minin and Suchard 2008a,b) to compute the expected number of dispersal events ('Markov jumps') and the expected time spent in each geographic area ('Markov rewards'). Here we focus on inferring the full dispersal history using the simulation-based stochastic-mapping algorithm.

A full dispersal history is sampled every certain number of generations (specified in the XML file) during an MCMC. At a sampling generation, the geographic state at each internal node of the phylogeny is first simulated using the ancestral-state-reconstruction algorithm (Yang et al. 1995; Huelsenbeck and Bollback 2001; Pagel et al. 2004) based on parameter values sampled at that generation. (Note that the implementation extensions and programming issues described above in Section S1.1 that underlie the computation of transition-probability matrix under the interval-specific model also affect the ancestral-state reconstruction as the probability of transitioning from the start state to the end state of a branch is used to sample the end state conditioning on the start state, propagating the sampled root state to the tips of the phylogeny.) The stochastic-mapping algorithm is then responsible for simulating the dispersal history over each branch of the phylogeny, conditioning on the start and end states of each branch.

Let a given branch of length $t$ time units start at time $T_0$ with state $i$ and end at time $T_m$ with state $k$. Further, let the dispersal process change (either by changing the average or relative dispersal rates) $m-1$ times on the branch at times $\{T_1, \dots, T_{m-1}\}$, resulting in $m$ intervals. For interval $l$, denote the average dispersal rate as $\mu_l$, the instantaneous-rate matrix as $\mathbf{Q}_l$, and the duration as $t_l$. Prior to our extension, `BEAST` performed stochastic mapping along the branch using the same routine regardless whether the model was constant or interval-specific; *i.e.*, it used a single $\mu$ and a single $\mathbf{Q}$ to perform the simulation. Specifically, the single $\mu$ was assumed to be the average of the average dispersal rates spanned by the branch, computed as:

$$\bar{\mu} = \frac{\sum_{l=1}^{m} \mu_l t_l}{t}, \tag{S7}$$

and the single $\mathbf{Q}$ was assumed to be $\mathbf{Q}_m$, the instantaneous-rate matrix of the last interval spanned by the branch.

We resolved this issue by adding a new routine to the `MarkovJumpsBeagleTreeLikelihood` class of `BEAST`, which simulates a dispersal history along the branch following a two-step procedure: (1) first, we sample the state (area) at each of the $m-1$ time points along the branch, and; (2) then we simulate the history between each time point, conditional on the states sampled in the first step; the second step is based on the fact that both the average and relative dispersal rates spanned by interval $l$ are constant. To sample states at each time point, we first compute a transition-probability matrix for each interval:

$$\mathbf{P}_l = \exp\left(\mathbf{Q}_l \mu_l t_l\right). \tag{S8}$$

We then calculate the probability of state $j$ at the first time point, $T_1$, given that the branch begins in state $i$ and ends in state $k$, as:

$$\text{conditional probability of } j = \frac{\text{joint probability of } i, j, k}{\text{marginal probability of } i \text{ to } k \text{ transition}}$$

$$P(j \mid i, k) \propto \mathbf{P}_{ij,1} \times \left[\prod_{l=2}^{m} \mathbf{P}_l\right]_{jk}, \tag{S9}$$

where the first term is the probability of transitioning from state $i$ (the state at the beginning of the branch) to state $j$ at the first time point, and the second term is the probability of transitioning from state $j$ to state $k$ (the state at the end of the branch) over the remaining time intervals. We compute this for each state $j$, and sample the state in proportion to these probabilities. We then repeat this process for each remaining time point, recursively conditioning on the state sampled at the previous time point and the state at the end of the branch.

Once the state at each time point is sampled, we invoke the existing endpoint-conditioned uniformization stochastic-mapping routine (Hobolth and Stone 2009) to simulate the history in each interval conditional on its start and end states. The resulting simulated histories across intervals along the branch are then pasted together so that the history output format leaves unchanged.

(Note that in principle the simulation-free stochastic-mapping algorithm implemented in `BEAST` could be modified very similarly to work under the interval-specific model, but we did not make such changes as it was unclear to us in the first place what would be the issues with the current implementation of the algorithm when $\mu$ and/or $\mathbf{Q}$ vary across intervals.)

Implementation details and source-code edits of this fix are available in this pull request to the source-code repository of `BEAST`. We also implemented this stochastic-mapping function in `R`, which uses the model parameters and ancestral state of each internal node sampled during `BEAST` MCMC as the input. These two independent implementations provide a means to validate our methods (we include both implementations in our GitHub and Dryad repositories). These two independent implementations produce effectively identical estimates of the number of viral dispersal events (Fig. S2).

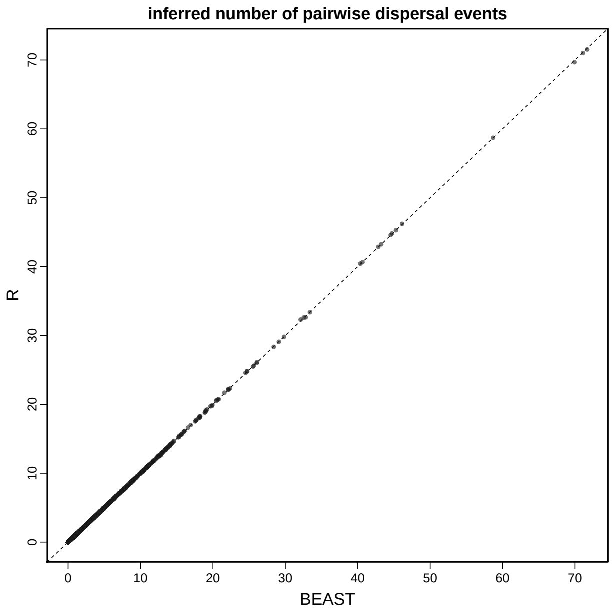


Figure S2: **Comparison of the estimated number of pairwise dispersal events in two independent implementations.** Each dot represents the mean estimate of the number of dispersal events in a given time interval between a given pair of areas. Independent implementations in `BEAST` and `R` produce effectively identical estimates.

#### S1.3 Assessing adequacy of interval-specific discrete-geographic phylodynamic models

We use posterior-predictive simulation (Gelman et al. 1996; Bollback 2002) to assess the adequacy of our interval-specific phylodynamic models. Posterior-predictive simulation requires: (1) the ability to simulate geographic datasets under the interval-specific models for a given set of parameter values, and; (2) summary statistics that allow us to compare the resulting simulated datasets to the observed dataset. We describe each of these components below.

#### Simulating under interval-specific discrete-geographic phylodynamic models

We draw $m$ random samples from the joint posterior distribution of the model; each sample $i$ consists of a fully specified phylodynamic model, $\theta_i = \{\Psi_i, \mathbf{Q}_i, \mu_i\}$. For each sample, we simulate a new geographic dataset on the sampled tree, $\Psi_i$, given the sampled parameters of the geographic model, $\{\mathbf{Q}_i, \mu_i\}$; we label the newly simulated dataset $G_i^{\text{sim}}$.

Under a constant-rate phylodynamic model, we simulate full dispersal histories forward in time over a tree using the `sim.history()` function in the `R` package `phytools` (Revell 2012). We implemented an extension of the `sim.history()` function to simulate dispersal histories under interval-specific phylodynamic models. These functions allow us to perform posterior-predictive simulation to assess the adequacy of both the constant-rate and the interval-specific phylodynamic models. We provide these `R` scripts in our GitHub and Dryad repositories.

#### Summary statistics

We define a summary statistic, which we generically denote $T(G \mid \theta_i)$, where $G$ is either the simulated or observed dataset. (We note that the dependence of the statistic on $\theta_i$—while often suppressed or ignored in phylogenetic applications of posterior-predictive simulation—is consistent with posterior-predictive discrepancy analysis, as described by Gelman et al. 1996.) For each simulated dataset, we compute a discrepancy statistic,

$$D_i = T(G_i^{\text{sim}} \mid \theta_i) - T(G^{\text{obs}} \mid \theta_i),$$

where $G^{\text{obs}}$ is the observed geographic dataset and $G^{\text{sim}}$ is a simulated dataset.

If the inference model provides an adequate description of the true data-generating process, the posterior-predictive distribution of $D$ should contain zero with high probability. Accordingly, for the $m$ predictive datasets for a given model and dataset combination, we calculate the posterior-predictive $p$ value as:

$$P = \frac{1}{m} \sum_{i=1}^{m} D_i \geq 0.$$

Values between 0.025 and 0.975 indicate that the model is adequate and cannot be rejected (*i.e.*, zero falls within the 95% posterior-predictive interval).

We developed two summary statistics to assess the adequacy of interval-specific phylodynamic models: (1) the *parsimony statistic*, and; (2) the *tipwise-multinomial statistic*. The parsimony statistic is calculated as the difference in the parsimony score for the observed areas and the simulated areas across the tips of the tree (where the parsimony score is the minimum number of dispersal events required to explain the distribution of areas across the tips of a tree). We compute parsimony scores using the `parsimony()` function in the `R` package, `phangorn` (Schliep 2010). The tipwise-multinomial statistic is inspired by the multinomial statistic that was first proposed by Goldman (1993) and later used by Bollback (2002) to assess the adequacy (absolute fit) of substitution models to sequence alignments. Our tipwise statistic treats the set of states (areas) across the tips of the tree as an outcome of a multinomial trial. Specifically, we calculate the tipwise-multinomial statistic as the difference in the multinomial probabilities for the observed the set of areas and the simulated the set of areas across the tips of the tree. We calculate each multinomial probability as:

$$T(G \mid \theta_i) = \sum_{i=1}^{k} n_i \ln(n_i/n),$$

where $n$ is the number of tips in the tree, and $n_i$ is the number of tips that occur area $i$. (Note that this statistic is also similar to the entropy statistic used to assess genetic variability along sequences; Shannon 1948; Schneider et al. 1986.)

###### Time-slice summary statistics

We could use our summary statistics to assess the ability of a discrete-geographic phylodynamic model to describe the entire history of dispersal; *i.e.*, to assess whether the model adequately describes the process that generated the data over the entire phylogeny. Of potential concern, however, is the sufficiency of the summary statistics to detect variation in the data-generating process over time. For example, it is conceivable that a discrete-geographic phylodynamic model may be capable of simulating datasets with parsimony scores that are very similar to those for the observed dataset—implying that the model provides an adequate description of the process that generated the entire geographic history—but the underlying dispersal events simulated under this model may nevertheless occur at inappropriate times.

To assess the ability of discrete-geographic phylodynamic models to describe the temporal distribution of dispersal events, we extend the parsimony and tipwise-multinomial summary statistics to assess time slices of the geographic history[1]. We calculate these summary statistics for $k$ pre-specified time slices, resulting in $k$ parsimony statistics and $k$ tipwise-multinomial statistics for each simulated dataset. We compute the time-slice variant of the parsimony statistic as follows: (1) we first infer the most-parsimonious dispersal history (*i.e.*, the minimum number of dispersal events) for a given simulated dataset and the observed dataset using the `ancestral.pars()` function in the `R` package, `phangorn` (Schliep 2010); (2) we then assign each inferred dispersal event to one of the $k$ time slices based on the time span of the branch along which the dispersal event was inferred (when a dispersal event is inferred to occur along a branch that spans two or more time slices, we locate the event uniformly along the branch, and then assign it to the corresponding slice), and finally; (3) we compute the difference in the number of dispersal events between the simulated and observed dataset for each time slice. We compute the time-slice variant of the tipwise-multinomial statistic in a similar manner; *i.e.*, we first find the set of tips in each time slice, and then compute the tipwise-multinomial statistic for that time slice (as described above) for the corresponding set of tips. Further details regarding the computation of these summary statistics are available in an `R` script, `posterior_predictive_teststatistics_functions.R`, included in our GitHub and Dryad repositories.

#### S1.4 Running `BEAST` analyses under the interval-specific phylodynamic models

To aid the application of our newly developed interval-specific phylodynamic models with `BEAST`, we provide a hands-on tutorial—that describes how to specify the model in an XML file—in our GitHub and Dryad repositories.

[1]Note that the time slices that we define for summary statistics are distinct from the intervals specified in an interval-specific discrete-geographic phylodynamic model. The former are motivated to better assess the adequacy of a discrete-geographic phylodynamic model, the latter are motivated to accommodate variation in dispersal dynamics in the empirical data. Accordingly, we might use time-slice summary statistics to assess the adequacy of both constant-rate or interval-specific discrete-geographic phylodynamic models.

### S2 Simulation Study

#### S2.1 Simulation design

We performed a simulation study to explore the statistical behavior of our interval-specific phylodynamic models. Specifically, we sought to assess: (1) our ability to perform reliable inference under interval-specific models; (2) the impact of model misspecification, and; (3) our ability to identify the correct model. To provide a meaningful evaluation of the statistical behavior of a method, it is critical for a simulation study to explore realistic parameter space (*i.e.*, to subject the method to simulated datasets that are similar to those it will actually encounter in empirical analyses). Accordingly, we focused our simulation study on simulated datasets that resemble our empirical SARS-CoV-2 reduced dataset. That is, our simulation study explored a region of parameter space that is centered on the joint posterior probability distribution of discrete-geographic phylodynamic model parameters estimated from our empirical analyses. To that end, we first analyzed our empirical dataset under each of two models, $1\mu1\mathbf{Q}$ and $2\mu2\mathbf{Q}$, and then centered the parameter values of our simulation on the resulting posterior median estimates of the corresponding parameters ($\mu$ and $\mathbf{Q}$). Specifically, we used these empirically based parameter values to simulate 200 geographic datasets under each of two models, $1\mu1\mathbf{Q}$ and $2\mu2\mathbf{Q}$. Finally, we performed separate analyses of each simulated dataset under each of the two models, resulting in four true:inference model combinations ($1\mu1\mathbf{Q}$:$1\mu1\mathbf{Q}$, $2\mu2\mathbf{Q}$:$2\mu2\mathbf{Q}$, $1\mu1\mathbf{Q}$:$2\mu2\mathbf{Q}$, and $2\mu2\mathbf{Q}$:$1\mu1\mathbf{Q}$). Below, we provide additional details of how we generated simulated datasets, analyzed these simulated datasets, and summarized results from our analyses of the simulated datasets.

#### S2.2 Generating simulated datasets

##### Model specification for empirical analyses

We performed analyses of our reduced SARS-CoV-2 dataset under a constant-rate ($1\mu1\mathbf{Q}$) and an interval-specific discrete-geographic phylodynamic ($2\mu2\mathbf{Q}$) model. Our reduced SARS-CoV-2 dataset has 1271 sequences. To reduce the computational burden of our simulation study we: (1) aggregated our 23 study areas into three more coarsely defined areas: China, North America, and the rest of the world, and; (2) conditioned our analyses on the MCC summary phylogeny inferred from our reduced SARS-CoV-2 dataset (described in Section S3.3). We specified the single boundary for the $2\mu2\mathbf{Q}$ model at February 2 (corresponding to the onset of international air-travel bans against China). For both the constant-rate ($1\mu1\mathbf{Q}$) and interval-specific ($2\mu2\mathbf{Q}$) models, we used an asymmetric $\mathbf{Q}$ matrix (Edwards et al. 2011) that allows the relative rate of dispersal from area $i$ to area $j$ to be different from the relative rate of dispersal from area $j$ to area $i$. We specified diffuse priors on the parameters (for the average dispersal rate, $\mu$, and relative dispersal rates, $\mathbf{Q}$) for each phylodynamic model (Table S1).

We also specified a root-frequency vector, $\omega$, which represents the prior probability that the tree begins in each of the geographic areas. For models with a single $\mathbf{Q}$, it is possible to use the stationary frequency implied by the $\mathbf{Q}$ as this root-frequency vector. However, interval-specific models do not have a global stationary frequency (*i.e.*, each interval has a separate stationary frequency); in this case, it is conventional to treat $\omega$ as a free parameter and estimate it from the data. To be as consistent as possible between the constant-rate and interval-specific models, we specified $\omega$ as a free parameter for both models.

Table S1: **Priors used in analyses of the reduced SARS-CoV-2 dataset.**

| Parameter | Description | Prior |
|---|---|---|
| $\mu_l$ | Average dispersal rate in interval $l$ | $\text{Exp}(1/\lambda)$; $\lambda \sim \Gamma(0.5, 0.5)$ |
| $r_{ij,l}$ | Relative dispersal rate from $i$ to $j$ in interval $l$ | $\Gamma(1, 1)$ |
| $\boldsymbol{\omega}$ | Root frequencies | $\text{Dir}(1, 1, 1)$ |

##### Parameter estimation

For each of model ($1\mu1\mathbf{Q}$ and $2\mu2\mathbf{Q}$), we inferred the joint posterior distribution of parameters from our empirical dataset by running four independent MCMC simulations using our modified version of `BEAST` (see Section S1) with the `BEAGLE` library (compiled from the 'hmc-clock' branch, commit 'dd36bf5'; Ayres et al. 2019). We ran each replicate MCMC simulation for 700000–800000 generations, sampling every 500–1000 generations. We discarded the initial 100000–250000 generations (as burn-in) from each replicate MCMC simulation, and then combined the remaining posterior samples from all replicate simulations using `LogCombiner` version 1.10.5. (The number of generations, sampling frequency, and the length of the burn-in are presented as ranges here and below because we deliberately ran the analyses under more complex models longer and sampled less frequently.) We then assessed MCMC performance for the resulting composite posterior sample by inspecting the log files using `Tracer` (Rambaut et al. 2018) version 1.7.1, and using the `coda` package (Plummer et al. 2006) in `R` (R Core Team 2020). Specifically, we ensured that the computed ESS values for all continuous parameters were $\gg$2000. Details of these analyses are available in the XML scripts included in our GitHub and Dryad repositories.

##### Simulating geographic datasets using the inferred parameter values

We simulated a total of 200 geographic datasets under each of two models: the constant-rate ($1\mu1\mathbf{Q}$) and interval-specific ($2\mu2\mathbf{Q}$) phylodynamic models, using our forward-in-time simulator (the simulator `R` script is included in our GitHub and Dryad repositories). We simulated these geographic datasets over the MCC summary tree inferred from our reduced SARS-CoV-2 dataset (see Section S3.3), with parameter values (for $\mu$ and $\mathbf{Q}$) of the simulating models ($1\mu1\mathbf{Q}$ and $2\mu2\mathbf{Q}$) set to the corresponding posterior median estimates from our empirical analyses (described in Section S2.2). Specifically, we used the following parameter values for constant-rate ($1\mu1\mathbf{Q}$) model:

$$\mu = 0.0320, \ \mathbf{Q} = \begin{pmatrix} - & 1.4018 & 0.1740 \\ 0.0190 & - & 0.7366 \\ 0.1104 & 1.2983 & - \end{pmatrix},$$

and the following parameter values for the interval-specific ($2\mu2\mathbf{Q}$) model:

$$\mu_1 = 0.0242, \ \mathbf{Q}_1 = \begin{pmatrix} - & 1.4214 & 1.2043 \\ 0.0107 & - & 0.7309 \\ 0.0654 & 1.3779 & - \end{pmatrix}; \quad \mu_2 = 0.0602, \ \mathbf{Q}_2 = \begin{pmatrix} - & 0.6974 & 0.0814 \\ 1.0008 & - & 0.2428 \\ 0.8077 & 0.5744 & - \end{pmatrix}.$$

Values of the diagonal elements are specified in the usual manner (*i.e.*, set equal to the negative sum of the off-diagonal elements in the corresponding row).

#### S2.3 Analyzing simulated datasets

##### Model specification

For each simulated dataset, we inferred the joint posterior distribution under each of the two models, $1\mu1\mathbf{Q}$ and $2\mu2\mathbf{Q}$, using the same priors (listed in Table S1) specified in the empirical analyses that generated parameter values used to simulate the datasets.

##### Parameter estimation

For each inference model, $1\mu1\mathbf{Q}$ and $2\mu2\mathbf{Q}$, we estimated the joint posterior distribution for each simulated dataset by running two to four independent MCMC simulations using our modified version `BEAST` (see Section S1) with the `BEAGLE` library (compiled from the 'hmc-clock' branch, commit 'dd36bf5';

Ayres et al. 2019). We ran each replicate MCMC simulation for 300000–800000 generations, sampling every 500–1000 generations. When a sample was drawn, we performed stochastic mapping using the endpoint-conditioned uniformization algorithm (Hobolth and Stone 2009) and our modified algorithm for interval-specific discrete-geographic phylodynamic models (see Section S1) implemented in `BEAST` to simulate a dispersal history over the MCC phylogeny. We discarded the initial 50000–100000 generations (as burn-in) from each replicate MCMC simulation, and then combined the remaining posterior samples from all replicate simulations using `LogCombiner` version 1.10.5. We then assessed MCMC performance for the resulting composite posterior sample by inspecting the log files using `Tracer` (Rambaut et al. 2018) version 1.7.1, and using the `coda` package (Plummer et al. 2006) in `R` (R Core Team 2020). Specifically, we ensured that the computed ESS values for all continuous parameters were $\gg$500. Details of these analyses are available in the XML scripts included in our GitHub and Dryad repositories.

#### S2.4 Summarizing results of the simulation analyses

For each of the four true:inference model combinations, we first computed the coverage probability (the frequency with which the true value was contained in the $X$% posterior credible interval) as a function of the size ($X$) of the credible interval for all model parameters ($\mu$ and $\mathbf{Q}$) and for the pairwise and total number of dispersal events. For each true:inference model combination, we also summarized the absolute error (estimated minus true values) for the model parameters ($\mu$ and $\mathbf{Q}$) and the pairwise and total number of dispersal events.

When the inference model is correctly specified (*i.e.*, scenarios $1\mu1\mathbf{Q}$:$1\mu1\mathbf{Q}$, and $2\mu2\mathbf{Q}$:$2\mu2\mathbf{Q}$), there is a one-to-one correspondence between the true:inference model parameters, which allows us to simply compare the true:estimated values for each parameter. By contrast, when the inference model is misspecified (*i.e.*, scenarios $1\mu1\mathbf{Q}$:$2\mu2\mathbf{Q}$, and $2\mu2\mathbf{Q}$:$1\mu1\mathbf{Q}$), there is a lack of direct correspondence between the true:inference model parameters. When the inference model is *overspecified* (*i.e.*, where $2\mu2\mathbf{Q}$:$1\mu1\mathbf{Q}$), we compared the inferred parameter value for each interval to the true, time-constant value (*e.g.*, we compared interval-specific estimates of $\mu_1$ and $\mu_2$ to the time-constant true value, $\mu$). Conversely, when the inference model is *underspecified* (*i.e.*, where $2\mu2\mathbf{Q}$:$1\mu1\mathbf{Q}$), we compared estimates of the time-constant parameter values to each true, interval-specific parameter values (*e.g.*, we compared time-constant estimates of $\mu$ to both interval-specific true values, $\mu_1$ and $\mu_2$).

#### S2.5 Results

When the inference model is correctly specified (*i.e.*, where both the true and inference models include [or exclude] interval-specific parameters, $2\mu2\mathbf{Q}$:$2\mu2\mathbf{Q}$ and $1\mu1\mathbf{Q}$:$1\mu1\mathbf{Q}$), our simulation study demonstrates that estimates under interval-specific models are as reliable as those under constant-rate models (Fig. 3 and Figs. S3–S7, green and blue). Moreover, when the inference model is overspecified (*i.e.*, it includes interval-specific parameters not included in the true model) inferences are comparable to those under correctly specified models (Fig. 3 and Figs. S3–S7, purple). However, when the inference model is underspecified (*i.e.*, it excludes interval-specific parameters of the true model) inferences are severely biased (Fig. 3 and Figs. S3–S7, orange).

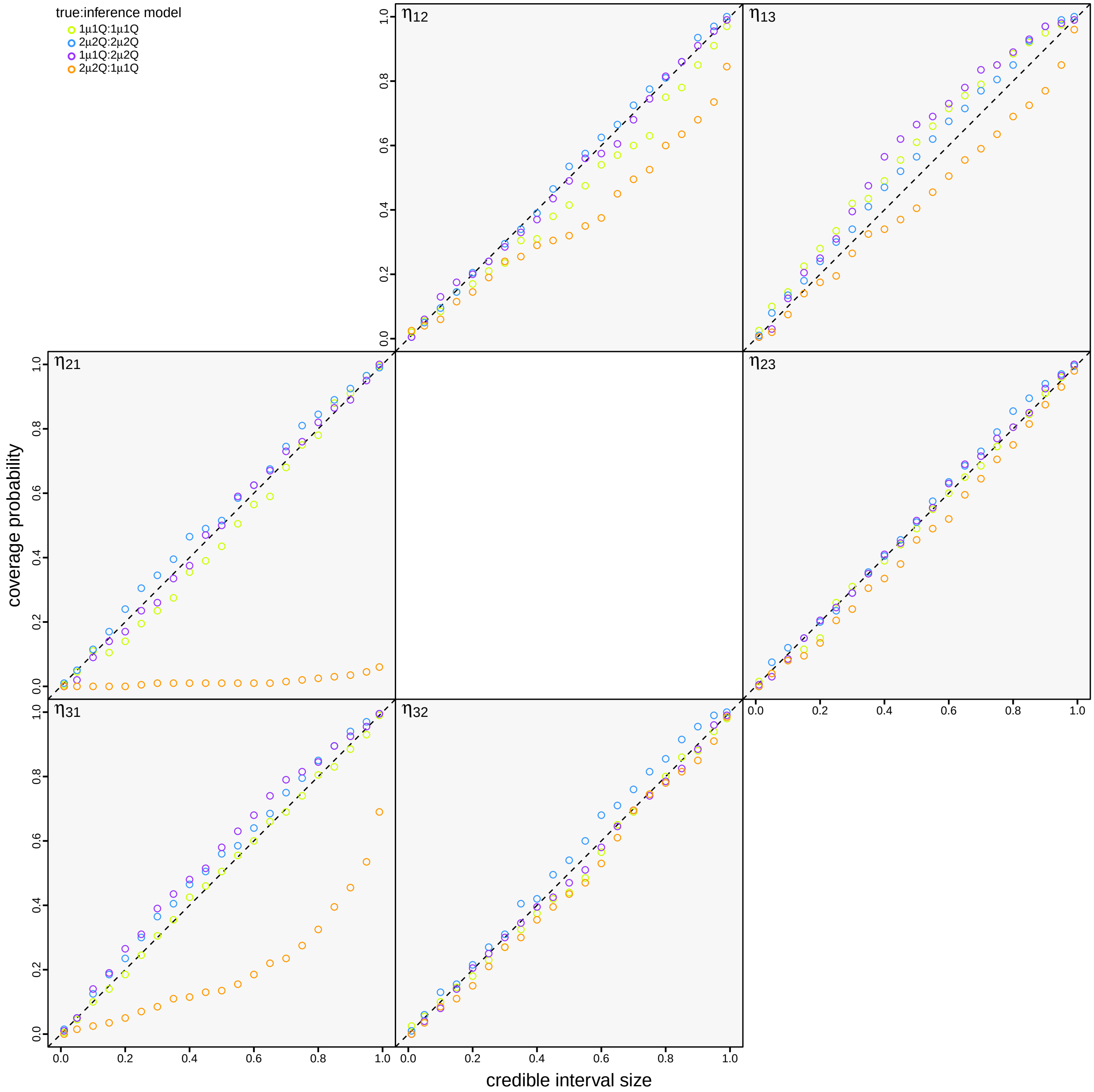


Figure S3: **Coverage probabilities for the number of dispersal events between each pair of areas.** For each true:inference model combination, we computed the coverage probability (the frequency with which the true number of dispersal events was contained in the $X$% credible interval; y-axis) as a function of the size ($X$) of the credible interval (x-axis). Each panel summarizes the estimated coverage probabilities for the number of dispersal events, $\eta$, between areas $i$ and $j$, $\eta_{ij}$. The panels are arranged to mirror the six pairwise, off-diagonal dispersal routes of the **Q** matrix; *e.g.*, the cell in the first row and second column depicts the estimated coverage probability for the number of dispersal events from area 1 to area 2, etc. When the model is true, we expect the coverage probability to be equal to the size of the credible interval (Cook et al. 2006). As expected, coverage probabilities fall along the one-to-one line when the model is correctly specified (green and blue). Moreover, coverage probabilities are also appropriate when the inference model is overspecified (*i.e.*, the inference model includes interval-specific parameters not included in the true model; purple). However, coverage probabilities are extremely unreliable when the inference model is underspecified (*i.e.*, the inference model excludes interval-specific parameters of the true model; orange).

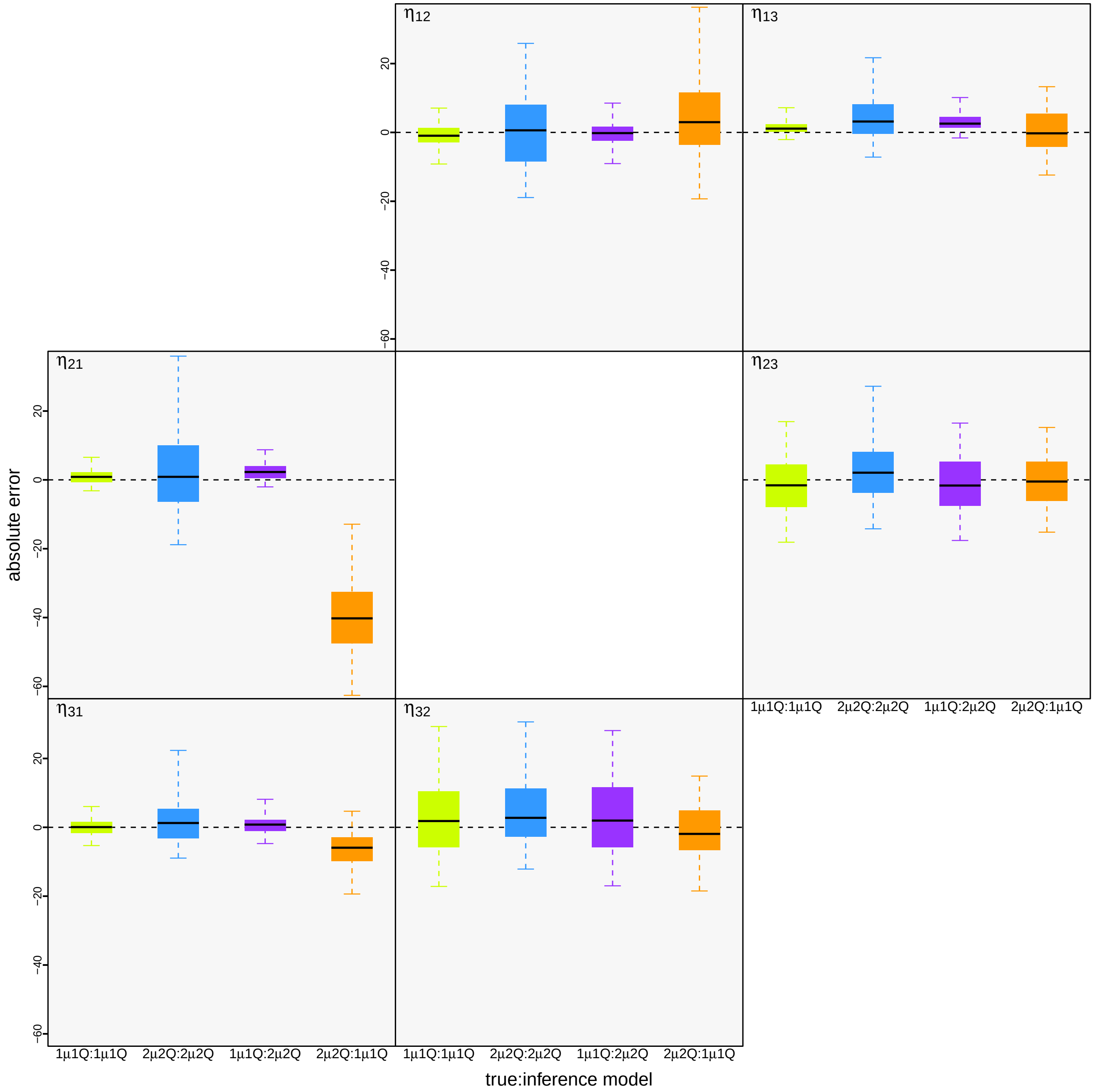


Figure S4: **Absolute error (estimated−true values) for the number of dispersal events between pairs of areas.** For each true:inference model combination, we summarized the absolute error for the number of dispersal events as boxplots (median [horizontal bar], 50% probability interval [boxes], and 95% probability interval [whiskers]). Each panel summarizes the absolute error for the number of dispersal events, $\eta$, between areas $i$ and $j$, $\eta_{ij}$. The six panels are arranged to mirror the corresponding six off-diagonal elements of the **Q** matrix (*c.f.*, Figure S3). Again, when the model is underspecified (orange) inferences are strongly biased compared to those under the correctly specified (green and blue) and overspecified (purple) models.

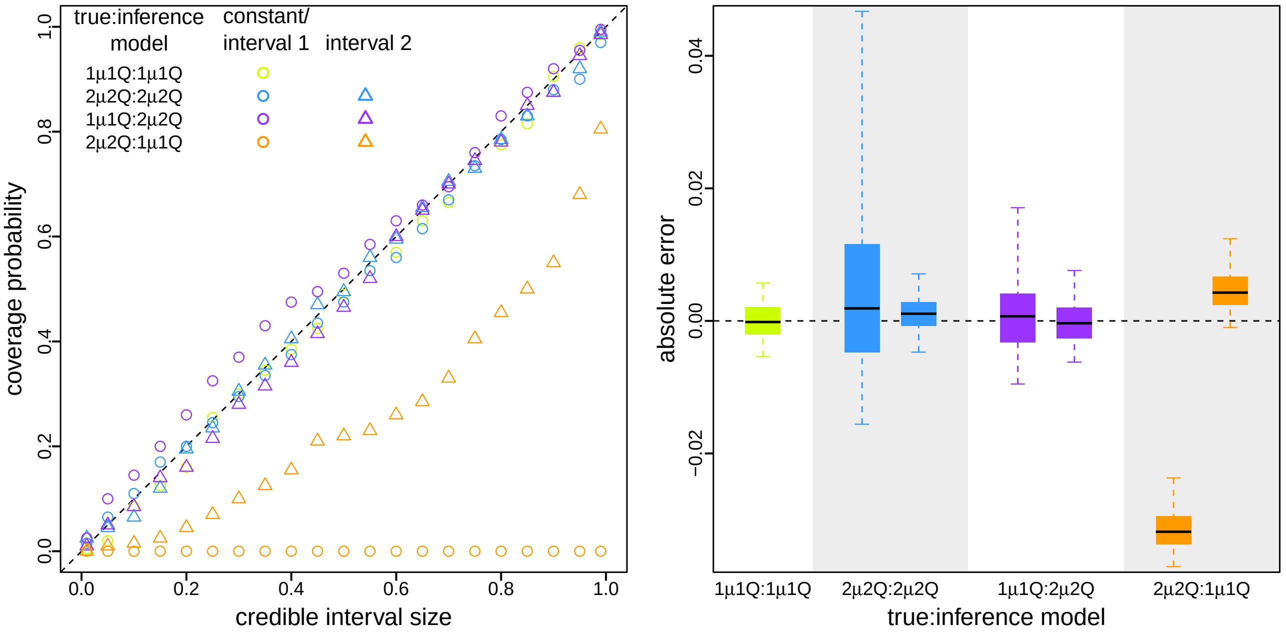


Figure S5: **Reliable inference of average dispersal rate requires a correctly specified discrete-geographic phylodynamic model.** We simulated 200 geographic datasets under each of two models, 1$\mu$1**Q**) 2$\mu$2**Q**. For each simulated dataset, we separately inferred the average dispersal rate under each model, resulting in four true:inference model combinations (1$\mu$1**Q**:1$\mu$1**Q**, 2$\mu$2**Q**:2$\mu$2**Q**, 1$\mu$1**Q**:2$\mu$2**Q**, and 2$\mu$2**Q**:1$\mu$1**Q**). For 1$\mu$1**Q**:2$\mu$2**Q**, we compared interval-specific parameter estimates to the true, time-constant parameter value (*i.e.*, we compared estimates of $\mu_1$ and $\mu_2$ to the true, time-constant value, $\mu$). Conversely, for 2$\mu$2**Q**:1$\mu$1**Q**, we compared the time-constant parameter estimates to each of the true, interval-specific values (*i.e.*, we compared estimates of $\mu$ to each of the true values, $\mu_1$ and $\mu_2$). Left) For each true:inference model combination, we plotted the coverage probability (y-axis) as a function of the size of the credible interval (x-axis). When the true or inference model is interval-specific, we plot separate true:inference comparisons for the first (circles) and second (triangles) time intervals. As expected (Cook et al. 2006), coverage probabilities fall along the one-to-one line when the model is correctly specified (green and blue); additionally, coverage probabilities are also appropriate when the inference model is overspecified (purple). However, coverage probabilities are extremely unreliable when the inference model is underspecified (orange). Right) For each true:inference model combination, we summarized the absolute error (estimated − true values) for the average dispersal rate as boxplots (median [horizontal bar], 50% probability interval [boxes], and 95% probability interval [whiskers]). When the true or inference model is interval-specific, we separately plot absolute error for the first (left) and second (right) intervals. Again, when the model is underspecified (orange) inferences are strongly biased compared to those under the correctly specified (green and blue) and overspecified (purple) models.

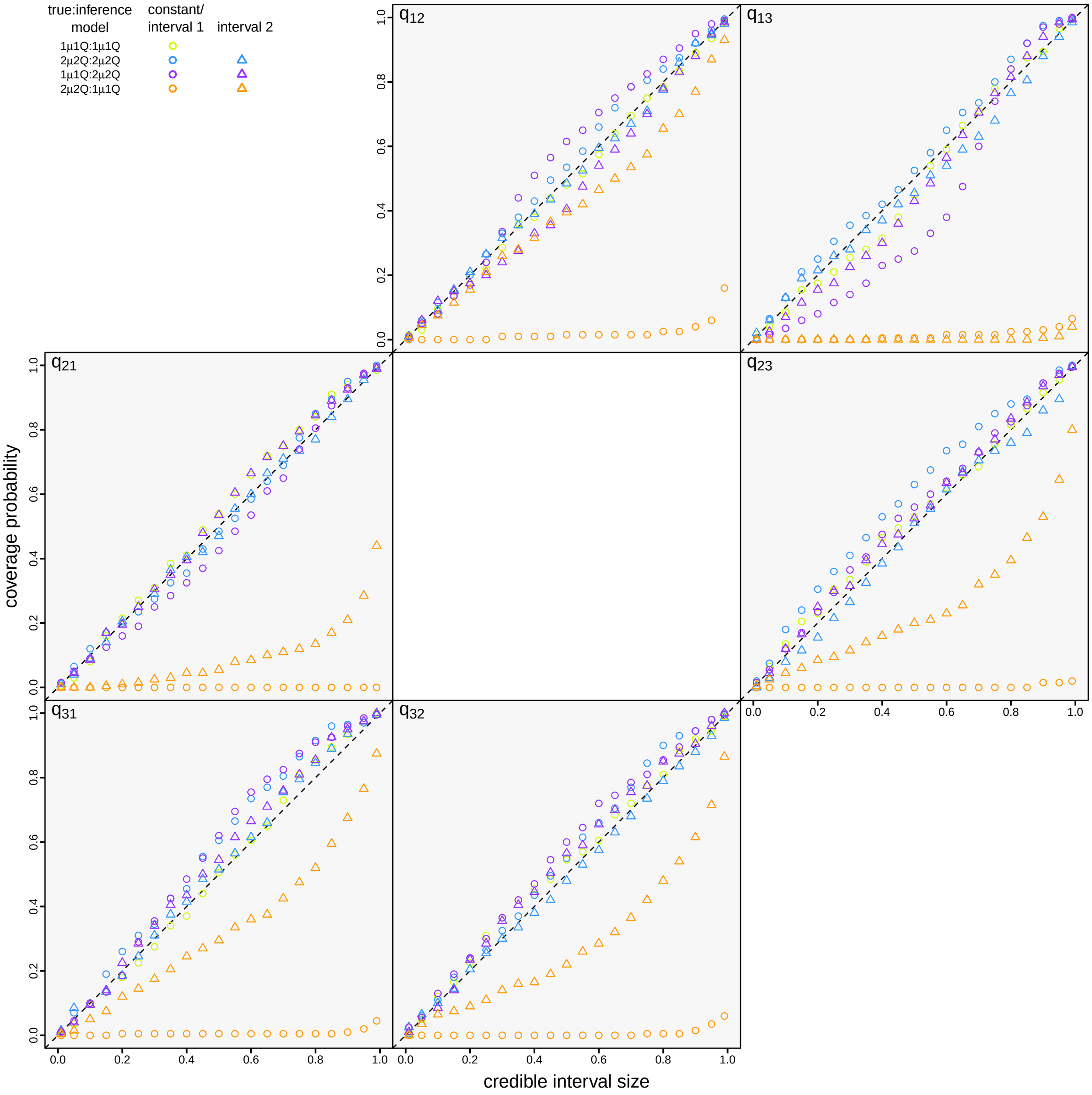


Figure S6: **Coverage probabilities for the relative dispersal rates between each pair of areas.** For each of the four true:inference model combinations, we plotted estimates of the coverage probability (y-axis) as a function of the size of the credible interval (x-axis). Each panel summarizes the estimated coverage probabilities for the relative dispersal rate between areas $i$ and $j$, $q_{ij}$. The panels are arranged to mirror the corresponding off-diagonal elements of the **Q** matrix (*c.f.*, Figure S3). For $1\mu1\mathbf{Q}$:$2\mu2\mathbf{Q}$, we compared interval-specific parameter estimates to the true, time-constant parameter value (*i.e.*, we compared estimates of $q_{ij,1}$ and $q_{ij,2}$ to the true, time-constant value, $q_{ij}$). Conversely, for $2\mu2\mathbf{Q}$:$1\mu1\mathbf{Q}$, we compared the time-constant parameter estimates to each of the true, interval-specific values (*i.e.*, we compared estimates of $q_{ij}$ to each of the true values, $q_{ij,1}$ and $q_{ij,2}$). When the true or inference model is interval-specific, we separately plot true:inference comparisons for the first (circles) and second (triangles) intervals. As expected (Cook et al. 2006), coverage probabilities fall along the one-to-one line when the model is correctly specified (green and blue); additionally, coverage probabilities are also appropriate when the inference model is overspecified (purple). However, coverage probabilities are extremely unreliable when the inference model is underspecified (orange).

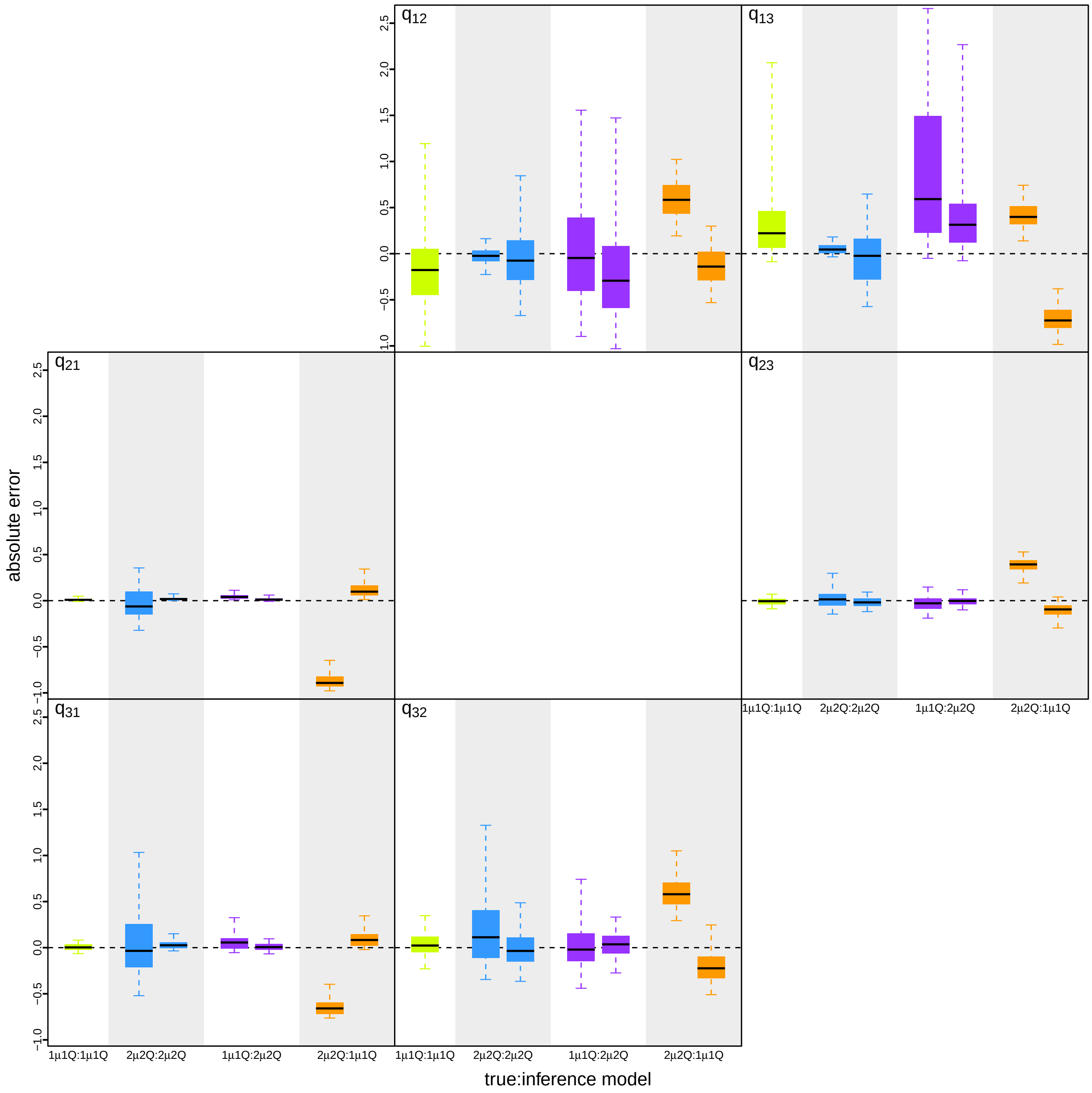


Figure S7: **Absolute error (estimated minus true values) for the relative dispersal rates between pairs of areas.** For each combination of true:inference models, we summarized the absolute error for the relative dispersal rates as boxplots (median [horizontal bar], 50% probability interval [boxes], and 95% probability interval [whiskers]). Each panel summarizes the absolute error for the relative dispersal rates between areas $i$ and $j$, $q_{ij}$. The panels are arranged to mirror the six off-diagonal relative dispersal rates of the **Q** matrix (*c.f.*, Figure S3). For 1$\mu$1**Q**:2$\mu$2**Q**, we compared interval-specific parameter estimates to the true, time-constant parameter value (*i.e.*, we compared estimates of $q_{ij,1}$ and $q_{ij,2}$ to the true, time-constant value, $q_{ij}$). Conversely, for 2$\mu$2**Q**:1$\mu$1**Q**, we compared the time-constant parameter estimates to each of the true, interval-specific values (*i.e.*, we compared estimates of $q_{ij}$ to each of the true, interval-specific values, $q_{ij,1}$ and $q_{ij,2}$). When the true or inference model is interval-specific, we separately plot absolute error for the first (left) and second (right) intervals. Again, when the model is underspecified (orange) inferences are strongly biased compared to those under the correctly specified (green and blue) and overspecified (purple) models.

#### S2.6 Assessing model fit to simulated datasets

##### Assessing relative fit of the true and alternative models using Bayes factors

We used Bayes factors to assess the relative fit of the true and alternative models to each simulated dataset. Specifically, for each dataset, we first estimated the marginal likelihood under each of two models ($1\mu 1\mathbf{Q}$ and $2\mu 2\mathbf{Q}$), and then computed the Bayes factor as twice the difference in the resulting log marginal likelihoods (Kass and Raftery 1995). We estimated marginal likelihoods for each inference model using both thermodynamic-integration (Lartillot and Philippe 2006) and stepping-stone (Xie et al. 2011; Baele et al. 2012) estimators. Our analyses to estimate marginal likelihoods conditioned on the MCC summary phylogeny (see Section S3.3).

For each simulated dataset, we ran two replicate power-posterior MCMC simulations for both models ($1\mu 1\mathbf{Q}$ and $2\mu 2\mathbf{Q}$) using our modified version of `BEAST` (see Section S1) with the `BEAGLE` library (compiled from the 'hmc-clock' branch, commit 'dd36bf5'; Ayres et al. 2019). For each replicate power-posterior MCMC simulation, we used 24 powers placed at evenly-spaced quantiles of a $\text{Beta}(0.3, 1.0)$ distribution. For each power, we discarded the initial 70000–80000 generations as burn-in and then sampled every 100 generations during the remaining 160000–180000 generations. (The number of generations and the length of the burn-in of each power are presented as ranges here and below because we deliberately ran the MCMC longer at each power for the power-posterior analyses under more complex models, and also set up replicate MCMCs with increasing length as one way of assessing the reliability of our marginal-likelihood estimates.) We assessed the reliability of our marginal-likelihood estimates by comparing values from all the replicate power-posterior MCMCs. Details of these analyses (*e.g.*, proposal weights) are available in the XML scripts included in our GitHub and Dryad repositories.

##### Assessing absolute fit of each model using posterior-predictive simulation

For each true:inference model combination, we assessed absolute model fit to each simulated dataset using posterior-predictive simulation (Gelman et al. 1996) with a set of 20 time-slice summary statistics. For each *simulated* dataset, we simulated $m = 800-1000$ *predictive* datasets using the parameter values that were randomly sampled from the inferred joint posterior distribution of the corresponding *simulated* dataset under each of the two models. We then generated posterior-predictive distributions from each set of $m$ predictive datasets under 20 separate time-slice summary statistics, *i.e.*, for all combinations of the two types of summary statistics (parsimony and tipwise-multinomial statistics) and 10 time slices spanning the entire dispersal history of the early phase of COVID-19 (including week 0 that covers the duration from the origin of SARS-CoV-2 to January 5, 2020, and weeks 1–9 corresponding to the nine weeks between January 6 and March 8, 2020). For each posterior-predictive distribution, we computed the posterior-predictive $p$ value (see Section S1.3) to assess the adequacy of (*i.e.*, absolute fit) the corresponding inference model.

##### Results

Our simulation study demonstrates the importance of identifying scenarios where an inference model is underspecified; failure to accommodate interval-specific variation in the study data will severely bias parameter estimates. Fortunately, our simulation study demonstrates that we can reliably identify when a given model is correctly specified, overspecified, or underspecified using a combination of Bayes factors (to assess the relative fit of competing models to the data; Fig. 4, left) and posterior-predictive simulation (to assess the absolute fit of each candidate model to the data; Fig. 4, right, Fig. S8 and table S2). Using a combination of Bayes factors and posterior-predictive simulation allows us to not only identify the best of the candidate models, but also to ensure that the best model provides an adequate description of the true process that gave rise to our study data.

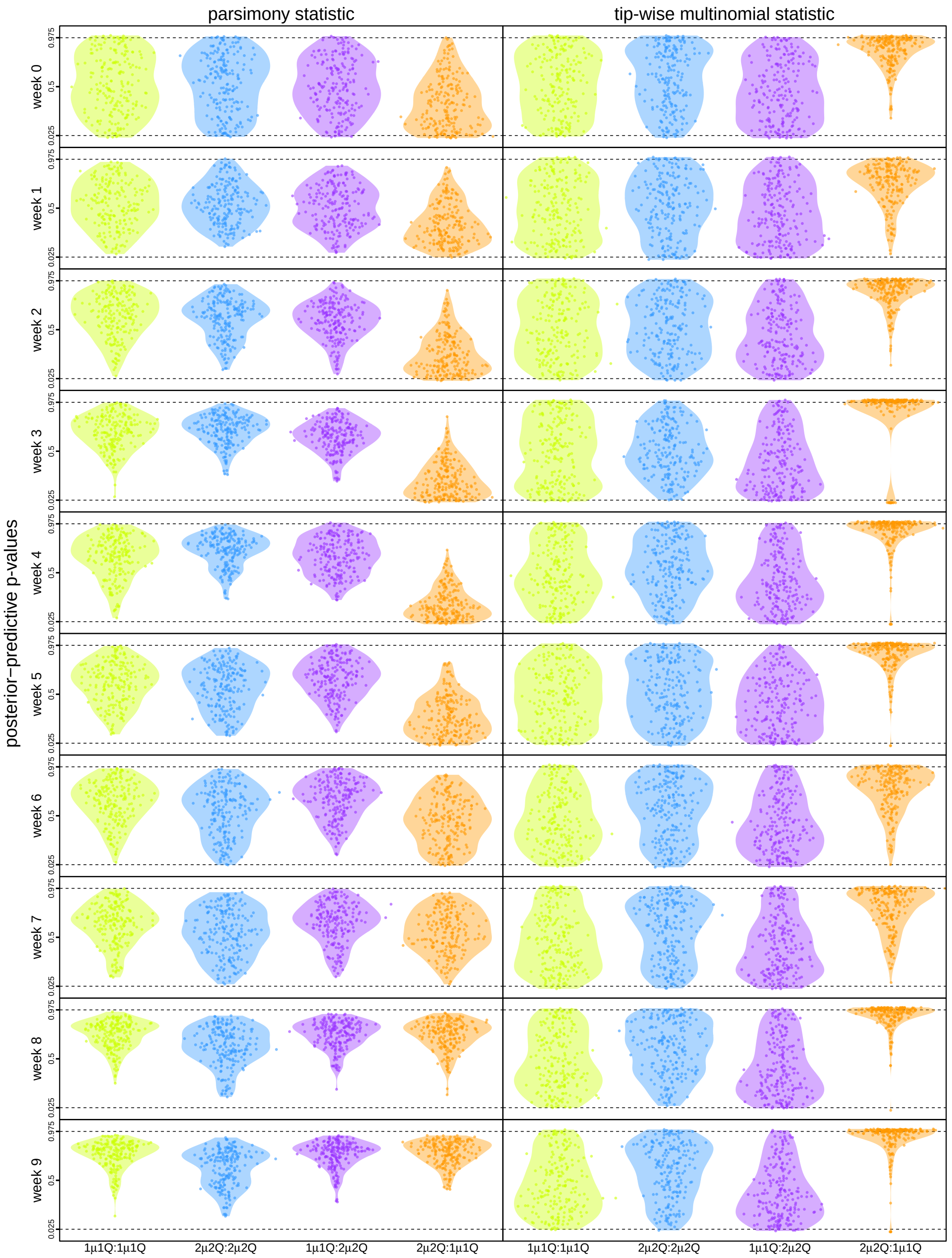


Figure S8: **Assessing the absolute fit of true and alternative discrete-geographic phylodynamic models to simulated datasets.** We assessed the absolute fit of alternative models to the simulated datasets. For each combination of true:inference model, we assessed absolute model fit (*i.e.*, model adequacy) using posterior-predictive simulation with a set of 20 time-slice summary statistics. We group results based on the parsimony (left column) and tipwise-multinomial (right column) summary statistics; the rows in each column corresponds to one of the 10 (weekly) time slices, and each cell plots the posterior-predictive distributions of the corresponding statistic for each of the four true:inference model combinations. Each dot represents the posterior-predictive $p$ value for a single dataset, and the violin plots summarize the distribution of these $p$ values for all datasets under the corresponding true:inference model combination. Dashed lines indicate critical posterior-predictive $p$ values (of 0.025 and 0.975); a dot above the top dashed line or below the bottom dashed line indicates that the corresponding inference model provides an inadequate description of the true process that gave rise to that dataset. As expected, the true model is overwhelmingly inferred to be adequate (green and blue). Encouragingly, model overspecification appears to have a negligible impact on model adequacy (purple). By contrast, an underspecified model severely impacts model adequacy (orange).

Table S2: **Percent of simulated datasets that were inadequately modeled.** The organization of the table mirrors that of Fig. S8. Each cell of the table indicates the percent of simulated datasets for which the inferene model was inferred to provide an inadequate description of the true process that generated the simulated datasets. That is, each cell indicates the percent of the posterior-predictive $p$ values that fall outside the critical (0.025 and 0.975) thresholds (*i.e.*, the corresponding percent of dots above the top dashed line or below the bottom dashed line in Fig. S8). Values indicating significant model inadequacy (*i.e.*, $\geq$5%) are indicated in red text.

| | parsimony statistic | | | | tipwise-multinomial statistic | | | |
|---|---|---|---|---|---|---|---|---|
| | $1\mu1\mathbf{Q}$:$1\mu1\mathbf{Q}$ | $2\mu2\mathbf{Q}$:$2\mu2\mathbf{Q}$ | $1\mu1\mathbf{Q}$:$2\mu2\mathbf{Q}$ | $2\mu2\mathbf{Q}$:$1\mu1\mathbf{Q}$ | $1\mu1\mathbf{Q}$:$1\mu1\mathbf{Q}$ | $2\mu2\mathbf{Q}$:$2\mu2\mathbf{Q}$ | $1\mu1\mathbf{Q}$:$2\mu2\mathbf{Q}$ | $2\mu2\mathbf{Q}$:$1\mu1\mathbf{Q}$ |
| week 0 | 4.5 | 3.5 | 3.0 | **6.0** | 4.5 | 4.0 | 5.0 | **11.5** |
| week 1 | 0.0 | 0.5 | 0.0 | 0.5 | 3.0 | **6.5** | 4.0 | 3.0 |
| week 2 | 0.5 | 0.0 | 0.0 | 3.5 | 4.0 | 3.5 | 2.5 | **16.0** |
| week 3 | 0.0 | 0.0 | 0.0 | **6.0** | 4.0 | 1.0 | 4.0 | **74.5** |
| week 4 | 0.0 | 0.0 | 0.5 | **7.5** | 2.5 | 4.0 | 3.0 | **37.5** |
| week 5 | 0.0 | 0.0 | 0.5 | 4.0 | 3.0 | 3.5 | 3.0 | **31.5** |
| week 6 | 0.0 | 0.5 | 0.0 | 1.0 | 3.5 | 5.0 | 2.5 | **7.5** |
| week 7 | 0.0 | 0.0 | 0.0 | 0.0 | 5.0 | 2.5 | 5.0 | **6.5** |
| week 8 | 0.0 | 0.0 | 0.0 | 0.0 | 1.0 | 1.0 | 1.5 | **36.0** |
| week 9 | 0.0 | 0.0 | 0.0 | 0.0 | 2.0 | 2.5 | 2.0 | **45.5** |
| average | 0.5 | 0.45 | 0.4 | 2.85 | 3.25 | 3.35 | 3.25 | **26.95** |

### S3 Empirical Application

#### S3.1 Overview

The results of our empirical study are based on a complex and comprehensive series of computationally intensive analyses. In this section, we provide a high-level overview of our data collection and data analyses to clarify the rationale of our empirical study, while directing readers to the corresponding subsections below that provide additional details on the various analyses that we performed.

##### Data Acquisition and Curation

###### Epidemiological data

We used two types of epidemiological information in this study: (1) the number of confirmed COVID-19 cases, and; (2) the intervention measures involving China that were enacted during the early phase of the pandemic. We compiled a dataset of the number of confirmed COVID-19 cases recorded on each day for each country/province/state based on various sources (WHO 2020; DXY 2020; NHCPRC 2020; ECDC 2020; USCDC 2020) via two intermediate portals (Wu et al. 2020b; Dong et al. 2020; see Section S3.2). We used these case-number data to assess the fraction of total cases represented by our genomic sequences, and to estimate the approximate date by which SARS-CoV-2 had spread to most geographic areas. We collected information on international travel bans with China and domestic mitigation measures within China from multiple sources (Wikipedia 2020a,b; Kraemer et al. 2020; Tian et al. 2020; Hsiang et al. 2020; Lai et al. 2020; see Section S3.2).

###### Travel data

We used the daily number of commercial passenger flights obtained from FlightAware as a proxy for the global air-travel volume (see Section S3.2).

###### Delineation of time intervals and geographic areas

To explore the dynamics of viral geographic dispersal in the early phase of the COVID-19 pandemic, we partitioned the study period into five time intervals: (1) interval 1 from late 2019 (origin time) to Jan. 12, 2020; (2) interval 2 from Jan. 1 3, 2020 to Jan. 25, 2020; (3) interval 3 from Jan. 26, 2020 to Feb. 2, 2020; (4) interval 4 from Feb. 3, 2020 to Feb. 16, 2020, and; (5) interval 5 from Feb. 16, 2020 to Mar. 8,

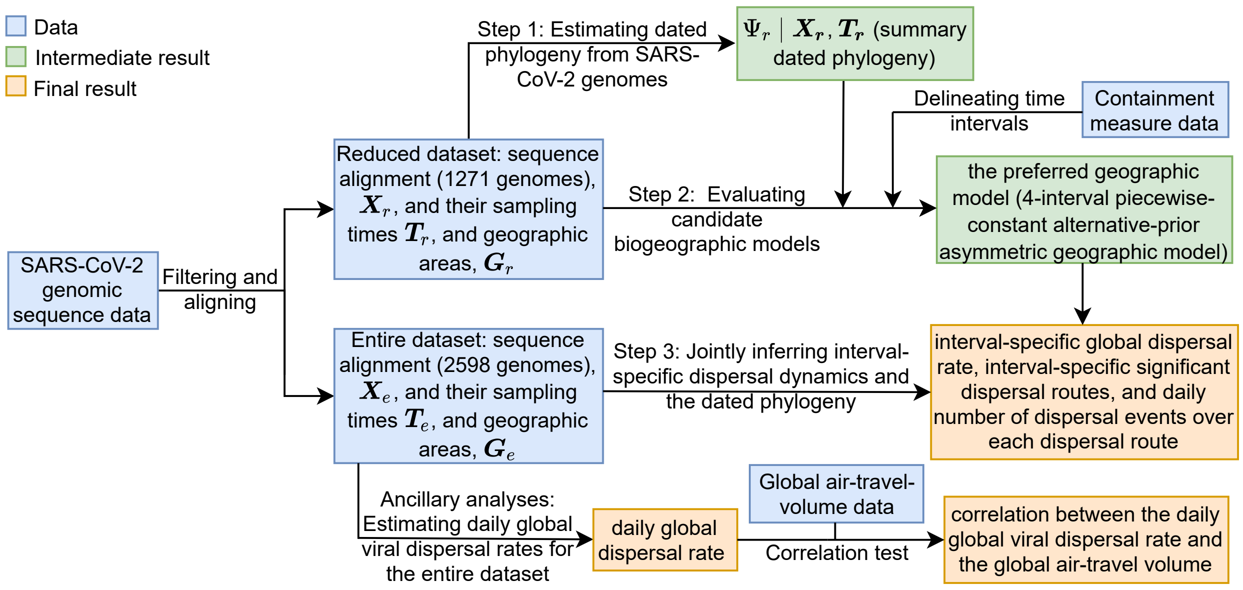


Figure S9: **Workflow of the empirical analyses in our study.**

2020. Boundaries between these intervals coincide with the initiation of containment measures (*e.g.*, international travel bans with China) or other events associated with changes in the level of population movement (*e.g.*, start of the Spring Festival travel season); see Section S3.2. For our phylodynamic analyses, we discretized the globe into geographic areas to study the early spread of SARS-CoV-2. We grouped geographically adjacent countries/territories (for non-focal regions) or states/provinces (for focal regions) to specify a total of 23 geographic areas (Fig. S10).

##### SARS-CoV-2 genomic sequence data

We curated two genomic sequence datasets for this study, one with 1271 sequences (the "reduced dataset") and the other with 2598 sequences (the "entire dataset"). The reduced dataset was produced on Apr. 19, 2020, based on all available SARS-CoV-2 genomic sequences from the Global Initiative on Sharing All Influenza Data (GISAID, Shu and McCauley 2017) as of that date. The entire dataset was produced by adding sequences that were available on GISAID as of Sept. 22, 2020. See Section S3.2 for details on the sequence curation and alignment, and differences between the reduced and entire datasets.

#### Phylodynamic Analyses

Our objective is to infer the joint posterior probability distribution of the viral phylogeny, divergence times, and biogeographic history under a composite phylodynamic model that is appropriate for the entire SARS-CoV-2 dataset. The composite phylodynamic model is comprised of four main components: (1) a substitution model that describes the evolution of nucleotide sequences over the tree; (2) a branch-rate prior model that characterizes how rates of substitution vary across branches of the tree; (3) a branching-process model that specifies the prior distribution of the tree topologies and divergence times, and; (4) a biogeographic model that describes how viruses disperse between geographic areas.

For each of these components, there are numerous candidate models; the vast space of composite phylodynamic models makes it computationally prohibitive to evaluate the fit of each candidate model to the entire dataset. Accordingly, we adopt a three-step model-selection procedure: (1) we first estimate the dated phylogeny for the reduced dataset under a relaxed-clock model with biologically motivated specification of the substitution model, branch-rate prior model, and branching-process prior model; (2) we then condition on the resulting dated phylogeny to select among candidate biogeographic models using the reduced dataset, and; (3) finally, we perform joint inference of the dated phylogeny and biogeographic history for the entire dataset using the preferred composite phylodynamic model.

##### Step 1: Estimating the dated phylogeny of the reduced dataset

We inferred a dated phylogeny by performing Bayesian analyses of the reduced SARS-CoV-2 sequence dataset under a relaxed-clock model, which includes the first three of the four model components of the composite phylodynamic model: (1) a substitution model; (2) a branch-rate prior model, and; (3) a branching-process prior model.

Specifically, we specified a partitioned substitution model to accommodate possible variation in the evolutionary process across genomic regions. The SARS-CoV-2 genome is comprised of 11 gene regions (we list these gene regions and their corresponding coordinates in the reference genome in Table S3). We partitioned the SARS-CoV-2 genomes into six data subsets, with three subsets for the ORF1ab gene region (one for each codon position), and three subsets for the remaining ten combined gene regions (one for each codon position). For each of these data subsets, we specified an independent TN93 substitution model (Tamura and Nei 1993). We used partition-specific rate multipliers to capture differences in the substitution rate across the six data subsets. We specified a discrete-gamma model to accommodate substitution-rate variation across sites within each data subset. Our preliminary analyses specified an independent discrete-gamma model for each of the six data subsets, which revealed a similar degree of among-site rate variation within each data subset (*i.e.*, with similar posterior estimates of the six $\alpha$-shape parameters). Accordingly, to decrease model complexity, we specified a shared, discrete-gamma model (Yang 1994) to accommodate substitution-rate variation across sites of the entire alignment. We specified an uncorrelated lognormal (UCLN) branch-rate prior model (Drummond et al. 2006; Li and Drummond 2012; Rannala and Yang 2007) by drawing i.i.d. rate multipliers for each branch from a shared underlying lognormal distribution, where the parameters of this distribution (mean and standard deviation) are estimated from the data. For the branching-process prior model, we used a coalescent model with exponential population growth.

We performed MCMC simulations to approximate the joint posterior distribution of the relaxed-clock model parameters and the dated phylogeny using `BEAST` (Suchard et al. 2018). We then used `TreeAnnotator` to generate a summary phylogeny from the combined posterior sample of dated phylogenies as a maximum clade credibility (MCC) tree. We provide a more detailed description of these analyses in Section S3.3. The phylogeny inferred from these analyses was used both for our simulation study and also in the next step to evaluate candidate biogeographic models.

**Step 2: Evaluating candidate biogeographic models using the reduced dataset**

We explored a pool of nine candidate biogeographic models. These models assign interval-specific parameters—for the average rate of viral dispersal, $\mu$, and/or relative rates of viral dispersal, $\mathbf{Q}$—to one, two, four, or five pre-specified time intervals; *i.e.*, $1\mu1\mathbf{Q}$, $1\mu2\mathbf{Q}$, $2\mu1\mathbf{Q}$, $2\mu2\mathbf{Q}$, $1\mu4\mathbf{Q}$, $4\mu1\mathbf{Q}$, $4\mu4\mathbf{Q}$, $5\mu5\mathbf{Q}$, and $5\mu5\mathbf{Q}^*$. We specified interval boundaries based on external information regarding events within the study period that might plausibly impact viral dispersal dynamics, including: (A) start of the Spring Festival travel season in China (the highest annual period of domestic travel, January 12); (B) onset of mitigation measures in Hubei province, China (January 26); (C) onset of international air-travel restrictions against China (February 2), and; (D) relaxation of domestic travel restrictions in China (February 16). Phylodynamic models with two intervals include event C, models with four intervals include events A, C, and D, and the $5\mu5\mathbf{Q}$ model includes all four events. The final candidate model, $5\mu5\mathbf{Q}^*$, includes five arbitrary and uniform (bi-weekly) intervals.

We assessed both the *relative* and *absolute* fit of these candidate biogeographic models to our reduced SARS-CoV-2 dataset. We assessed the *relative fit* of competing biogeographic models by computing Bayes factors based on their marginal-likelihood estimates. We performed power-posterior MCMC simulations using `BEAST` (Suchard et al. 2018) to estimate marginal likelihoods using both thermodynamic-integration (Lartillot and Philippe 2006) and stepping-stone (Xie et al. 2011; Baele et al. 2012) estimators. We also assessed the *absolute fit* of each model using posterior-predictive simulation (Gelman et al. 1996). For each model, we first inferred the joint posterior distribution from the observed biogeographic data (*i.e.*, the geographic location of each sampled sequence) by performing MCMC simulations using `BEAST` (Suchard et al. 2018). We then simulated predictive datasets by repeatedly sampling at random from the corresponding joint posterior probability distribution for a given the model. Finally we generated posterior-predictive distributions from each predictive dataset under various summary statistics (as described in Section S1.3), which measure the discrepancy between the observed dataset and the simulated dataset. We provide a more detailed description of these analyses in Section S3.3. The preferred biogeographic model identified by these analyses was then used in our subsequent joint phylodynamic analyses, described below.

**Step 3: Joint phylodynamic inference of the entire dataset**

We performed joint inference of the phylogeny, divergence times, and biogeographic history using the entire SARS-CoV-2 dataset based on a phylodynamic model that includes (1) a relaxed-clock model, and (2) a biogeographic model. The relaxed-clock model specified in these joint analyses is identical to that specified in Step 1 (with minor changes in the prior specification to reflect differences in viral sampling). The biogeographic model specified in these joint analyses is identical to the biogeographic model selected in Step 2: the 4-interval ($4\mu4\mathbf{Q}$) model.

We performed MCMC simulations to approximate the joint posterior distribution of the phylodynamic-model parameters using BEAST (Suchard et al. 2018). We also performed posterior-predictive simulation to confirm that the preferred biogeographic model provides an adequate fit to the entire SARS-CoV-2 dataset under the joint inference. We provide a more detailed description of these analyses in Section S3.3.

**Ancillary analyses: Estimating daily global viral dispersal rates for the entire dataset**

We performed additional analyses to explore the correlation between daily global air-travel volume and daily average global SARS-CoV-2 dispersal rate during the early phase of the COVID-19 pandemic. We first estimated the daily global dispersal rate using the entire SARS-CoV-2 dataset under a more granular interval-specific phylodynamic model that allows daily variation in the average viral dispersal rate. We then computed the correlation between these estimates and independent information on the daily volume of global air travel during this period. The phylodynamic model we specified for these analyses was identical to that used in Step 3, except that we further discretized the time intervals for the average global dispersal rate to vary daily. In these analyses, we accommodated phylogenetic uncertainty by averaging over the marginal posterior probability distribution of dated phylogenies inferred in Step 3. We performed MCMC simulations to approximate the joint posterior distribution of the daily-rate model parameters using BEAST (Suchard et al. 2018). We then performed a standard correlation test between the daily global air-travel volume and the estimated mean daily global SARS-CoV-2 dispersal rates by computing Pearson's r and the corresponding $p$ value, focussing on the period spanning from Jan. 31 (by which date the virus achieved a global distribution) to Mar. 8, 2020. We provide a more detailed description of these analyses in Section S3.3.

#### Computational Burden of Interval-Specific Discrete-Geographic Phylodynamic Models

Our interval-specific models extend the model developed by Bielejec et al. (2014); the computational complexity of our model is effectively identical to their model (see Bielejec et al. 2014 for details). Here we provide additional informal observations on the computational burden associated with our model.

Generally speaking, compared to the constant-rate model, interval-specific models require more computational effort both because of the additional computations at each iteration of the MCMC and because more MCMC cycles are required to collect sufficient samples to ensure acceptable ESS values for all parameters. Regarding the computation per MCMC cycle, interval-specific models usually require additional round(s) of matrix multiplication in computing the transition-probability matrix of each branch for both likelihood calculation (when the relative rates of dispersal vary across intervals) and stochastic mapping (when either the average or the relative rates of dispersal vary across intervals). The increase in computational burden scales with the number of intervals (or more precisely, it scales with the number of interval boundaries that each branch overlaps, summed over all branches; *i.e.*, the additional computation is minimized in the extreme case where interval boundaries only overlap with nodes but not any branches). For discrete-geographic inferences, the additional computation per MCMC iteration seems to be on the same scale as the likelihood computation under the constant-rate model (*e.g.*, in our simulation study, the $2\mu 2\mathbf{Q}$ model appeared to be about twice as slow per iteration compared to the $1\mu 1\mathbf{Q}$ model).

Regarding the computation per MCMC simulation, given that the number of free parameters scales linearly with the number of intervals, we expect to run the BEAST MCMC simulations for $m$ times more iterations for an $m+1$-interval model compared to the constant-rate model. Similar to the statistical performance issue discussed above, we think that whether a given parameter is harder or easier to sample (*i.e.*, fewer or more effective samples per iteration) under an interval-specific model compared to the constant-rate model is an empirical question. In summary, analyses under an interval-specific model may be 2 to 10 times slower compared to those under the constant-rate model. In our experiments, analyses under the interval-specific models took about a few days to a week to complete.

#### Data and Code Availability

GISAID accession IDs of the SARS-CoV-2 sequences used in this study, as well as the flight-volume data (obtained from FlightAware, LLC) and intervention-measure data, are maintained in the GitHub repository (https://github.com/jsigao/interval_specific_phylodynamic_models_supparchive) and archived in the Dryad repository (https://datadryad.org/stash/share/vTbeDwLq2uSL9rL4NCe_Cocp2bY7BgWTI2tUgoNrLDA). Our repositories also contain BEAST XML scripts used to perform the phylodynamic analyses, R scripts used to perform simulations and post processing, and a modified version of the BEAST program used for some of the analyses in this study.

#### S3.2 Detailed Description of Data Acquisition and Curation

##### Epidemiological Data

###### COVID-19 case numbers

We obtained the number of confirmed COVID-19 cases from five major sources: (1) the WHO COVID-19 situation reports (WHO 2020), (2) the COVID-19 dashboard published on a Chinese medical website, Ding Xiang Yuan (DXY), that integrates data from local governmental reports (DXY 2020), (3) the National Health Commission of the People's Republic of China (NHCPRC) COVID-19 situation reports (NHCPRC 2020), (4) the European Centre for Disease Prevention and Control (ECDC) COVID-19 situation update (ECDC 2020), and (5) the US Centers for Disease Control and Prevention (USCDC) COVID-19 data tracker (USCDC 2020). Rather than directly collecting data from these sources, we accessed them via two intermediate portals: the `R` (R Core Team 2020) package `nCov2019` (Wu et al. 2020b), and the COVID-19 Data Repository by the Center for Systems Science and Engineering at Johns Hopkins University (Dong et al. 2020).

###### Intervention measures

We focused on two types of intervention measures enacted during the early phase of the COVID-19 pandemic that involved China: targeted-containment measures involving China (*i.e.*, international air-travel bans), and domestic-mitigation measures within China. We compiled information on these containment measures from various news reports, Wikipedia pages (Wikipedia 2020a,b), and peer-reviewed publications (Kraemer et al. 2020; Tian et al. 2020; Hsiang et al. 2020; Lai et al. 2020). For domestic measures within China, we focused on measures that were likely to interrupt travel among regions, including lockdowns at city or province levels, inter-city travel restrictions, and home or neighborhood isolation. See `international_airtravelban_withchina.csv` for a collection of countries or territories that enacted international travel bans with China (and the associated initiation date), and `china_domestic.csv` for a collection of provinces or cities that enacted mitigation measures in China (including the associated implementation period and type of measure); these spreadsheets are included in our GitHub and Dryad repositories.

##### Travel Data

We acquired global air-travel-volume data from FlightAware, detailing the number of all commercial passenger flights (subdivided by each aircraft type) per day between Dec. 30, 2019 and Mar. 8, 2020. We transformed these daily aircraft-volume data to provide an estimate of the daily air-travel passenger volume (Fig. 5, dashed line) by multiplying the number of flights for each type of aircraft by the capacity (seat number) for the corresponding type of aircraft. The original air-travel-volume data are contained in `nflights_daily_byaircraft.csv`, and the number of seats for each aircraft type is provided in `aircraft_nseats.csv` (included in our GitHub and Dryad repositories).

##### Definition of Time Intervals

We partitioned the early phase of the COVID-19 pandemic into five time intervals: (1) interval 1 from late 2019 (origin time) to Jan. 12, 2020; (2) interval 2 from Jan. 13 to Jan. 25; (3) interval 3 from Jan. 26 to Feb. 2; (4) interval 4 from Feb. 3 to Feb. 16, and; (5) interval 5 from Feb. 17 to Mar. 8, 2020.

The Jan. 12 boundary coincides with the start of the Spring Festival travel season in China (the highest annual period of domestic travel). The Jan. 26 boundary coincides with onset of widespread mitigation measures in China to restrict domestic travel: these measures began with the city-wide lockdown of Wuhan on Jan. 23 (that were extended to the entire Hubei province in the following days), followed by the declaration of level-1 emergency in all mainland provinces between Jan. 24–29, the extension of the Spring Festival national holiday (effectively school and workplace closure) announced on Jan. 27, and the enactment of stringent home- or neighborhood-isolation orders in various cities outside Hubei beginning Feb. 2. The Feb. 2 boundary coincides with the initiation of international air-travel bans with China (imposed by 34 countries by this date) and the cancellation (or significant reduction) of international air services involving China (by over 130 airlines; International Civil Aviation Organization 2020; Wikipedia 2020b), following the declaration of Public Health Emergency of International Concern (PHEIC) by the World Health Organization (WHO) on Jan. 31. The Feb. 17 boundary coincides with the lifting of travel restrictions in China (except in Hubei, where the travel restrictions were not lifted until late Mar.).

#### SARS-CoV-2 Genomic Sequence Data

We curated two SARS-CoV-2 genomic sequence datasets for our study, one with 1271 sequences (the "reduced dataset") and the other with 2598 sequences (the "entire dataset").

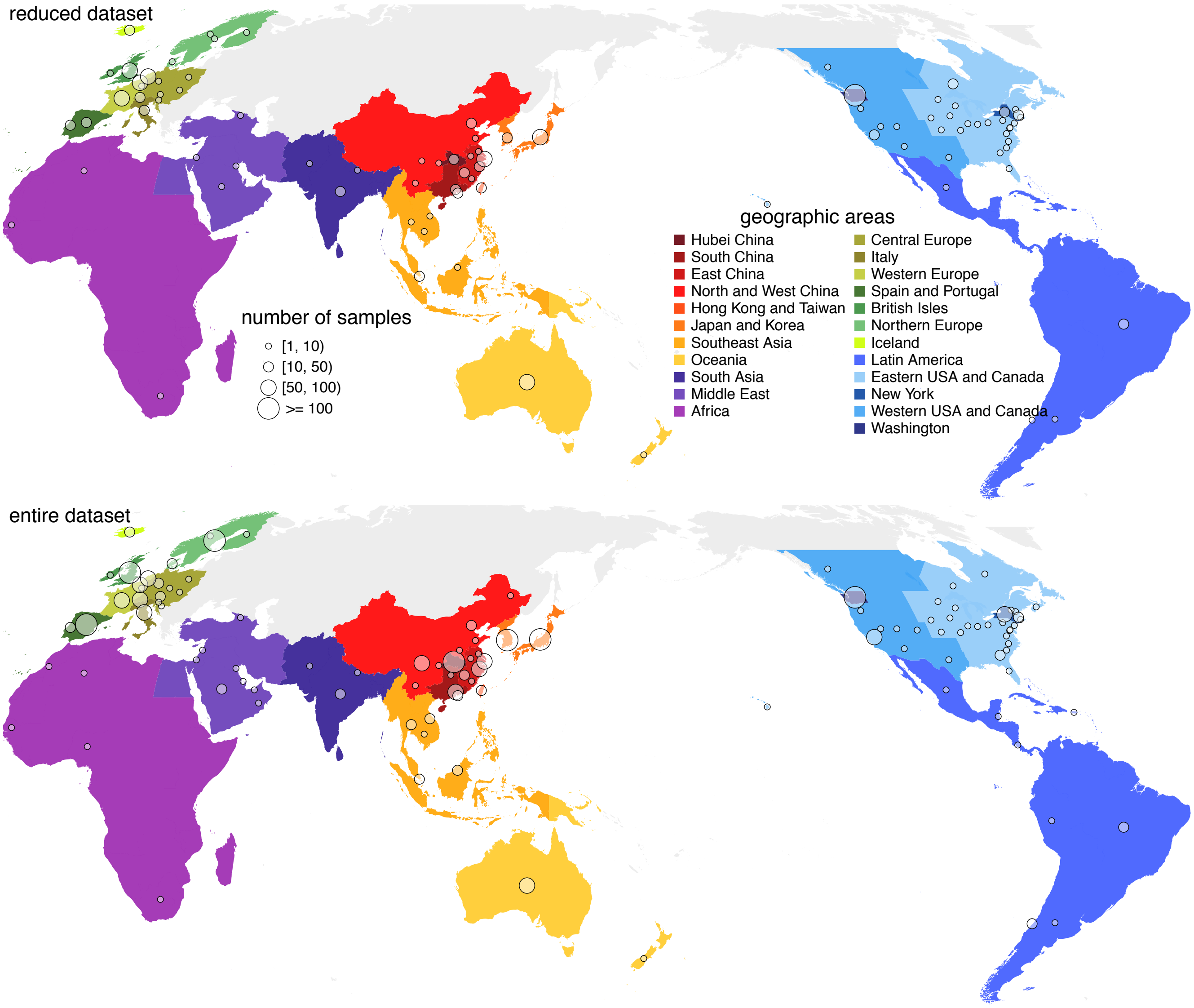


Figure S10: **SARS-CoV-2 genomes sampled from each discretized geographic area.** Our study includes two SARS-CoV-2 datasets—the reduced (1271 sequences, top) and entire (2598 sequences, bottom)—that comprise viral genomes collected between Dec. 24, 2019–Mar. 8, 2020 from 23 discrete geographic areas (colored regions); circles indicate the number and location of samples in our study.

##### Assembling the reduced dataset

The reduced dataset consists of all available SARS-CoV-2 genomic sequences available as of Apr. 19, 2020 from GISAID (https://www.gisaid.org/; Shu and McCauley 2017). As our focus is on the crucial early phase of the COVID-19 pandemic, we excluded sequences that were collected after Mar. 8, leaving 2003 sequences in the dataset. We first filtered the dataset by excluding sequences that fit any of the following conditions: (1) fewer than 29000 sites (not counting missing or gap sites); (2) lacking associated metadata (*e.g.*, sampling time or location); (3) lacking the precise sampling date or geographic location (state/province for sequences from China, Canada, or U.S.A. and country for the others); (4) sampled from a non-human host; (5) multiple sequences from the same individual (in which case we randomly selected one of sequence and discarded the others), or; (6) duplicates of other sequences in the dataset [for this purpose, we used the "exclude list" used by `Nextstrain` (https://github.com/nextstrain/ncov/blob/master/defaults/exclude.txt) as a reference]. Application of these filters resulted in a genomic dataset consisting of 1620 sequences.

We then inferred an alignment of these nucleotide sequences using `MUSCLE` version 3.8 (Edgar 2004). We performed a second round of filtration of the resulting alignment. First, we excluded sequences that appeared to be anomalously divergent; this was achieved by comparing each sequence to the reference genome while assuming that the rate of mutation accumulation should not exceed 10 mutations per genome per month. We also excluded sequences with many ambiguous sites (*i.e.*, sites for which the nucleotide could not be unambiguously identified); specifically, we discarded sequences with more than 15 ambiguous sites, and sequences with at least 10 ambiguous sites and fewer than 10 sites differing from

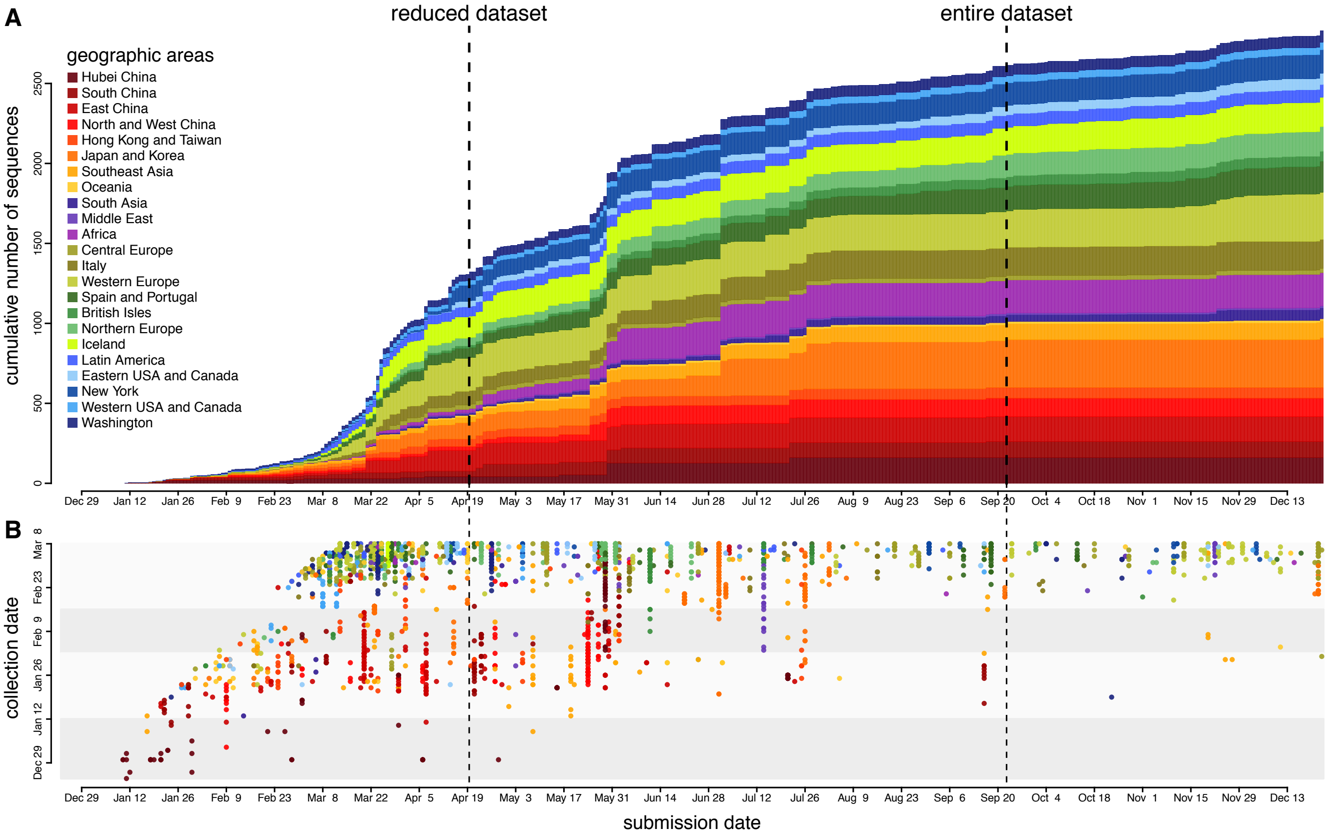


Figure S11: **Submission and collection dates for SARS-CoV-2 genomic sequences. (A)** Cumulative number of SARS-CoV-2 sequences submitted to GISAID that were collected during the early phase of the COVID-19 pandemic. The color of each segment in the stacked bar plot indicates the number of sequences submitted from the corresponding geographic area on that day. **(B)** Submission and collection dates for each SARS-CoV-2 sequence included in our study. The deposition rate of sequences collected prior to Mar. 8 drastically decreased in Sept. 2020.

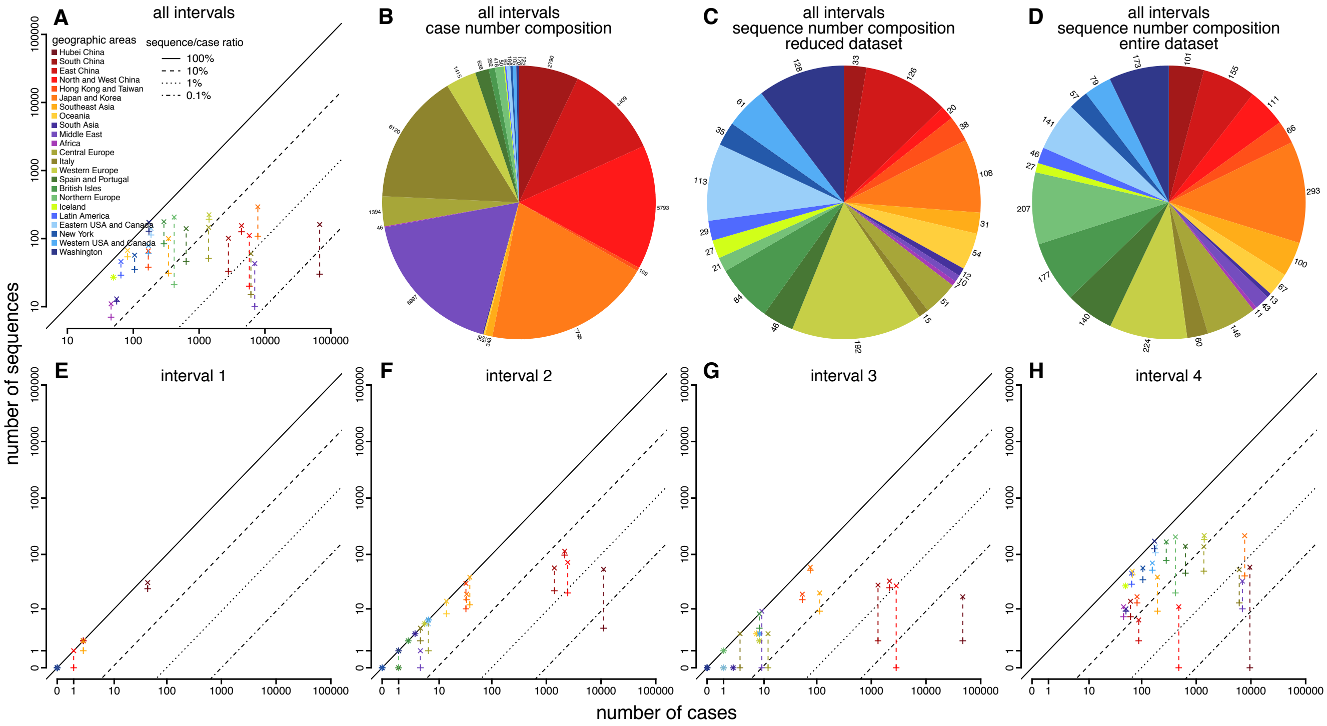


Figure S12: **Number of SARS-CoV-2 genome sequences versus number of confirmed COVID-19 cases. (A)** Number of sequences versus number of confirmed cases across the early phase of the pandemic. + and x indicate the reduced and entire datasets, respectively; they are connected by dashed line for each geographic area to show the increase of the number of sequences (and thus the sequence/case ratio) in the entire dataset. **(B)** Geographic distribution of confirmed case numbers (excluding Hubei). **(C)** Geographic distribution of sequence number (excluding Hubei; reduced dataset). **(D)** Geographic distribution of sequence number (excluding Hubei; entire dataset). **(E–H)** Number of sequences versus number of confirmed cases for each interval of the early phase, respectively.

the reference genome. Next, we excluded sequences with nonsense mutations; to this end, we translated the nucleotide alignment into an amino-acid alignment using the `seqinr` package (Charif and Lobry 2007) in `R` (R Core Team 2020) to identify sequences with premature stop codons. We assumed that the rate of amino-acid substitution accumulation should not exceed 4 substitutions per genome per month; we therefore discarded sequences with more than 6 ambiguous amino-acid sites, and sequences with at least 3 ambiguous amino-acid sites but fewer than 3 sites differing from the reference genome. After filtering, our reduced dataset included 1271 sequences.

#### Assembling the entire dataset

We also compiled a more comprehensive dataset by curating all sequences available from GISAID as of Sept. 22, 2020. Specifically, we downloaded an alignment from GISAID, which was inferred using `MAFFT` (Katoh and Standley 2013). After excluding sequences that were collected after Mar. 8, 2020, the alignment included 4012 sequences. We then performed the same two-step filtration procedure that we applied to the reduced dataset, culminating in an alignment (“entire dataset”) with 2598 sequences.

The entire dataset is more comprehensive than the reduced dataset: it contains more than twice the number of sequences (Fig. S11), and is also more evenly sampled, as the sequence-to-case ratios of many undersampled geographic areas are significantly higher, especially for the third and fourth intervals of our study (Fig. S12). Moreover, the entire dataset contains SARS-CoV-2 genomic sequences that are likely to represent the vast majority of such data that will ever be available; the deposition rate of sequences collected from the early phase of the pandemic drastically decreased in Sept., 2020 (Fig. S11).

Table S3: **Genomic coordinates of the SARS-CoV-2 coding regions.**

| Region | Starting coordinate | Ending coordinate |
|---|---|---|
| ORF1ab* | 266 | 21555 |
| S | 21563 | 25384 |
| ORF3a | 25393 | 26220 |
| E | 26245 | 26472 |
| M | 26523 | 27191 |
| ORF6 | 27202 | 27387 |
| ORF7a | 27394 | 27759 |
| ORF7b | 27756 | 27887 |
| ORF8 | 27894 | 28259 |
| N | 28274 | 29533 |
| ORF10 | 29558 | 29674 |

*During translation, ORF1ab experiences a $-1$ ribosomal frameshift at site 13468, so the range is (266–13468, 13468–21555).

##### Trimming and partitioning the curated alignment

For each curated dataset, we trimmed the 5′UTR and 3′UTR as well as the other non-coding regions, retaining only coding regions in the alignment. Table S3 lists the coding regions and their corresponding coordinates in the reference genome (Wuhan-Hu-1, Wu et al. 2020a). After removing the stop codon for each coding region, both the reduced and entire alignments for the complete coding region included 29,232 nucleotide sites.

#### S3.3 Detailed Description of Phylodynamic Analyses

##### Estimating a Dated Phylogeny for the Reduced SARS-CoV-2 Dataset

###### Overview

In this section, we describe the analyses that we performed to infer a dated phylogeny for the reduced sample of COVID-19 viruses. We use the phylogeny inferred from these analyses both in our simulation study (Section S2) and also in our subsequent analyses to evaluate candidate biogeographic models (see *Evaluating Candidate Biogeographic Models*).

###### Model specification

We inferred a dated phylogeny by performing Bayesian analyses of the reduced SARS-CoV-2 sequence dataset under a relaxed-clock model, which includes three main components: (1) a substitution model; (2) a branch-rate prior model; and (3) a branching-process prior model. Below, we describe each of these model components and the corresponding priors for the parameters of those models (note that we used an empirical Bayesian approach to specify non-default priors for several parameters; *i.e.*, where the results of preliminary analyses and/or published results were used to specify the parameters of priors. Details of the priors are described in Table S4.

*Substitution model*

The substitution model collectively describes the process of molecular evolution of the SARS-CoV-2 genomes over the branches of the phylogeny. The process of molecular evolution is apt to vary among regions of these viral genomes. For example, the ORF1ab gene of SARS-CoV-2 encodes nonstructural proteins and is therefore likely to have been subjected to strong purifying selection (Li et al. 2020b), whereas other genes, such as the spike (S) gene, encode structural proteins that determine antigenicity and other immune properties of SARS-CoV-2, and are therefore likely to have been subjected to strong positive selection (Korber et al. 2020; Plante et al. 2020; Hou et al. 2020; Volz et al. 2021). Accordingly, we specified a partitioned substitution model to accommodate possible variation in the evolutionary process across genomic regions. Specifically, we partitioned the SARS-CoV-2 genomes into six data subsets, with three subsets for the ORF1ab gene region (one for each codon position), and three subsets for the remaining ten combined gene regions (one for each codon position). (For a complete list of the gene regions and their corresponding coordinates in the reference genome, see Table S3).

For each of these data subsets, we specified an independent TN93 substitution model (Tamura and Nei 1993), with transition-transversion rate-ratio parameters $\kappa_1$ and $\kappa_2$ (the instantaneous rates of A to G and C to T substitutions, respectively, relative to the transversion rate) and $\pi$ (the stationary frequency of each nucleotide). For each transition-transversion rate-ratio parameter, we specified lognormal priors with a prior mean of 3.74 and a 95% prior interval of $[0.56, 13.04]$.

Table S4: **Priors used to estimate a dated phylogeny of the sampled SARS-CoV-2 sequences.**

| Parameter | Description | Prior |
|---|---|---|
| $\kappa_1$ | Ratio of the A $\rightarrow$ G rate to the transversion rate | Lognormal($\mu = 1.0, \sigma = 0.8$)* |
| $\kappa_2$ | Ratio of the C $\rightarrow$ T rate to the transversion rate | Lognormal($\mu = 1.0, \sigma = 0.8$) |
| $\boldsymbol{\pi}$ | Nucleotide stationary frequencies | Dir(1, 1, 1, 1) |
| $\boldsymbol{m}$ | Partition-specific rate multipliers | Dir(1, 1, 1, 1, 1, 1) |
| $\alpha$ | Shape and scale parameter of the $\Gamma_4$ distribution | Lognormal($\mu = -2.1, \sigma = 0.5874$) |
| $\mathbb{E}[\boldsymbol{r}]$ | Mean of the UCLN | Lognormal($\mu = -12.7, \sigma = 0.5874$) |
| $SD(\boldsymbol{r})$ | Standard deviation of the UCLN | Exp($\lambda = 1/(2.0\text{e}{-6})$) |
| $N_T$ | Effective number of infected individuals at sampling time, $T$ | Lognormal($\mu = 7.5, \sigma = 1.0$) |
| $r$ | Exponential growth rate of the coalescent model | Laplace(0.07, 0.01) |

*$\mu$ and $\sigma$ in this table are the mean and standard deviation of the normal distribution.

To accommodate possible variation in the overall rate of substitution *between* gene regions, we specified independent rate multipliers for each of the six data subsets. To accommodate variation in substitution rates across sites *within* each data subset, we specified a discrete-gamma model (Yang 1994). Our preliminary analyses specified an independent discrete-gamma model for each of the six data subsets, which revealed a similar degree of among-site rate variation within each data subset (*i.e.*, with similar posterior estimates for the six $\alpha$-shape parameters). Accordingly, to decrease model complexity, we specified a shared, discrete-gamma model for the entire alignment. We specified a lognormal hyperprior on the $\alpha$-shape parameter, with a prior mean of 0.15 and a 95% prior interval spanning one order of magnitude around the mean, $[0.039, 0.39]$. This prior reflects our expectation of a high degree of substitution-rate variation across sites, motivated by our observation that most sites in our SARS-CoV-2 alignment are invariant, while a small number of sites appear to be highly variable.

*Branch-rate model*

The branch-rate model describes how the overall substitution rate varies across branches of the tree. Our composite relaxed-clock model specifies the uncorrelated lognormal (UCLN) branch-rate prior model (Drummond et al. 2006; Li and Drummond 2012; Rannala and Yang 2007), which accommodates variation in the overall substitution rate across branches by drawing i.i.d. rate multipliers for each branch from a shared underlying lognormal distribution, where the parameters of this distribution (mean and standard deviation) are estimated from the data. For the mean of the UCLN, we specified a lognormal hyperprior with an expectation of 3.63e−6 substitutions/site/day and 95% prior interval of [0.96e−6, 9.64e−6], motivated by published substitution-rate estimates of approximately 30 substitutions/genome/year (*cf.* Duchene et al. 2020). We specified an exponential hyperprior on the standard deviation of the UCLN such that the branch-specific substitution rates are expected to vary over approximately one order of magnitude.

*Branching-process model*

The branching-process model describes the prior distribution of tree topologies and divergence times. We used a coalescent model with exponential population growth as our branching-process model. This model assumes that the viral population size grows as a deterministic exponential function (Beaumont 1999; Drummond et al. 2002), which is motivated by the fact that our SARS-CoV-2 dataset was sampled from the early, explosive stage of the COVID-19 pandemic. This model is completely described by two free parameters: $N_T$, the effective number of infected individuals in the population at the sampling time, $T$ (*i.e.*, the last sampling date in our dataset, Mar. 8, 2020), and $r$, the exponential growth rate. We specified empirically informed and biologically realistic priors on these parameters.

*Prior on the exponential growth rate, $r$.* We specified a prior on $r$ using external information about the $R_0$, the basic reproductive number, and $\tau$, the duration of the infectious period; these quantities are related through the equation $r = (R_0 - 1)/\tau$. We specified a Laplace prior on $r$, with location and scale parameter values specified according to published estimates of $R_0$ and $\tau$ for COVID-19 (Chinazzi et al. 2020; Li et al. 2020a; Hao et al. 2020; Vaughan et al. 2020; Nadeau et al. 2021; Wölfel et al. 2020; van Kampen et al. 2021; Byrne et al. 2020). This prior on $r$ directly translates to an expected population doubling time, $t = \ln(2)/r$, of 10.6 days (with 95% prior interval ranging from 6.9 to 17.2 days).

*Prior on the effective population size, $N_T$.* Given a population doubling time of $t$, the expected number of individuals at time $T$ is $N_T = N_0 2^{(T-T_0)/t}$, where $N_0$ is the number of individuals at the beginning of the process, and $T_0$ is the origin time of the process. (This equation follows from the fact that there are $(T - T_0)/t$ doubling cycles in a period of duration $T - T_0$.) We therefore specified a prior on $N_T$ informed by our previously determined prior on $t$, as well as several realistic values for $N_0$ (1 or 2) and $T_0$ (some time in late Nov., 2019). Based on these values, we chose a lognormal prior on $N_T$ such that the mean was 2981 and 95% prior interval spanned $[254, 12840]$. Given that there were at least 3940 reported cases on Mar. 8, 2020 alone, it may seem unreasonable to specify a prior such that the expected number of individuals is as low as 2981. However, we note that $N_T$ represents the *effective* number of infected individuals in the population, which is typically substantially smaller than the *total* number of infected individuals in the population. Additionally, we note that: (1) the posterior-mean estimate of $N_T$ under this prior is ≈801, indicating that, if anything, this prior mean is too high, and; (2) sensitivity analyses suggested that posterior estimates of $N_T$ were not very sensitive to this prior (results not shown).

**Parameter estimation**

We performed six independent MCMC simulations to approximate the joint posterior distribution of the relaxed-clock model parameters using `BEAST` version 1.10.5 (Suchard et al. 2018) with the `BEAGLE` library (compiled from the 'hmc-clock' branch, commit 'dd36bf5'; Ayres et al. 2019) to accelerate computation. We ran each replicate MCMC simulation for 50 million generations, sampling continuous parameters every 1000 generations and trees every 10,000 generations. Details of these analyses (*e.g.*, proposal weights) are available in the XML scripts included in our GitHub and Dryad repositories. After discarding the first 20% as the burn-in from each replicate simulation, we combined the remaining posterior samples of trees from all the replicates and then down-sampled every 50,000 generations using `LogCombiner` version 1.10.5. Following initial inspection of the log files using `Tracer` (Rambaut et al. 2018) version 1.7.1, we further evaluated MCMC performance using the `coda` package (Plummer et al. 2006) in `R` (R Core Team 2020). We assessed convergence of replicate MCMC simulations by calculating the ESS for each continuous parameter for the combined posterior samples; ensuring that values for the substitution-model parameters were all $\gg$10000 and those for the branch-rate and branching-process models were all $\gg$200. We then used `TreeAnnotator` version 1.10.5 to generate a summary phylogeny from the combined posterior sample of trees—as a maximum clade credibility (MCC) tree—where the age of each internal node is computed by marginalizing over the age of that node across all samples. Note that as the age of each node is summarized independently across the posterior distribution of trees, it is possible for the MCC summary tree to have negative branch lengths (*i.e.*, where an ancestral node is younger than its descendant node). To avoid potential issues caused by this phenomenon in downstream analyses, we assigned a small positive value (0.001 days) as the duration of these "time-traveling" branches.

#### Evaluating Candidate Biogeographic Models

##### Overview

In this section, we describe our analyses to explore candidate biogeographic models that describe the geographic progression of the SARS-CoV-2 virus during the early phase of the COVID-19 pandemic. We begin by defining the space of candidate models that we will evaluate, and then describe the analyses that we performed to assess both the *relative fit* (by computing Bayes factors to compare competing models) and the *absolute fit* (using posterior-predictive simulation) of these candidate biogeographic models to our reduced SARS-CoV-2 dataset. In evaluating candidate biogeographic models, we condition on the MCC summary phylogeny inferred using the reduced dataset described above (see Section S3.3: *Estimating a Dated Phylogeny for the Reduced SARS-CoV-2 Dataset*).

##### Candidate biogeographic models

*Specifying priors for biogeographic models*

For a biogeographic history with $k$ discrete areas, the stochastic process of geographic dispersal over the branches of the tree is fully specified by a $k \times k$ instantaneous-rate matrix, $\mathbf{Q}$, where an element of the matrix, $q_{ij}$, is the instantaneous rate of change between state $i$ and state $j$ (*i.e.*, the instantaneous rate of dispersal from area $i$ to area $j$). Each element, $q_{ij}$, of the instantaneous-rate matrix, $\mathbf{Q}$, is specified as:

$$q_{ij} = r_{ij}\delta_{ij},$$

where $r_{ij}$ is the rate of dispersal between areas $i$ and $j$, and $\delta_{ij}$ is an indicator variable that takes one of two states (1 or 0); when $\delta_{ij} = 1$, a dispersal route from area $i$ to area $j$ exists, when $\delta_{ij} = 0$ it does not. The total number of dispersal routes, $\sum \delta_{ij}$, for a given biogeographic model is denoted $\Delta$. We used an asymmetric $\mathbf{Q}$ matrix (Edwards et al. 2011) that allows the rate of dispersal from area $i$ to area $j$ to be different from the rate of dispersal from area $j$ to area $i$ (*i.e.*, $r_{ij}$ can be different from $r_{ji}$, and $\delta_{ij}$ can also be different from $\delta_{ji}$). By convention, we rescale the $\mathbf{Q}$ matrix such that the expected number of dispersal events in one time unit is equal to the parameter $\mu$ (Yang 2014). We specified the root frequency $\omega$—the prior probability of the geographic area at the root—as a stochastic random variable to be estimated from the data.

*Prior on the number of dispersal routes.* We specified a Poisson prior on the number of dispersal routes, $\Delta$, with the rate parameter of the Poisson distribution, $\lambda = \binom{k}{2}$, representing a prior belief that half of all possible dispersal routes are included in the biogeographic model; this results in a relatively flat prior probability that any given dispersal route exists for all values of $k$.

*Prior on the average dispersal rate.* Recall that the rate matrix, $\mathbf{Q}$, is rescaled so that the average rate of dispersal between all areas is $\mu$. For a tree of length $T$ (*i.e.*, the sum of the durations of all branches in the tree), the expected number of dispersal events is $\mu \times T$. Therefore, the prior on $\mu$ represents our prior belief about the number of dispersal events over the tree. We specified an exponential prior on $\mu$ with rate parameter $\theta$, and a mean of $1/\theta$. Rather than assuming a fixed value for the mean of the exponential prior, we treat it as a random variable to be estimated from the data. Specifically, we specified a gamma hyperprior on $1/\theta$; this gamma hyperprior has shape parameter $\alpha = 0.5$ and rate parameter $\beta = 0.5$ (enforcing the shape and rate parameters to be equal ensures that the resulting prior on $\mu$ is proper). The resulting prior—known as the $K$-distribution (Jakeman and Pusey 1978)—is a rather diffuse prior on $\mu$, as is the resulting prior distribution on the number of dispersal events.

Table S5: **Priors used in evaluating candidate biogeographic models.**

| Parameter | Description | Prior |
|---|---|---|
| $\Delta_l$ | Number of dispersal routes in interval $l$ | Pois(253) |
| $\mu_l$ | Average dispersal rate in interval $l$ | $\text{Exp}(1/\lambda)$; $\lambda \sim \Gamma(0.5, 0.5)$ |
| $r_{ij,l}$ | Relative dispersal rate from $i$ to $j$ in interval $l$ | $\Gamma(1,1)$ |
| $\boldsymbol{\omega}$ | Root frequencies | $\text{Dir}(1,1,\ldots,1)$ |

*Space of candidate biogeographic models*

We explored a pool of nine candidate biogeographic models. These models assign interval-specific parameters—for the average rate of viral dispersal, $\mu$, and/or relative rates of viral dispersal, $\mathbf{Q}$—to one, two, four, or five pre-specified time intervals; *i.e.*, $1\mu1\mathbf{Q}$, $2\mu1\mathbf{Q}$, $1\mu2\mathbf{Q}$, $2\mu2\mathbf{Q}$, $4\mu1\mathbf{Q}$, $1\mu4\mathbf{Q}$, $4\mu4\mathbf{Q}$, $5\mu5\mathbf{Q}$, and $5\mu5\mathbf{Q}^*$. For example, $4\mu1\mathbf{Q}$ is an interval-specific biogeographic model that allows the average dispersal rate to vary among the four time intervals (but assumes that the relative dispersal rates are constant among intervals). Conversely, $1\mu4\mathbf{Q}$ assumes interval-specific relative dispersal rates (but assumes a constant average dispersal rate across the four time intervals). The $4\mu4\mathbf{Q}$ model may be viewed as a composite of former two models, as it allows *both* the average *and* relative dispersal rates to vary independently among the four intervals.

We specified interval boundaries based on external information regarding events within the study period that might plausibly impact viral dispersal dynamics, including: (A) start of the Spring Festival travel season in China (the highest annual period of domestic travel, Jan. 12); (B) onset of mitigation measures in Hubei province, China (Jan. 26); (C) onset of international air-travel bans against China (Feb. 2), and; (D) relaxation of domestic travel restrictions in China (Feb. 16). Biogeographic models with two intervals include event C, models with four intervals include events A, C, and D, and the $5\mu5\mathbf{Q}$ model includes all four events. The final candidate model, $5\mu5\mathbf{Q}^*$, includes five arbitrary and uniform (bi-weekly) intervals. (See Section S3.2 for additional details on these time intervals.)

**Evaluating the models**

*Assessing relative fit of candidate biogeographic models using Bayes factors*

We evaluated the *relative fit* of each candidate biogeographic model to our SARS-CoV-2 dataset using Bayes factors. This Bayesian model-comparison approach requires that we first estimate the marginal likelihood for each candidate biogeographic model, and then compute the Bayes factor for each pair of competing models as twice the difference in their log marginal likelihoods (Kass and Raftery 1995). We estimated marginal likelihoods for each candidate biogeographic model using both thermodynamic-integration (Lartillot and Philippe 2006) and stepping-stone (Xie et al. 2011; Baele et al. 2012) estimators. These marginal-likelihood estimators tend to be unstable when inferring the phylogeny and biogeographic history jointly, owing to the diffuse (hyper)priors on node-age and branch-rate model parameters, as well as the vast tree space (see Baele et al. 2015). Accordingly, we estimated marginal likelihoods for our candidate biogeographic models by conditioning on the summary phylogeny (the MCC tree) that we inferred using sequence data alone (see Section S3.3).

For each candidate biogeographic model, we first ran eight replicate power-posterior MCMC simulations in `BEAST` (Suchard et al. 2018) with the `BEAGLE` library (version 3.2.0; Ayres et al. 2019). For the constant-rate phylodynamic models, we used `BEAST` version 1.10.5; for the interval-specific phylodynamic models, we used our extended version of `BEAST` (see Section S1). The accuracy of marginal-likelihood estimates using power posteriors depends on the number of powers as well as the number of generations per power (Xie et al. 2011). Therefore, to assess the reliability of our marginal-likelihood estimates, we used an increasing number of powers and an increasing number of generation per power (so the specific values are represented as ranges below) across replicates and checked the variation of estimates among replicates. Specifically, for each replicate power-posterior MCMC simulation, we used 36–64 powers placed at evenly-spaced quantiles of a $\text{Beta}(0.3, 1.0)$ distribution. For each power, we discarded the initial 65000–160000 generations as burn-in and then sampled every 100 generations over the remaining 210000–480000 generations.

To assess the stability of the marginal-likelihood estimates, we also set up a "golden run" for the power-posterior analysis under each model with a large number of powers (128, placed at evenly-spaced quantiles of a $\text{Beta}(0.3, 1.0)$ distribution) and a large number of generations (three million) per power. In the interest of time, here we ran the `BEAST` analyses under each power in parallel by specifying a single XML script per power and running them independently; for each analysis we ran four replicate MCMCs, each of length one million generations with the first 25% discarded as burnin. We combined the output of each independent run to produce the output of the golden run. We then subsampled the golden-run output under each model either by the number of powers or by the number of generations per power to produce a sequence of shorter runs with either fewer powers $(8, 16, 32, 64)$ while holding the number of generations per power same as the golden run, or fewer generations per power $(10000, 25000, \ldots, 2000000)$ while holding the number of powers same as the golden run. We examined the sequence of the estimated marginal likelihood under each candidate biogeographic model as a function of the number of powers and as a function of the number of generations per power to check the convergence behavior.

After confirming the marginal-likelihood estimate under each model converged, we combined the output from all the power-posterior analyses (including the initial eight replicates as well as the golden run) to compute a single marginal likelihood for each model. Details of these analyses (*e.g.*, proposal weights) are available in the XML scripts included in our GitHub and Dryad repositories.

*Assessing absolute fit of candidate biogeographic models using posterior-predictive simulation*

We assessed the *absolute fit* of each candidate biogeographic model to our reduced SARS-CoV-2 dataset using posterior-predictive simulation (Gelman et al. 1996). We first estimated the joint posterior probability distribution of parameters for the candidate model from the observed biogeographic dataset, and then we performed simulations using the parameter estimates randomly drawn from the inferred joint posterior distribution. We used the time-slice parsimony and tipwise-multinomial statistics (as described in Section S1.3) to assess the adequacy (*i.e.*, absolute fit) of each candidate model.

*Estimating the joint posterior probability distribution for each candidate biogeographic model.* For each of the candidate biogeographic models, we first inferred the joint posterior distribution from the observed biogeographic data (*i.e.*, the geographic location of each of the sequences in our reduced SARS-CoV-2 dataset) by performing four independent MCMC simulations using `BEAST` (Suchard et al. 2018) with the `BEAGLE` library (version 3.2.0; Ayres et al. 2019). Specifically, the analyses under the constant-rate ($1\mu1\mathbf{Q}$) model were performed using `BEAST` version 1.10.5, whereas those under the interval-specific biogeographic models were performed using our modified version of `BEAST` (see Section S1). For each replicate MCMC simulation, we ran 10 million generations, sampling every 2000 generations. We discarded the initial 10% of samples (as burn-in) from each replicate MCMC, and then combined the remaining posterior samples from all the replicates using `LogCombiner` version 1.10.5. We then assessed MCMC performance for the resulting composite posterior sample by inspecting the log files using `Tracer` (Rambaut et al. 2018) version 1.7.1 and the `coda` package (Plummer et al. 2006) in `R` (R Core Team 2020). We ensured that the computed ESS values for all continuous parameters were $\gg$100. Details of these analyses are available in the XML scripts included in our GitHub and Dryad repositories.

*Posterior-predictive simulations.* For each candidate biogeographic model, we simulated $m = 2500$ predictive datasets by repeatedly sampling at random from the corresponding joint posterior probability distribution. We then generated posterior-predictive distributions from each set of $m$ predictive datasets for 20 separate summary statistics. These 20 statistics include time-slice variants (with 10 time slices) of the two (parsimony and tip-wise multinomial) summary statistics. We specified 10 (weekly) time slices spanning the early phase of COVID-19 (where the first slice covers the period from the origin of SARS-CoV-2 to Jan. 5, 2020, and the remaining nine weekly slices spanning the period between Jan. 6 and Mar. 8). For each posterior-predictive distribution, we computed the posterior-predictive $p$ value (see Section S1.3) to assess the adequacy (*i.e.*, absolute fit) of the corresponding biogeographic model.

**Results**

Our golden-run experiments demonstrate that our marginal-likelihood estimates converged to stable values (Fig. S13). Bayes-factor comparisons of all candidate models decisively support (*i.e.*, $2 \ln \mathrm{BF} \gg 10$; Table S6) the 4-interval ($4\mu4\mathbf{Q}$) biogeographic model. The preference for this model is corroborated by the results of our posterior-predictive simulations: $4\mu4\mathbf{Q}$ was inferred to provide an adequate absolute fit to our reduced SARS-CoV-2 dataset for every summary statistic, whereas all less complex models (*i.e.*, with fewer interval-specific parameters for the average dispersal rates, $\mu$, and/or the relative dispersal rates **Q** (*i.e.*, were all inferred to be inadequate by at least two of the 20 time-slice summary statistics (Fig. S14). Accordingly, we use the $4\mu4\mathbf{Q}$ model for our joint phylodynamic analyses of the entire SARS-CoV-2 dataset described below (see Section S3.3).

*The relative fit of competing biogeographic models to the reduced SARS-CoV-2 dataset*

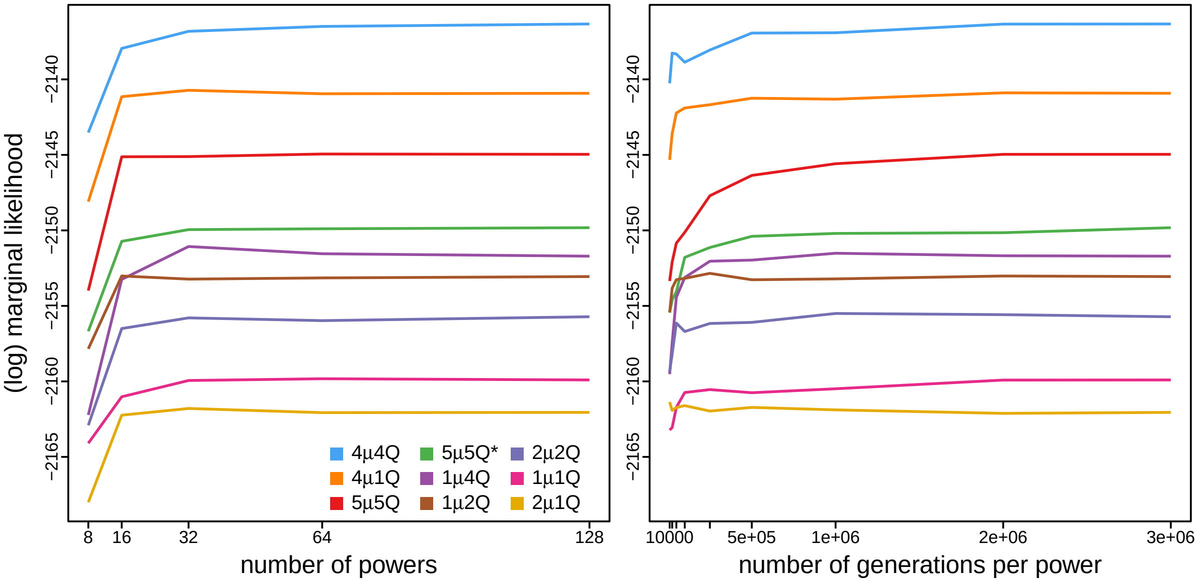


Figure S13: **Convergence of marginal-likelihood estimates of the candidate biogeographic models.** Convergence of the marginal-likelihood estimate under each candidate biogeographic model as a function of the number of powers (left panel) and the number of generations per power (right panel). To assess the convergence of the marginal-likelihood estimate, we set up a 'golden run' under each model where a large number of powers (128, placed at evenly-spaced quantiles of a $\text{Beta}(0.3, 1.0)$ distribution) and a large number of generations (three million) per power are used. We then subsampled each golden run either by the number of powers or by the number of generations per power to produce a sequence of shorter runs with either fewer powers $(8, 16, 32, 64$; while holding the number of generations per power same as the golden run) or fewer generations $(10000, 25000, 50000, \ldots, 2000000$; while holding the number of powers same as the golden run) per power. Each colored line shows the sequence of the estimated log marginal likelihood under a given model plateauing as the number of powers increases (left) or the number of generations per power increases (right). The settings (including number of powers and the number of generations per power) of the golden run appear to be sufficient to obtain stable marginal-likelihood estimates. The 4-interval ($4\mu4\mathbf{Q}$) model appears to be consistently preferred over all the other models across all settings.

Table S6: **Marginal-likelihood estimates of (and Bayes factor comparisons among) the candidate biogeographic models.** Column 1 lists the candidate biogeographic models. Column 2 lists the composite marginal-likelihood estimates (computed by combining the samples from replicate power-posterior MCMC simulations). The last column lists the inferred support ($2 \ln \text{BF}$) of the alternative interval-specific models compared to the constant-rate model ($1\mu1\mathbf{Q}$). The preferred biogeographic model ($4\mu4\mathbf{Q}$) is indicated in bold text.

| model | ln marginal likelihood | $2 \ln \text{BF}$ compared to $1\mu1\mathbf{Q}$ |
|---|---|---|
| $1\mu1\mathbf{Q}$ | −2159.57 | — |
| $2\mu1\mathbf{Q}$ | −2162.03 | −4.92 |
| $1\mu2\mathbf{Q}$ | −2152.87 | 13.40 |
| $2\mu2\mathbf{Q}$ | −2155.77 | 7.60 |
| $4\mu1\mathbf{Q}$ | −2141.03 | 37.08 |
| $1\mu4\mathbf{Q}$ | −2152.01 | 15.13 |
| $\mathbf{4\mu4Q}$ | **−2136.58** | **45.99** |
| $5\mu5\mathbf{Q}$ | −2144.60 | 29.94 |
| $5\mu5\mathbf{Q}^*$ | −2149.60 | 19.94 |

*Absolute fit of competing biogeographic models to the reduced SARS-CoV-2 dataset*

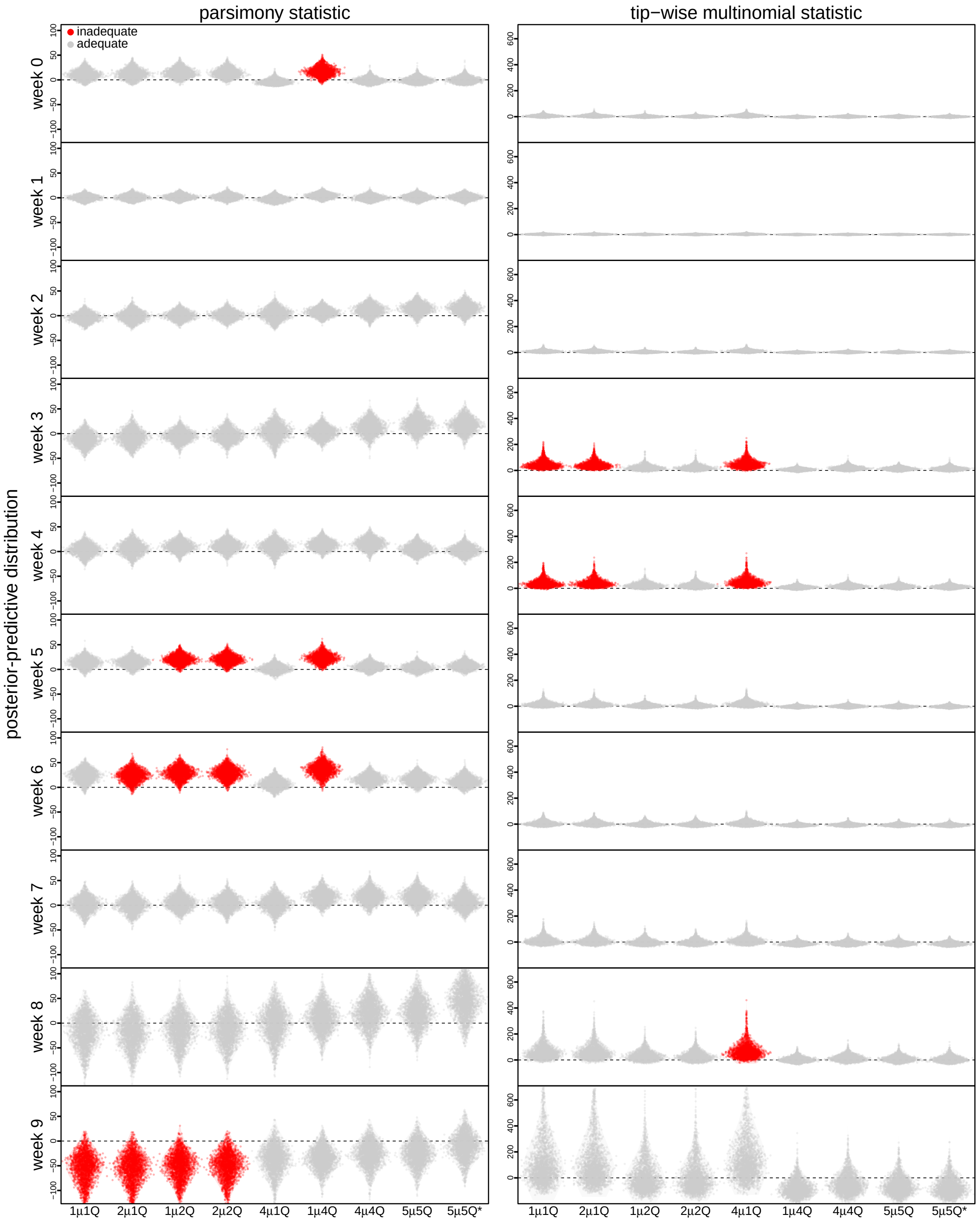


Figure S14: **Posterior-predictive distributions under the candidate biogeographic models.** Each column of panels corresponds to one of the two types of summary statistics (parsimony and tip-wise multinomial); each row of panels corresponds to one of the 10 (weekly) time slices. Each panel includes a set of nine violin plots (one for each of the candidate biogeographic models listed in Table S6). Each violin plot depicts the posterior-predictive distribution of the 2500 replicate simulations for the corresponding summary statistic under the corresponding candidate model. Each dot represents the value of the corresponding summary statistic for a single replicate posterior-predictive simulations, where the value is the discrepancy between the summary statistic for the observed dataset and the single simulated dataset. The horizontal dashed line indicates the value of the summary statistic under identical fit of the simulated and observed datasets. The violin plots in red indicate that the corresponding model provides an inadequate fit to the SARS-CoV-2 dataset (*i.e.*, it is incapable of generating geographic datasets that are similar to the observed data) under the corresponding time-slice summary statistic, as its 95% posterior-predictive interval does not overlap with the dashed line.

#### Joint Analyses of the Entire SARS-CoV-2 Dataset

##### Overview

In this section, we describe the analyses we performed to infer the joint posterior probability distribution of the phylodynamic model—comprising all parameters of the component relaxed-clock and biogeographic models—from the entire SARS-CoV-2 dataset (which includes the viral genome sequences, and the geographic areas and dates of viral sampling). For these analyses, we specified a relaxed-clock model that was similar to that used to estimate the dated phylogeny for the reduced SARS-CoV-2 dataset (Section S3.3), and specified the biogeographic model that was selected based on analyses of the reduced dataset (Section S3.3). Below, we provide details on: (1) the specified phylodynamic model; (2) the MCMC simulations we performed to estimate the joint posterior under this model, and; (3) the posterior-predictive simulations we performed to assess the absolute fit of the biogeographic model to the entire SARS-CoV-2 dataset.

##### Model specification

Our joint analyses of the entire SARS-CoV-2 dataset are based on a phylodynamic model that includes (1) a relaxed-clock model, and (2) a biogeographic model. The relaxed-clock model that we specified for our joint analyses of the entire SARS-CoV-2 sequence dataset is identical to that specified previously in our analyses of the reduced SARS-CoV-2 sequence dataset (see Section S3.3) with minor changes to the priors to accommodate differences in viral sampling (see Table S7). The biogeographic model that we specified for our joint analyses of the entire SARS-CoV-2 geographic dataset is identical to the biogeographic model that we selected previously based on analyses of the reduced SARS-CoV-2 dataset (see Section S3.3); specifically, the 4-interval ($4\mu4\mathbf{Q}$) model.

Table S7: **Priors used to jointly infer SARS-CoV-2 phylogeny and biogeographic history for the entire dataset.**

| Parameter | Description | Prior |
|---|---|---|
| $\kappa_1$ | Ratio of the A $\rightarrow$ G rate to the transversion rate | Lognormal$(\mu = 1.0, \sigma = 0.8)^*$ |
| $\kappa_2$ | Ratio of the C $\rightarrow$ T rate to the transversion rate | Lognormal$(\mu = 1.0, \sigma = 0.8)$ |
| $\boldsymbol{\pi}$ | Nucleotide stationary frequencies | Dir$(1, 1, 1, 1)$ |
| $\boldsymbol{m}$ | Partition-specific rate multipliers | Dir$(1, 1, 1, 1, 1, 1)$ |
| $\alpha$ | Shape and scale parameter of the $\Gamma_4$ distribution | Lognormal$(\mu = -2.1, \sigma = 0.5874)$ |
| $\mathbb{E}[\boldsymbol{r}]$ | Mean of the UCLN | Lognormal$(\mu = -12.5, \sigma = 0.5)$ |
| $SD(\boldsymbol{r})$ | Standard deviation of the UCLN | Exp$(\lambda = 1/(4.0\text{e}{-6}))$ |
| $N_T$ | Effective number of infected individuals at sampling time, $T$ | Lognormal$(\mu = 7.0, \sigma = 1.0)$ |
| $r$ | Exponential growth rate of the coalescent model | Laplace$(0.07, 0.01)$ |
| $\Delta_l$ | Number of dispersal routes in interval $l$ | Pois$(253)$ |
| $\mu_l$ | Average dispersal rate in interval $l$ | Exp$(1/\lambda)$; $\lambda \sim \Gamma(0.5, 0.5)$ |
| $r_{ij,l}$ | Relative dispersal rate from $i$ to $j$ in interval $l$ | $\Gamma(1, 1)$ |
| $\boldsymbol{\omega}$ | Root frequencies | Dir$(1, 1, \ldots, 1)$ |

$^*\mu$ and $\sigma$ in this table are the mean and standard deviation of the normal distribution.

##### Data analysis

*Estimating the joint posterior of phylodynamic model parameters using MCMC simulation*

We performed 20–30 independent MCMC simulations to approximate the joint posterior distribution of the phylodynamic-model parameters—including the phylogeny, divergence times, and biogeographic history—from the entire SARS-CoV-2 dataset using our modified version of `BEAST` (see Section S1) with the `BEAGLE` library (compiled from the 'hmc-clock' branch, commit 'dd36bf5'; Ayres et al. 2019) to accelerate computation. We ran each replicate MCMC simulation for 10–20 million generations, sampling continuous parameters every 1000 generations and the dated phylogeny every 10,000 generations. When a phylogeny was sampled, we performed stochastic mapping using the endpoint-conditioned uniformization algorithm (Hobolth and Stone 2009) and our modified algorithm to perform stochastic mapping under interval-specific models (see Section S1) to simulate dispersal histories over the sampled tree. Details of these analyses are available in the XML scripts included in our GitHub and Dryad repositories.

After discarding the first 10–75% of samples from each replicate MCMC simulation (as burn-in), we combined the remaining posterior samples of trees from all replicates and then down-sampled every 50,000 generations using `LogCombiner` version 1.10.5. Following initial inspection of the log files using `Tracer` (Rambaut et al. 2018) version 1.7.1, we further evaluated MCMC performance using the `coda` package (Plummer et al. 2006) in `R` (R Core Team 2020). We assessed convergence of replicate MCMC simulations by calculating the ESS for each continuous parameter for the combined posterior samples; ensuring that values for the substitution-model parameters were all $\gg$4000, those for the geographic model parameters were all $\gg$200, and that the ESS values for all parameters of the branch-rate and branching-process models were all $\gg$100.

*(Re)assessing adequacy of the biogeographic model using posterior-predictive simulation*

We previously established that the preferred biogeographic model provides an adequate description of the process of geographic dispersal during the early phase of the COVID-19 pandemic (see Section S3.3). However, those analyses were based on the reduced (rather than entire) SARS-CoV-2 dataset, and also conditioned on a single dated phylogeny—the MCC tree inferred in Section S3.3—rather than integrating over the posterior probability distribution of dated phylogenies. Accordingly, we performed additional posterior-predictive simulation to confirm that the preferred biogeographic model provides an adequate fit to the entire SARS-CoV-2 dataset under an inference scenario where geographic history is jointly integrated over the posterior distribution of dated phylogenies.

We performed a series of posterior-predictive simulations to assess the adequacy of the preferred biogeographic model ($4\mu4\mathbf{Q}$). As a point of reference, we also assessed the absolute fit of the constant-rate ($1\mu1\mathbf{Q}$) biogeographic model (*c.f.*, Table S6). For both biogeographic models, we simulated $m = 2500$ posterior-predictive datasets by repeatedly sampling at random from the corresponding joint posterior distribution of phylodynamic model parameters inferred from the entire SARS-CoV-2 dataset. We then generated posterior-predictive distributions from each set of $m$ predictive datasets under 20 separate statistics include the two (parsimony and tip-wise multinomial) summary statistics, each computed over 10 time slices. We specified 10 (weekly) time slices spanning the early phase of COVID-19 (where the time first slice covers the period from the origin of SARS-CoV-2 to Jan. 5, 2020, and the remaining nine weekly time slices spanning the period between Jan. 6 and Mar. 8). For each posterior-predictive distribution, we computed the posterior-predictive $p$ value (see Section S1.3) to assess the adequacy (*i.e.*, absolute fit) of the corresponding biogeographic model.

*Quantifying differences between prior and posterior distributions*

The discrete-geographic phylodynamic model has many parameters, which raises questions about our ability to infer the parameters from a single set of biogeographic observations. If there is insufficient information in the biogeographic data to estimate the parameters, we expect the posterior distribution of each model parameter to resemble its prior distribution. To quantify the degree to which the posterior distribution of the interval-specific model is updated by the data, we computed Kullback–Leibler (KL) divergence between the marginal posterior and the prior distributions of each pairwise relative dispersal rate under each of the constant-rate and preferred models. We represent the KL divergence as $D_{KL}(P\,||\,Q)$, where $P$ indicates the posterior distribution and $Q$ indicates the prior distribution.

We also used a symmetric version of the KL divergence—$D_{KL}(P\,||\,Q) + D_{KL}(Q\,||\,P)$—to quantify the difference in the inferred posterior distributions of the pairwise relative dispersal rates between candidate biogeographic models. In this case, $P$ represents the posterior distribution of one model, and $Q$ represents the posterior distribution of the other model. We focused on three pairs of models ($4\mu4\mathbf{Q}$ versus $1\mu1\mathbf{Q}$, $4\mu4\mathbf{Q}$ versus $4\mu1\mathbf{Q}$, and $1\mu4\mathbf{Q}$ versus $1\mu1\mathbf{Q}$) with different relative-rate intervals to assess the impact of allowing relative rates of dispersal to vary across intervals; we also examined such difference using two pairs of models ($4\mu 1\mathbf{Q}$ versus $1\mu 1\mathbf{Q}$, and $4\mu 4\mathbf{Q}$ versus $1\mu 4\mathbf{Q}$) with identical relative-rate intervals to assess the impact of allowing average dispersal rate to vary across intervals on the relative-rate estimates.

As we used BSSVS in our inferences, each pairwise relative dispersal rate, $q_{ij}$, is drawn from a mixture of discrete (when $\delta_{ij} = 0$) and continuous (when $\delta_{ij} = 1$) distributions. The probability density function is:

$$P(q_{ij}) = P(\delta_{ij} = 0) + P(\delta_{ij} = 1)P(r_{ij}). \tag{S10}$$

The KL divergence of distribution $P$ from distribution $Q$ for parameter $q_{ij}$ is then computed as:

$$\begin{aligned} D_{KL}^{q_{ij}}(P \,||\, Q) &= \int P(q_{ij}) \log \frac{P(q_{ij})}{Q(q_{ij})} dq_{ij} \\ &= P(\delta_{ij} = 0) \log \frac{P(\delta_{ij} = 0)}{Q(\delta_{ij} = 0)} + \int P(\delta_{ij} = 1) P(r_{ij}) \log \frac{P(\delta_{ij} = 1) P(r_{ij})}{Q(\delta_{ij} = 1) Q(r_{ij})} dr_{ij} \\ &= P(\delta_{ij} = 0) \log \frac{P(\delta_{ij} = 0)}{Q(\delta_{ij} = 0)} + P(\delta_{ij} = 1) \log \frac{P(\delta_{ij} = 1)}{Q(\delta_{ij} = 1)} + P(\delta_{ij} = 1) \int P(r_{ij}) \log \frac{P(r_{ij})}{Q(r_{ij})} dr_{ij}. \end{aligned} \tag{S11}$$

We computed each component of (S11) from the sampled distribution. The last component is the KL divergence between two continuous distributions; we computed it using a conventional approach based on the empirical cumulative distribution function (Pérez-Cruz 2008).

*Parameter summary*

We summarized the number of viral dispersal events between a given pair of geographic areas by counting the number of dispersal events from the source region (*e.g.*, China) to the destination region (*e.g.*, North America) that occurred on that day for a given simulated history, and then looped over all of the histories to obtain the posterior distribution of the number of pairwise dispersal events. Mean and 95% credible intervals for the daily number of viral dispersal events were then computed from the corresponding posterior distribution.

**Results**

Posterior-predictive simulations confirm that the preferred interval-specific biogeographic model ($4\mu 4\mathbf{Q}$) provides an adequate fit to the entire SARS-CoV-2 dataset, whereas the constant-rate biogeographic model is inferred to be inadequate (Fig. S15). The results of these joint analyses of the entire SARS-CoV-2 dataset are presented in the main text (Figs. 6–8) and Figs. S16–S18.

The computed KL divergence between the posterior and prior distributions shows that, under the interval-specific ($4\mu 4\mathbf{Q}$) model, the most recent interval—the interval with much longer total branch length and more dispersal events than the previous three intervals—appears to contain the most information in inferring the relative dispersal rates, and be comparable to the counterpart under the constant-rate model. The average amount of information gain in the first three intervals appear to be much more limited than the last interval, with noticeable exceptions (*e.g.*, Hubei to East China in interval 2, Japan and Korea to West USA and Canada in interval 3) which also show less information gain under the constant-rate model (Fig. S19).

The inferred posterior distributions of the relative rates of dispersal appear to be much more similar between the pair of comparing models who share the relative-rate intervals (Fig. S23) than between the pair of models with different relative-rate intervals (Figs. S20–S22), indicating that the observed differences in the relative-rate estimates between preferred interval-specific ($4\mu 4\mathbf{Q}$) and the constant-rate ($1\mu 1\mathbf{Q}$) models result from allowing the relative dispersal rates, instead of the average dispersal rate, to vary across intervals.

*Absolute fit of the constant-rate (1$\mu$1**Q**) and preferred (4$\mu$4**Q**) models to the entire SARS-CoV-2 dataset*

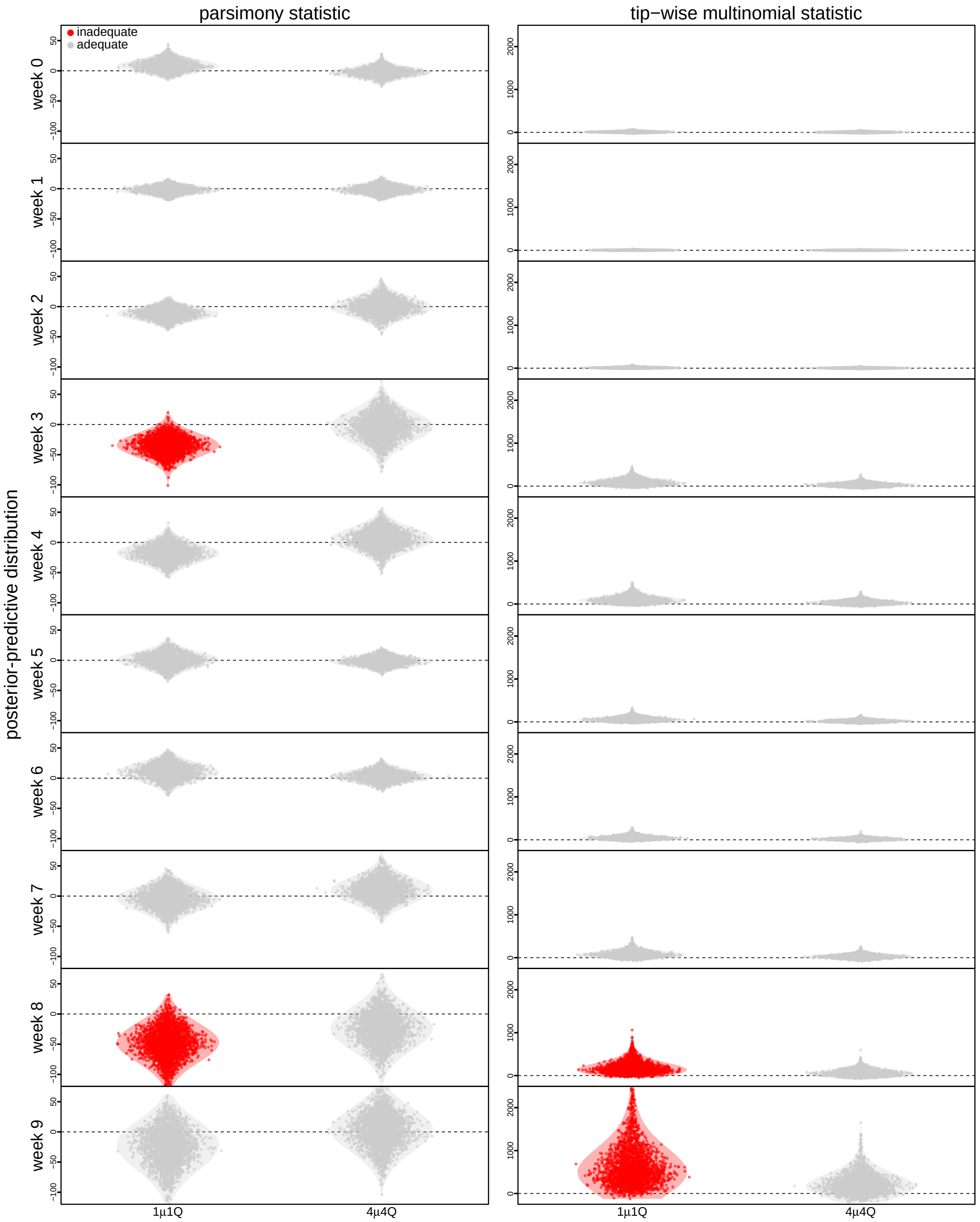


Figure S15: **Posterior-predictive distributions of biogeographic models under joint inference of the entire SARS-CoV-2 dataset.** Each column of panels corresponds to one of the two types of summary statistics (parsimony and tip-wise multinomial); each row of panels corresponds to one of the 10 (weekly) time slices. Each panel includes a two violin plots for the preferred (4$\mu$4**Q**, right) and constant-rate (1$\mu$1**Q**, left) biogeographic models. Each violin plot depicts the posterior-predictive distribution of the 2500 replicate simulations for the corresponding summary statistic under the corresponding candidate model. Each dot represents the value of the corresponding summary statistic for a single replicate posterior-predictive simulations, where the value is the discrepancy between the summary statistic for the observed dataset and the single simulated dataset. The horizontal dashed line indicates the value of the summary statistic under identical fit of the simulated and observed datasets. The violin plots in red indicate that the corresponding model provides an inadequate fit to the SARS-CoV-2 dataset (*i.e.*, it is incapable of generating geographic datasets that are similar to the observed data) under the corresponding time-slice summary statistic, as its 95% posterior-predictive interval does not overlap with the dashed line.

*Inferred support for dispersal routes under the constant-rate (1$\mu$1**Q**) and preferred (4$\mu$4**Q**) models*

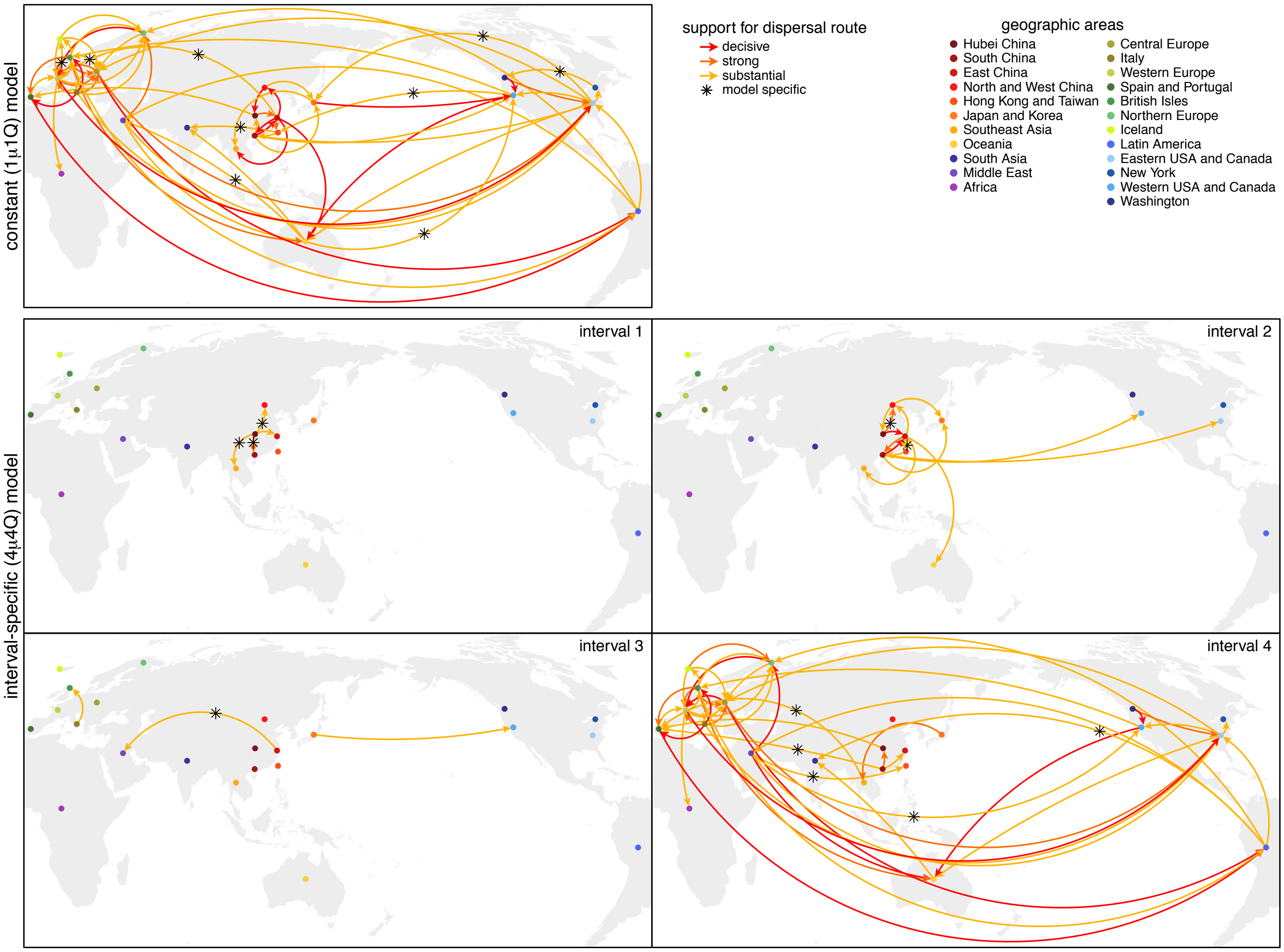


Figure S16: **Variation in viral dispersal routes during the early phase of the COVID-19 pandemic.** Arrows indicate routes inferred to play a significant role in viral dispersal during the early phase of the COVID-19 pandemic; colors indicate the level of evidential support for each dispersal route (as 2ln Bayes factors). The number, duration, and significance of dispersal routes inferred under the constant-rate (1$\mu$1**Q**) model differ strongly from those inferred under the preferred (4$\mu$4**Q**) interval-specific model. By assumption, the constant-rate (1$\mu$1**Q**) model implies an invariant set of dispersal routes. By contrast, the preferred (4$\mu$4**Q**) interval-specific model reveals that the number and intensity of dispersal routes varied over the four intervals. The first interval (Nov. 17–Jan. 12) is dominated by dispersal from Hubei to other areas in China, and the second interval (Jan. 12–Feb. 2) exhibits more widespread international dispersal originating from China. The third interval (Feb. 2–Feb. 16)—immediately following the onset of international air-travel bans with China—exhibits a sustained reduction in the number of dispersal routes. Note that the constant-rate model infers nine spurious dispersal routes (not detected under the interval-specific model). Conversely, the preferred interval-specific model reveals ten significant dispersal routes (not detected under the constant-rate model) that imply a more significant role for Hubei as a source of viral spread in the first and second intervals, and also reveals additional dispersal routes emanating from China (to the Middle East in the third interval and to Spain/Portugal in the fourth interval).

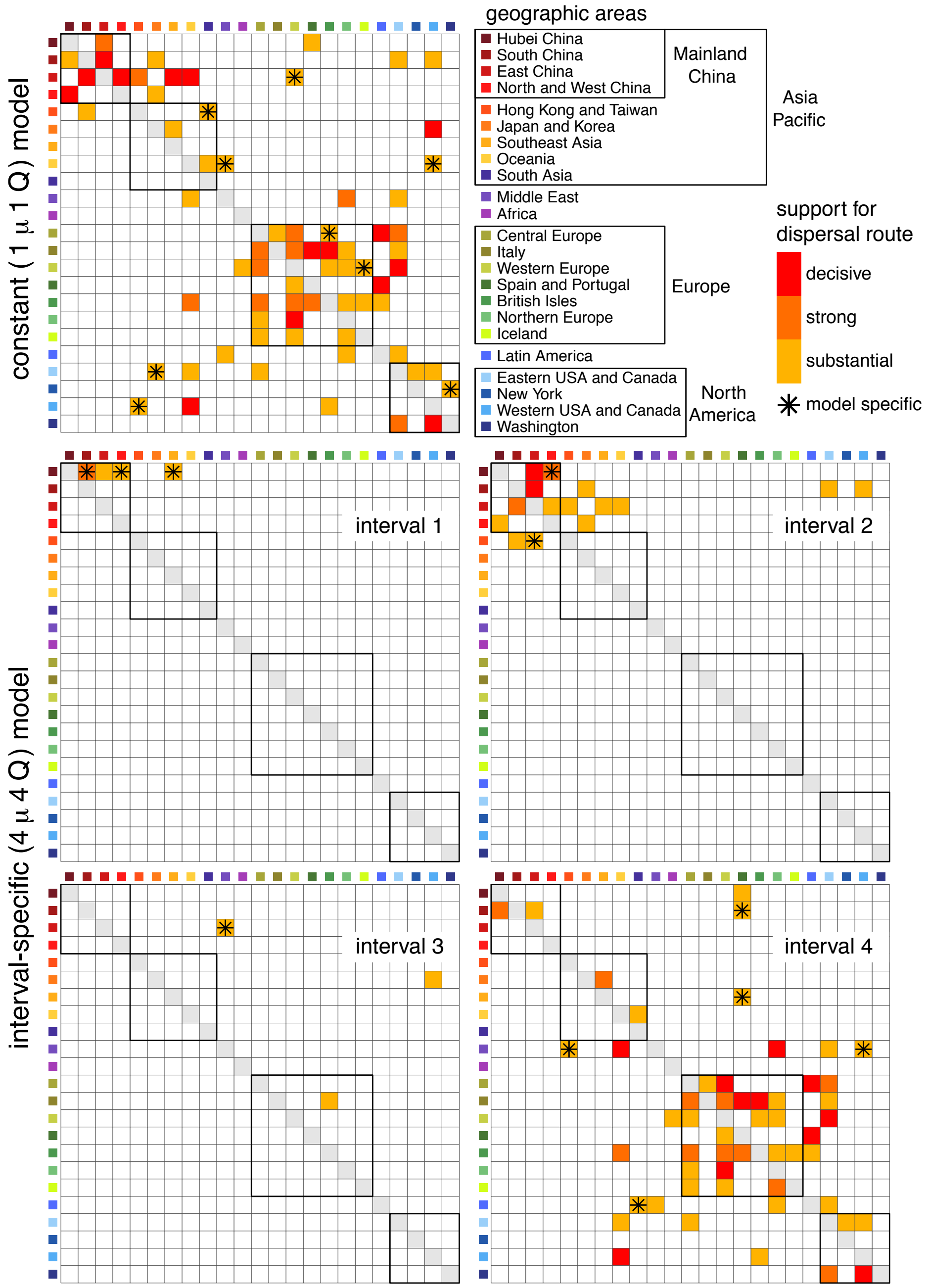


Figure S17: **Variation in viral dispersal routes during the early phase of the COVID-19 pandemic.** This is simply a heatmap representation of Fig. S16 that may improve the clarity of the evidential support for dispersal routes among all 23 study areas during the rarely phase of the COVID-19 pandemic. Each panel is a 23-by-23 matrix, where row $i$ indicates the 'source' area and each $j$ column indicates the 'destination' area, such that each $\delta_{ij}$ element of the matrix indicates the evidential support (as 2ln BF, see inset) for the dispersal route from area $i$ to area $j$. The boxes within each matrix indicate groups of areas within a region (*e.g.*, the four geographic regions of mainland China).

*Inferred pairwise dispersal parameters under the preferred (4$\mu$4**Q**) interval-specific model*

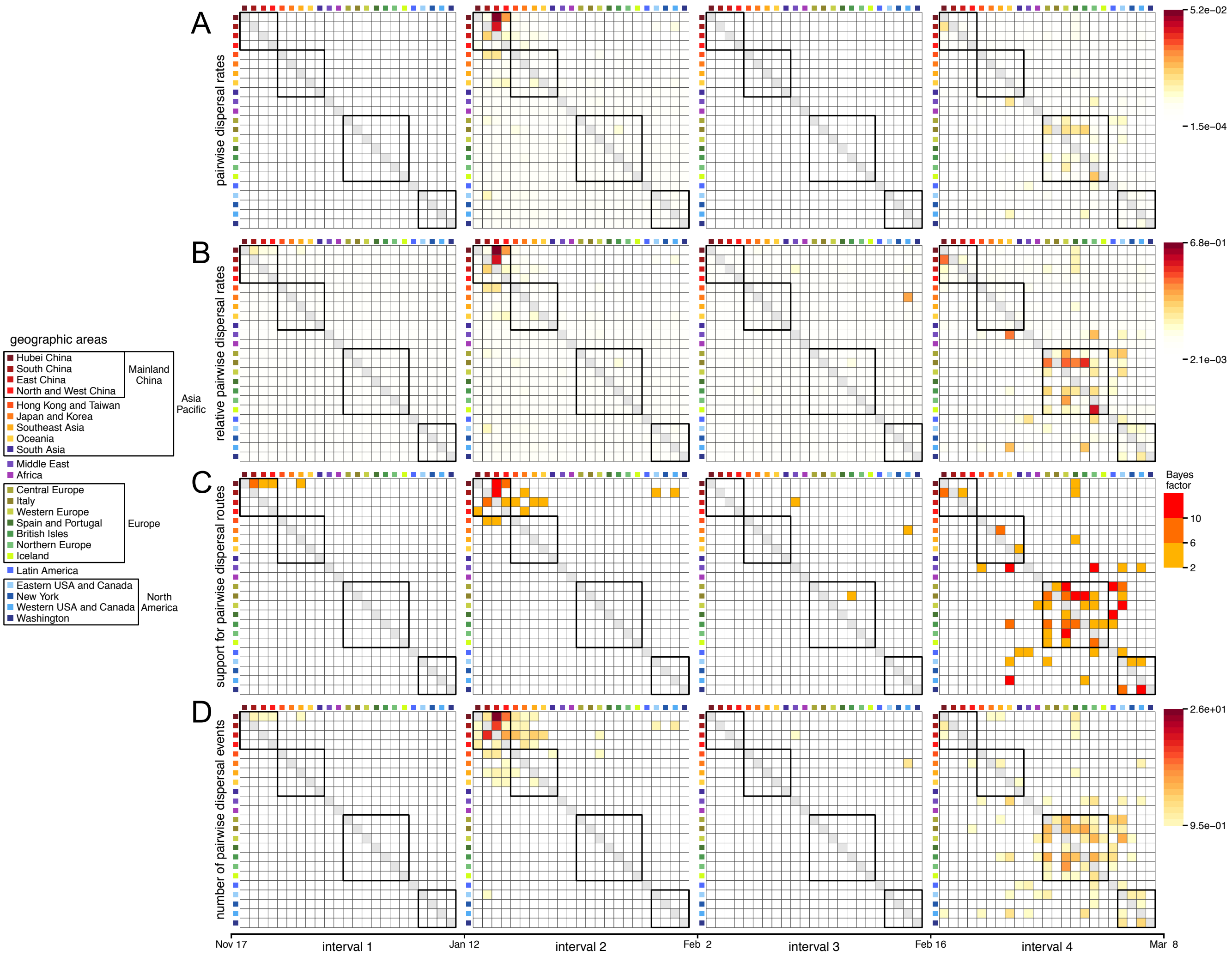


Figure S18: **Summary of dispersal parameters inferred under the preferred interval-specific (4$\mu$4Q) model.** The four time intervals exhibit distinct dispersal dynamics. **(A)** Absolute viral dispersal rate between each pair of discrete geographic areas. **(B)** Relative viral dispersal rates (*i.e.*, the absolute rates in panel A divided by the inferred global dispersal rate for the corresponding interval) between each pair of discrete geographic areas. **(C)** The evidential support (Bayes factors, inset legend, panel C, right) that a given dispersal route played a significant role in the spread of the virus. **(D)** Number of viral dispersal events between each pair of discrete geographic areas. Boxes in each panel indicate groups of areas (inset legend, left). The first interval is dominated by dispersal from Hubei to other areas in China, the second interval by more widespread dispersal within Asia and by dispersal from China to North America, culminating in cosmopolitan dispersal in the fourth interval. Note that interval three—immediately following the onset of international air-travel bans with China—exhibits a large reduction in the number of viral dispersal routes, including disruption of the dispersal routes from China to North America.

*Information gain on pairwise dispersal rates under the constant-rate ($1\mu1\mathbf{Q}$) and preferred ($4\mu4\mathbf{Q}$) models*

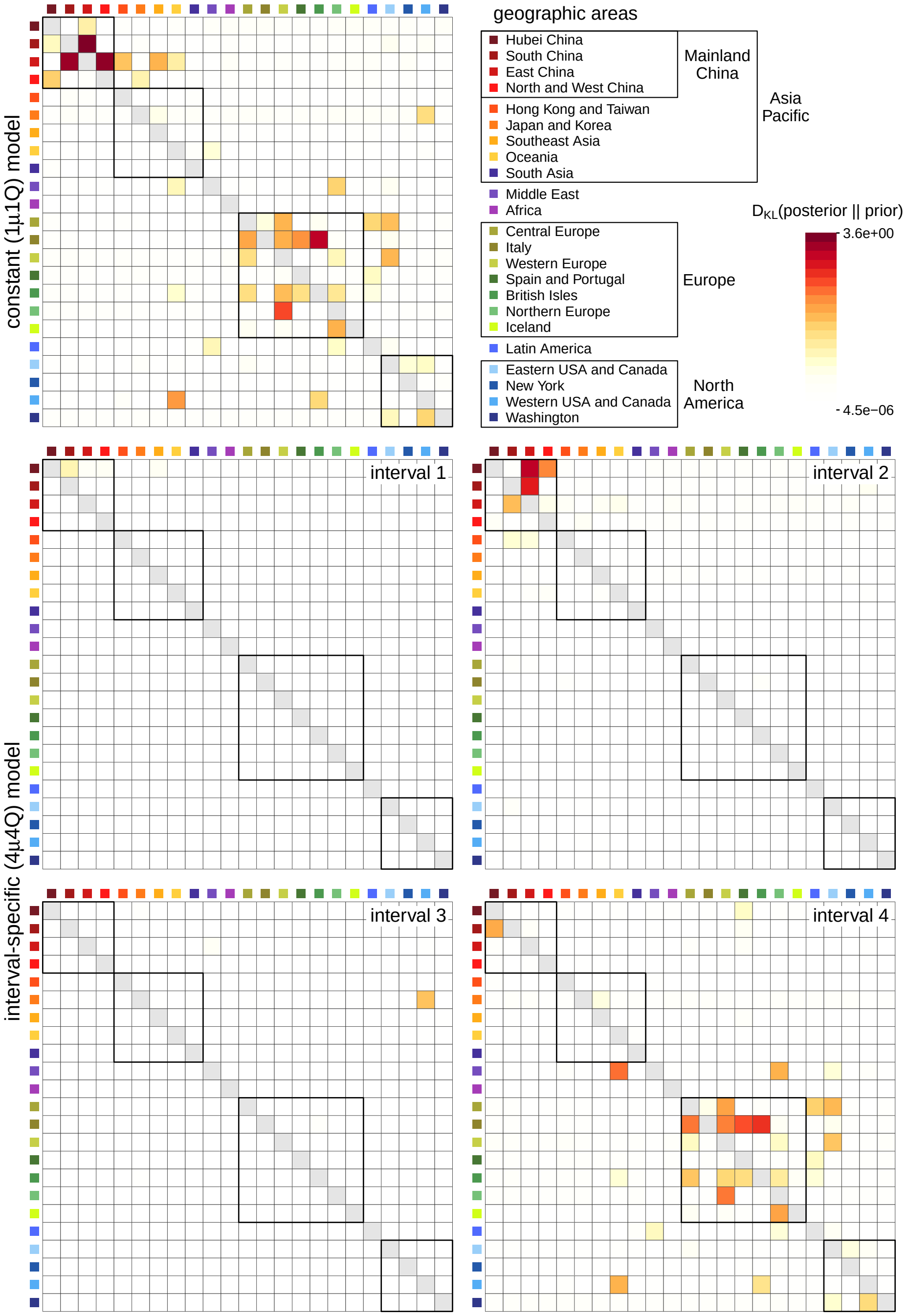


Figure S19: **The information gain on the pairwise relative dispersal rates under the constant-rate ($1\mu1\mathbf{Q}$) and preferred ($4\mu4\mathbf{Q}$) models.** We use Kullback–Leibler (KL) divergence to measure the information gain in moving from the prior to posterior distribution of the pairwise relative dispersal rates. Each panel is a 23-by-23 matrix, where row $i$ indicates the 'source' area and column $j$ indicates the 'destination' area, such that each element of the matrix indicates the KL divergence (colored according to the inset legend bar) between the inferred posterior distribution and the specified prior distribution (which is the same for all pairs) for the relative rate of dispersal from area $i$ to area $j$. The top row shows the information gain under the constant-rate model, while the remaining two rows show such measure under the preferred interval-specific ($4\mu4\mathbf{Q}$) model. Under the interval-specific ($4\mu4\mathbf{Q}$) model, the last interval appears to contain the most information in inferring the relative dispersal rates, and be comparable to the counterpart under the constant-rate model. The average amount of information gain in the first three intervals appear to be much more limited than the last interval, with noticeable exceptions (*e.g.*, Hubei to East China in interval 2, Japan and Korea to West USA and Canada in interval 3) which also show less information gain under the constant-rate model.

*Difference between inferred pairwise dispersal rates under biogeographic models*

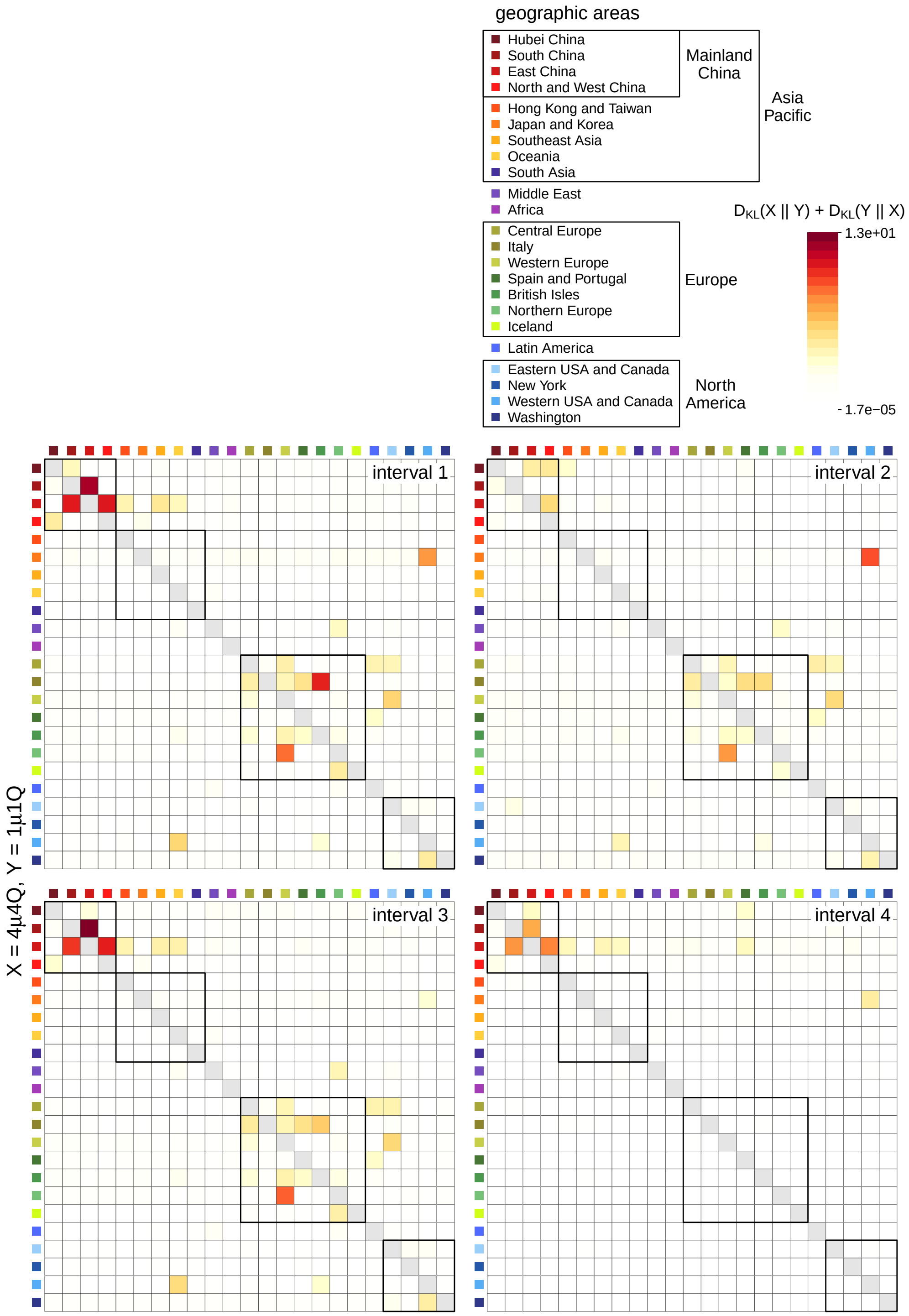


Figure S20: **Difference between inferred pairwise relative dispersal rates under models with different relative-rate intervals (4$\mu$4Q versus 1$\mu$1Q).** We use symmetric KL divergence to measure the difference in the inferred posterior distribution of the pairwise relative dispersal rates between the preferred interval-specific (4$\mu$4**Q**) and constant-rate (1$\mu$1**Q**) models. Each panel is a 23-by-23 matrix, where row $i$ indicates the 'source' area and column $j$ indicates the 'destination' area, such that each element of the matrix indicates the KL divergence (colored according to the inset legend bar; note that the heatmap color scale is shared among Figs. S20–S23) between the inferred posterior distribution of the relative rate of dispersal from area $i$ to area $j$ in the corresponding interval under the interval-specific model and the counterpart under the constant-rate model.

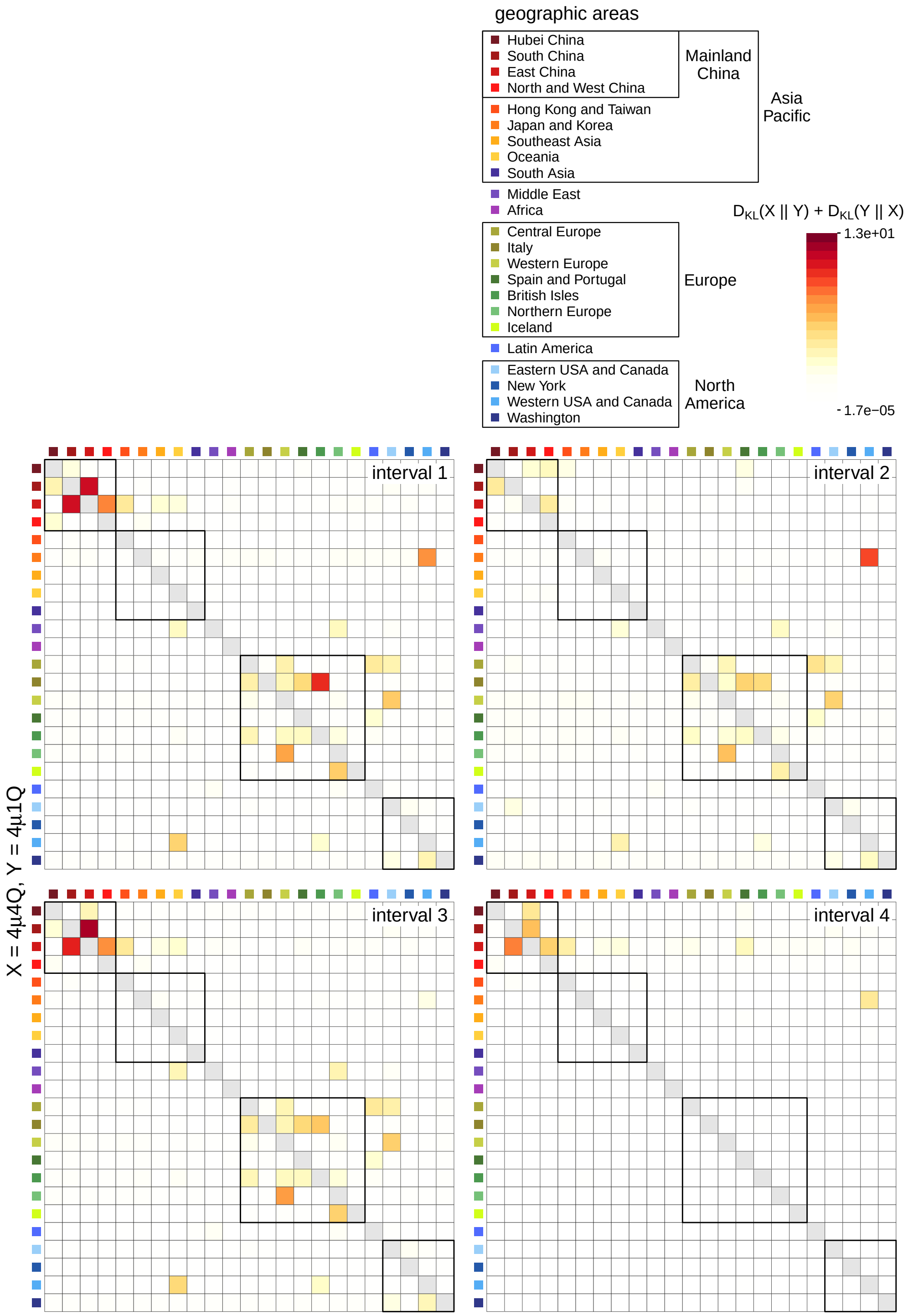


Figure S21: **Difference between inferred pairwise relative dispersal rates under models with different relative-rate intervals (4$\mu$4Q versus 4$\mu$1Q).** We use symmetric KL divergence to measure the difference in the inferred posterior distribution of the pairwise relative dispersal rates between two interval-specific (4$\mu$4**Q** and 4$\mu$1**Q**) models. Each panel is a 23-by-23 matrix, where row $i$ indicates the 'source' area and column $j$ indicates the 'destination' area, such that each element of the matrix indicates the KL divergence (colored according to the inset legend bar; note that the heatmap color scale is shared among Figs. S20–S23) between the inferred posterior distribution of the relative rate of dispersal from area $i$ to area $j$ in the corresponding interval under the 4$\mu$4**Q** model and the counterpart under the 4$\mu$1**Q** model.

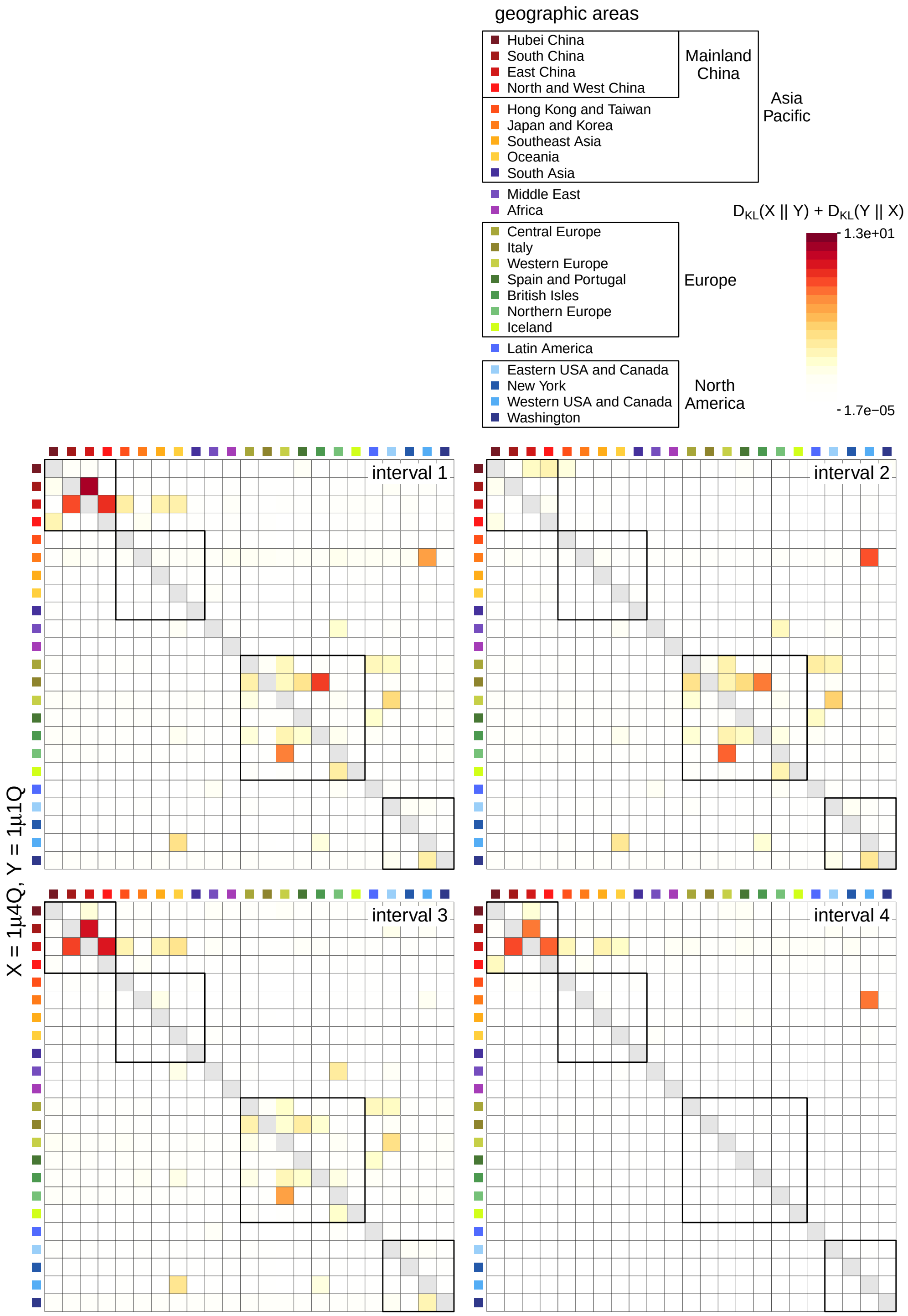


Figure S22: **Difference between inferred pairwise relative dispersal rates under models with different relative-rate intervals (1$\mu$4Q versus 1$\mu$1Q).** We use symmetric KL divergence to measure the difference in the inferred posterior distribution of the pairwise relative dispersal rates between an interval-specific (1$\mu$4**Q**) and the constant-rate (1$\mu$1**Q**) models. Each panel is a 23-by-23 matrix, where row $i$ indicates the 'source' area and column $j$ indicates the 'destination' area, such that each element of the matrix indicates the KL divergence (colored according to the inset legend bar; note that the heatmap color scale is shared among Figs. S20–S23) between the inferred posterior distribution of the relative rate of dispersal from area $i$ to area $j$ in the corresponding interval under the interval-specific model and the counterpart under the constant-rate model.

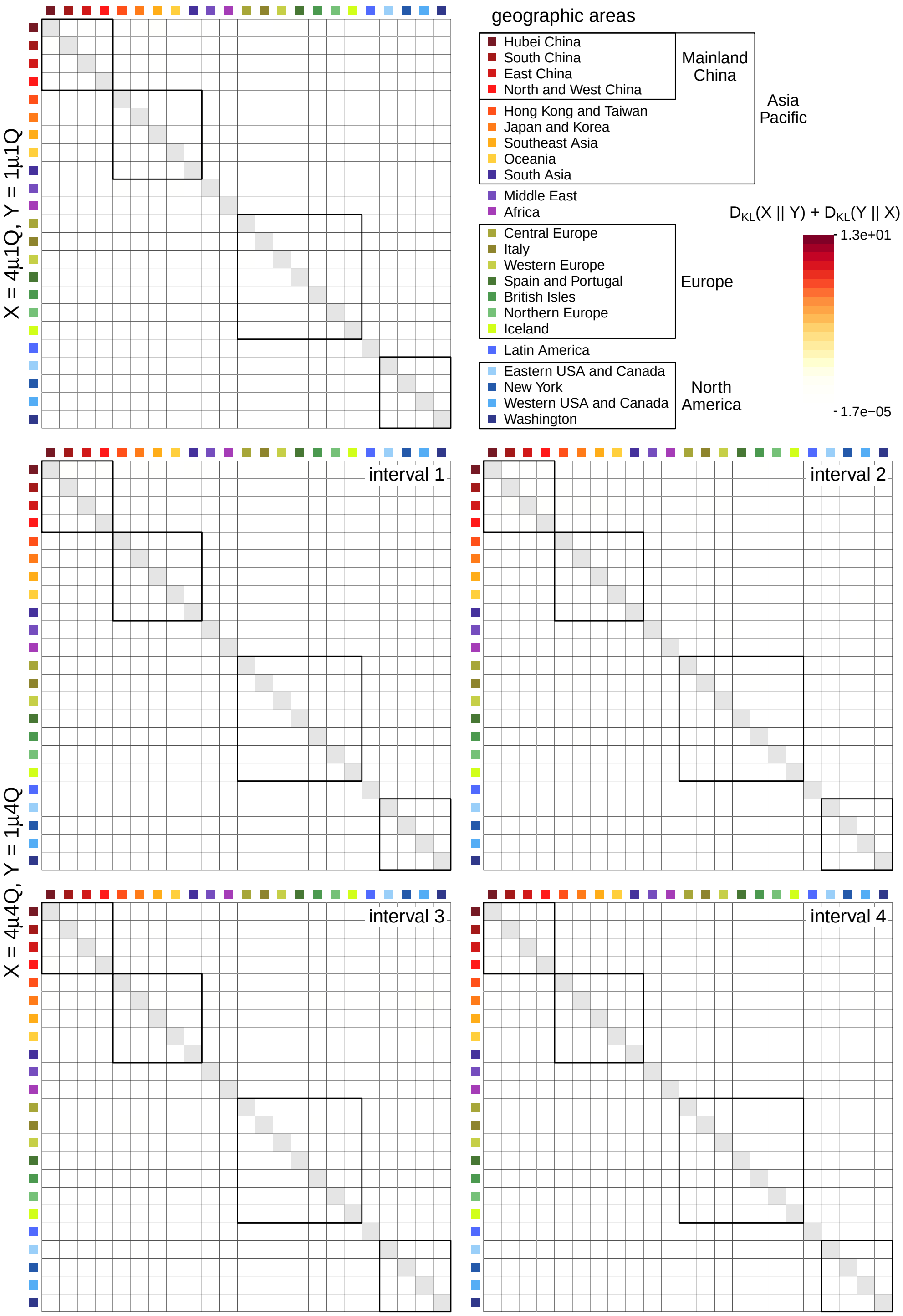


Figure S23: **Difference between inferred pairwise relative dispersal rates under models with the same relative-rate intervals (4$\mu$1Q versus 1$\mu$1Q, top row; 4$\mu$4Q versus 1$\mu$4Q, bottom two rows).** We use symmetric KL divergence to measure the difference in the inferred posterior distribution of the pairwise relative dispersal rates between the models with constant relative dispersal rates (4$\mu$1**Q** and 1$\mu$1**Q**; top row), and between the models with interval-specific relative dispersal rates (4$\mu$4**Q** and 1$\mu$4**Q**; bottom two rows). Each panel is a 23-by-23 matrix, where row $i$ indicates the 'source' area and column $j$ indicates the 'destination' area, such that each element of the matrix indicates the KL divergence (colored according to the inset legend bar; note that the heatmap color scale is shared among Figs. S20–S23) between the inferred posterior distribution of the relative rate of dispersal from area $i$ to area $j$ under the comparing models. The posterior distributions appear to be much more similar when the pair of comparing models share the relative-rate intervals than the pairs with different relative-rate intervals (see Figs. S20–S22).

#### Estimating Daily Global Viral Dispersal Rates

##### Overview

In this section, we describe our analyses to explore the correlation between daily global air-travel volume and global SARS-CoV-2 dispersal rates during the early phase of the COVID-19 pandemic. Because we were able to obtain data on the *daily* volume of global air travel during this period, we performed an analysis of the entire SARS-CoV-2 dataset under a more granular phylodynamic model that allows the average dispersal rate to vary from day to day. We then performed standard statistical tests to assess the degree of correlation between the inferred daily global viral dispersal rates and daily global air-travel volume.

##### Model specification

Our estimates of daily variation in global viral dispersal rates are based on the phylodynamic model ($4\mu4\mathbf{Q}$) that we previously used to infer the joint posterior of SARS-CoV-2 phylogeny and biogeographic history (see Section S3.3), except that we further discretized the number of time intervals in which the global average dispersal rate, $\mu$, was free to vary. Specifically, rather than allowing global average dispersal rate to vary between four time intervals, we specified an independent $\mu$ for each of the 70 days between Dec. 30, 2019 and Mar. 8, 2020 (with an additional independent $\mu$ for the time spanning the origin of SARS-CoV-2 to Dec. 29). The prior on each $\mu$ is specified according to the posterior estimates inferred in Section S3.3. We computed the posterior mean of the global viral dispersal rate across the entire history inferred from the joint analyses and used it as the prior mean, and we specified standard deviation of the prior distribution so that the 95% prior interval spans three orders of magnitude around the mean. Details of the priors are described in Table S8. Our inferences of geographic history under this model were averaged over the marginal posterior probability distribution of dated phylogenies inferred in Section S3.3. Details of these analyses are available in the XML scripts included in our GitHub and Dryad repositories.

##### Data analysis

*Parameter estimation*

We performed 15 independent MCMC simulations to approximate the joint posterior distribution of the biogeographic-model parameters from the entire SARS-CoV-2 dataset using our modified version of `BEAST` (see Section S1) and `BEAGLE` version 3.2.0 (Ayres et al. 2019). We ran each replicate MCMC simulation for 5 million generations, sampling continuous parameters every 2000 generations and the dated phylogeny every 10000 generations. When a phylogeny was sampled, we performed stochastic mapping using the endpoint-conditioned uniformization algorithm (Hobolth and Stone 2009) and our modified algorithm to perform stochastic mapping under interval-specific models (see Section S1) to simulate dispersal histories over the sampled tree. Details of these analyses are available in the XML scripts included in our GitHub and Dryad repositories.

For each replicate MCMC simulation, we discarded the first one million generations (as burn-in), and then combined the remaining posterior samples from all replicates using `LogCombiner` version 1.10.5. Following initial inspection of the log files using `Tracer` (Rambaut et al. 2018) version 1.7.1, we further evaluated MCMC performance using the `coda` package (Plummer et al. 2006) in `R` (R Core Team 2020).

Table S8: **Priors used to infer the daily global viral dispersal rates for the entire dataset.**

| Parameter | Description | Prior |
|---|---|---|
| $\Delta_l$ | Number of dispersal routes in interval $l$ | $\text{Pois}(253)$ |
| $\mu_p$ | Global dispersal rate in day $p$ | $\text{Lognormal}(\mu = -4.76, \sigma = 1.7622)^*$ |
| $r_{ij,l}$ | Relative dispersal rate from $i$ to $j$ in interval $l$ | $\Gamma(1,1)$ |
| $\boldsymbol{\omega}$ | Root frequencies | $\text{Dir}(1,1,\ldots,1)$ |

*$\mu$ and $\sigma$ in this table are the mean and standard deviation of the normal distribution.

We assessed convergence of replicate MCMC simulations by calculating the ESS for each continuous parameter for the combined posterior samples, ensuring that values for all parameters were $\gg$700.

*Correlation test*

We tested for correlation between the volume of daily global air travel, $\boldsymbol{V} = \{v_i\}$ (where $i = \{1, 2, \ldots, m\}$ and $m$ represents the total number of days included in our dataset), and the mean estimate of daily global SARS-CoV-2 dispersal rate, $\boldsymbol{\mu} = \{\mu_i\}$. To remove potential trend or seasonality in the time series of $\boldsymbol{V}$ and $\boldsymbol{\mu}$, we first transformed each of the two time series by taking the difference between each value of the time series and the value a week prior to it. Specifically, we computed $\boldsymbol{v}'$ as $\{v_j - v_{j-7}\}$ (where $j = \{8, \ldots, m\}$) and $\boldsymbol{\mu}'$ as $\{\mu_j - \mu_{j-7}\}$. We then generated various truncated dataset by including values from each of the two differenced time series ($\boldsymbol{v}'$ and $\boldsymbol{\mu}'$) with different start dates (ranging from Jan. 6 to Feb. 17, 2020; *i.e.*, $j$ ranges from 8 to 49) to the same end date (the end of our study period, Mar. 8, 2020; $j = 70$). Finally, we assessed the correlation for each truncated dataset by computing Pearson's $r$ and the corresponding $p$-value to determine the time that the correlation first established.

**Results**

The daily global dispersal rate estimates are presented in the main text (Fig. 6, light blue). Pearson's $r$ and the corresponding $p$-value between the volume of daily global air travel and the estimated mean rate of daily global SARS-CoV-2 dispersal are presented in Fig. S24. The correlation appears to increase when we focus on the time series of February (*i.e.*, discarding the January values), possibly reflecting that the geographic distribution of SARS-CoV-2 was still confined to specific regions (*e.g.*, China and some other Asian countries) prior to this point. The $p$-value increases quickly when we fewer than 25 time points are included in the correlation test, presumably reflecting a decrease in power as the number of data points decreases. Therefore, we report Pearson's $r$ and the corresponding $p$ value between the two time series over the interval from Jan. 31 (when the virus first achieved a cosmopolitan distribution; WHO 2020) to the end of our study period (Mar. 8, 2020) in the main text.

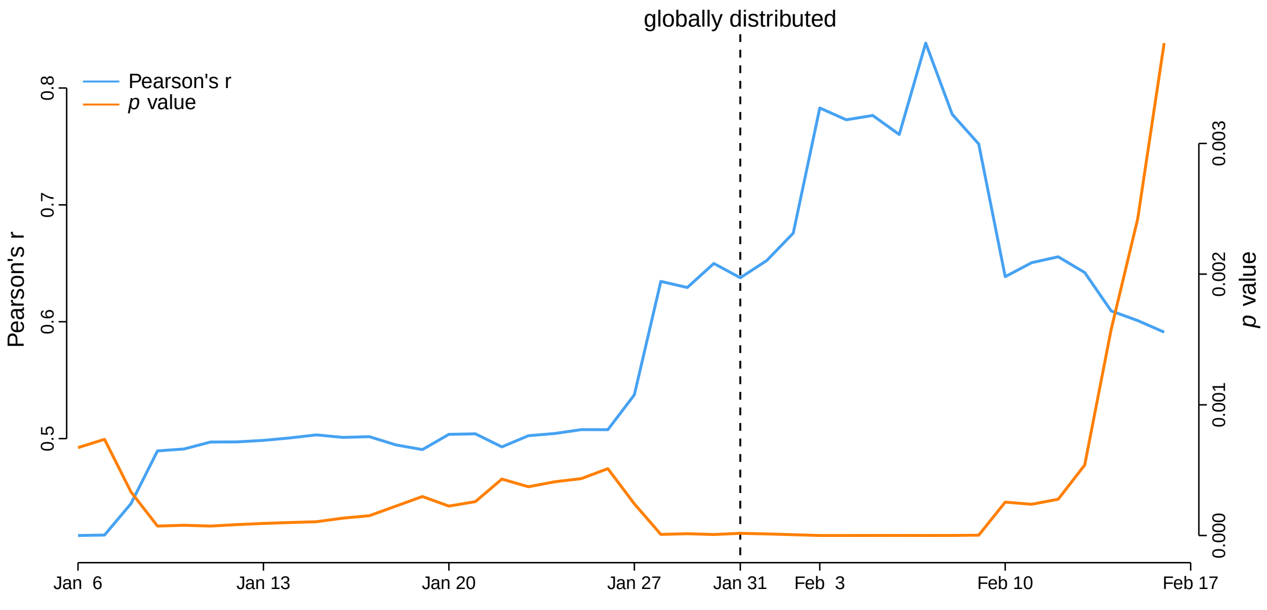


Figure S24: **Correlation between daily global air travel volume and the estimated mean daily global SARS-CoV-2 dispersal rate.** We generated time-series datasets by including values with different start dates (along the x axis) to the same end date (the end of our study period, Mar. 8, 2020). We then computed Pearson's $r$ and the corresponding $p$ value for each dataset, and plotted them as a function of the corresponding start date. The virus first achieved a cosmopolitan distribution on Jan. 31 (dashed line; WHO 2020).
